# Sporadic long-range contacts in out-of-venue settings dominate mass gathering events in Germany

**DOI:** 10.64898/2026.08.19.26359786

**Authors:** Steven Schulz, Alejandra Rincón Hidalgo, Andrzej K. Jarynowski, Marlli Zambrano, Janik Suer, Ashish Thampi, Luca Ferretti, Huynh Thi Phuong, Chao Xu, Rafael Mikolajczyk, Richard Pastor, Veronika K. Jaeger, André Karch, Vitaly Belik

## Abstract

Mass gathering events (MGEs) play a critical role for infectious disease dynamics on a population level as they provide opportunities for superspreading; however, underlying mechanisms remain insufficiently understood. We analyzed nationwide GPS-based, individual-level location data from mobile phone users in Germany between April and August 2024 with 16 m spatial precision. Potentially infectious contacts were inferred from close co-location and linked to contact settings using OpenStreetMap data. Various MGEs, including EURO 2024 matches, major concerts, festivals, and fairs were compared using a common contact metric. Non-football events generated substantially more contacts than football events. While overall national contact numbers remained stable, MGEs produced so-called “small-world” contacts which gather people from distant locations into close proximity and could strongly enhance infectious disease dynamics. Crucially, most high-risk contacts occurred within two hours before the event, not at the event itself, and concentrated in public transport, leisure, and event-adjacent areas. Our work provides the first systematic and comparative evaluation of contact exposure across various types of MGEs and contact settings. Event-type-specific dynamics, particularly indirect and mobility-driven contacts, critically shape infection risk. These insights can inform accurate transmission modeling, targeted intervention and event-management strategies.

## INTRODUCTION

Mass gathering events (MGEs) such as sport tournaments, concerts, and festivals gather large numbers of people into close spatial proximity. While MGEs are a key expression of human behavior and have high social and economic value, they also create ample opportunity for respiratory pathogens to spread in a population [1–3] and draw particular attention from public health authorities [4]. Identifying which types of MGEs and which patterns of human contact cause superspreading is crucial for the understanding and modeling of infectious disease dynamics, standardized risk assessment, and the design of effective, evidence-based interventions during epidemic situations [2, 5].

Contact-tracing has emerged as a new standard besides contact surveys to study contacts in infectious disease epidemiology. However, these data are unsuitable to unveil both general and detailed mechanisms of MGE-induced contacts because they lack spatiotemporal precision or population coverage (i.e., contact-tracing using Bluetooth or Wifi [6–8]), or both (i.e., contact surveys [9–12]). Also, contact-tracing is typically limited to specific venues, times, or experimental settings, such as monitored concerts and sporting events in large arenas [1, 8], isolated communities [13], cruise ships [14], or university campuses [15]. While these studies provide valuable detail, they rely on the strong assumption that controlled environments adequately represent the complex and fluid nature of real-world MGEs, thus lacking generality beyond specific types and settings of MGEs. National-scale mobile phone-based contact-tracing efforts in the UK with broad coverage revealed the seeding of local outbreaks as a consequence of the UEFA EURO 2020 tournament (held in 2021) [7], but lacked spatial and temporal granularity to pinpoint contact dynamics at individual MGEs and ascribe contacts to specific settings and behaviors.

A key question is how contacts change during MGEs compared with normal behavior: Are certain gatherings more likely to create high-risk contacts? Are specific locations, times, or behaviors associated with increased transmission risk? Despite their recognized role in disease spread, detailed data on MGE-driven contacts, their timing, location, and structure within the wider social network, remain limited. Addressing these questions requires real-world data that combines microscopic precision (on the scale of few meters and minutes) to detect and contextualize contacts with macroscopic coverage (over hundreds of kilometers, several months, and substantial fractions of the population) to ensure sufficient data from various MGEs with typically several thousand attendees.

The use of crowd-based GPS data in an epidemiological context is relatively new [16–19]. Previous work established that mobile phone data (GPS or CDR) can help understanding how infections spread [20–22] and inform optimized mitigation strategies, in particular during mass gatherings [23, 24]: It also revealed that human contact patterns arise from population flows, urban mobility and daily routines in cities [19, 25–30], shape disease transmission [21] and correlate with actual transmission dynamics and predict public health outcomes [16, 18, 21, 31–35].

In this study, we use anonymized individual GPS location data from mobile phones of around 0.4 % of the population in Germany (ca. 330,000 mobile phone users) to detect, quantify and compare contacts. Close-range contacts are defined as spatiotemporal co-location of individuals, during various types of MGEs (Fig. S2). This dataset overcomes key limitations of contact-tracing and contact surveys by providing a larger study cohort, a systematic coverage of MGEs, and the exquisite precision of GPS coordinates. We demonstrate how this approach enables a detailed and scalable analysis of how social interactions shift during MGEs, providing a foundation for more targeted and effective public health interventions.

## RESULTS

We analyze GPS location data collected from ca. 330,000 anonymous mobile phone users between April and August 2024, a 0.4 % sample of the German population, and derive contact patterns based on spatiotemporal co-location (Methods, SI Secs. S1, S3 and S6). This time window covers a variety of major MGEs across Germany, most notably the UEFA EURO 2024 football tournament, major concerts, festivals and fairs (Tab. S14).

### MGEs increase contact rates in host cities

Fig. 1 reveals emergent contact peaks in host cities in relation with various MGEs (Figs. S1 and S3), besides other non-periodical effects resulting from public and school holidays. This relation shows that MGEs have a significant effect on the contact rates of host cities (Tab. S2). Remarkably, these peaks arise due to contacts occurring both within and outside of MGE venues, as shown exemplarily for the case of football matches (Fig. 1, red vs. green line). Unlike for cities, Germany-wide data shows that weekly periodicity dominates the effect of MGEs on contact rates (Fig. S4, Tab. S3). These findings suggest that the effect of MGEs on contacts is local within host cities, but extends beyond the MGE venue. This is seemingly in contrast with previous observations from contact-tracing showing that MGEs lead to widespread spikes in potentially infectious contacts [7].

**FIG. 1.**
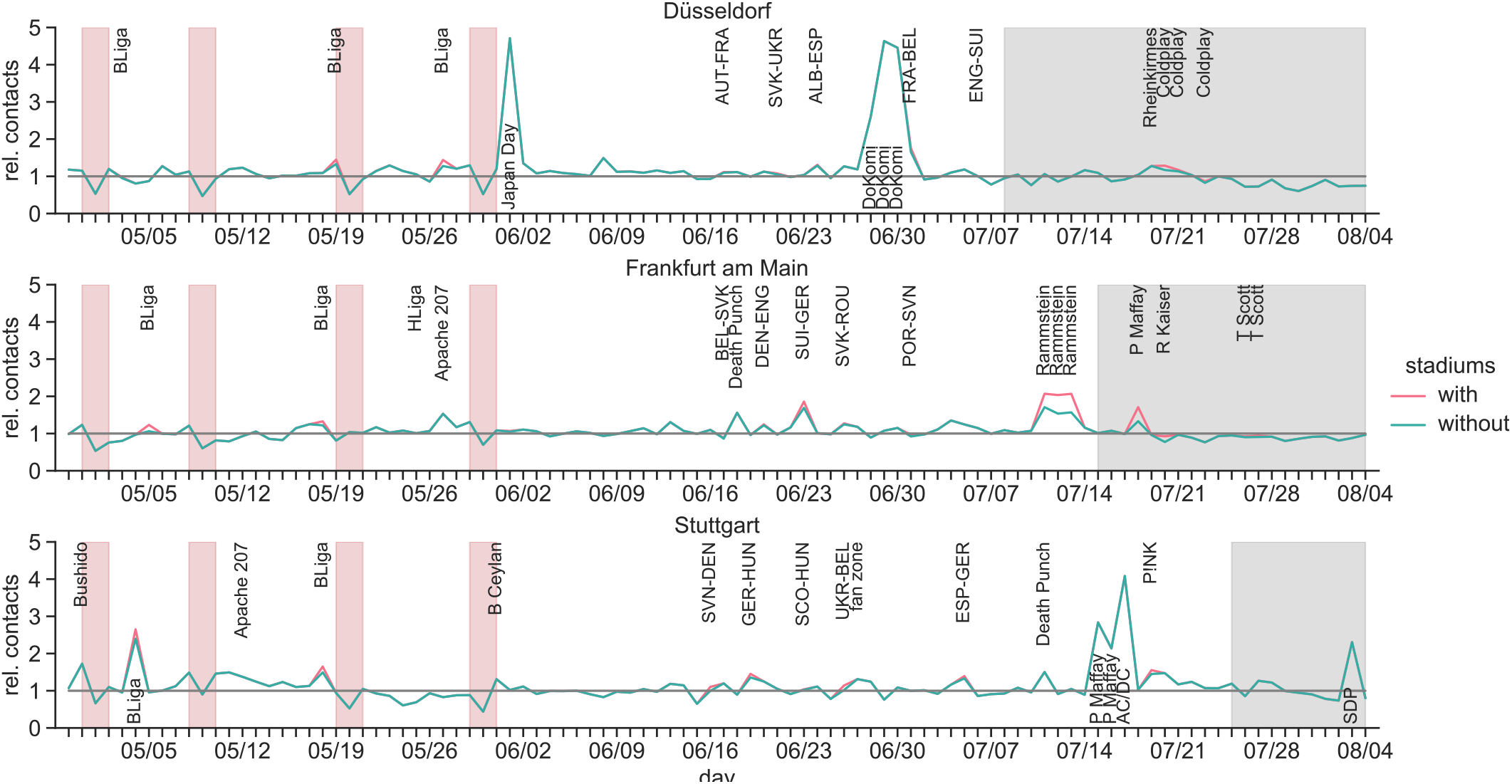
Contact variations in MGE host cities. Relative detrended daily contact levels 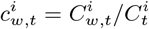 between April 29 and August 4, 2024 (calendar weeks 18 to 31) in the cities of Düsseldorf, Frankfurt and Stuttgart, i.e. 3 of 10 host cities *i* of the UEFA EURO 2024 tournament (rows). The weekday-dependent baseline is given by 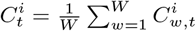. Data is shown when contacts inside host stadiums of the UEFA EURO 2024 tournament in these cities are included (red line) or excluded (green line) from the contact count 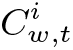 . Public holidays and school holidays are indicated by red and gray shading, respectively (note regional differences in public holiday observance and school holiday schedules). Major MGEs, including football, concerts and festivals are highlighted (Tab. S14). An extended figure covering all 10 host cities of UEFA EURO 2024 and Germany at-large is provided in Figs. S3 and S4, respectively.

### MGEs entail type-dependent out-of-venue contact rates

Fig. 2 compares contact rates within the host city and Germany at-large for various MGEs (Methods, SI Sec. S8; Fig. S6 shows additional data for host venues).

**FIG. 2.**
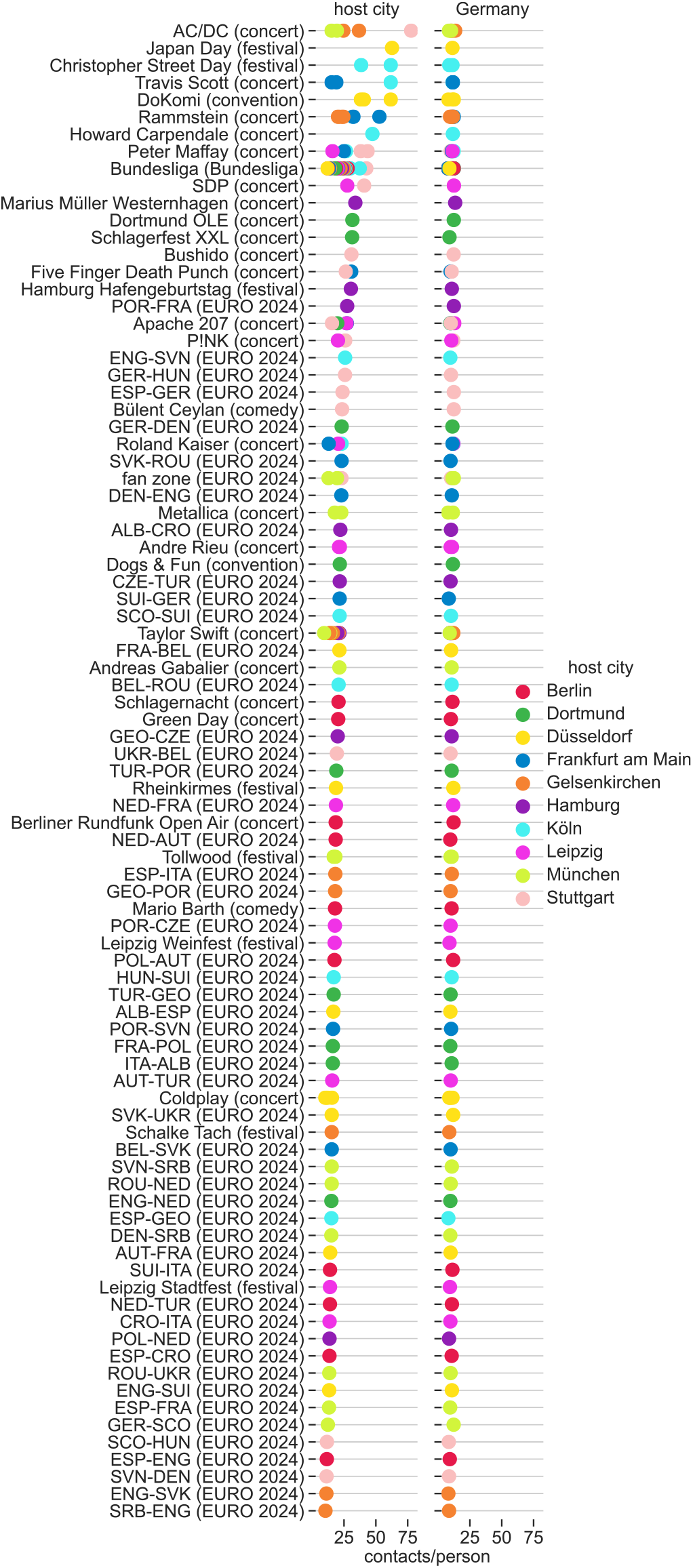
Locally contained and type-dependent contact increases in MGE host cities. Mean number of contacts per person *C/N* (or mean degree in network terminology) for various events across 10 host cities (Tab. S14). These values are computed separately for Germany (right column), within the host city (left column), and within the host venue (shown in the extended version of this figure in Fig. S6) of the MGE. Thus, these values represent the expected contact number for a person located within the host country, within the host city or within the host venue on the day of the MGE. Each dot represents a single occurrence of the specified MGE; multiple dots per MGE are shown when the same event was repeated in different cities and/or on different days. MGEs are sorted by decreasing host-city contacts per person.

Expectedly, contact rates within MGE venues are high (up to 2,000 contacts per person per day; Fig. S6). By contrast, effects from these local peaks are effectively diluted when considering Germany at-large, where exposures fluctuate around 10 contacts per person per day regardless of the MGE (Fig. 2). This confirms that MGE-related contacts are locally confined and do not substantially change contact rates at the national level. On the city-scale, contact rates are up to an order of magnitude higher than at the national level, between 30 and 100 contacts per person per day, with the increase depending on the MGE type (Fig. 2).

Most notably, festivals (such as Japan Day, Christopher Street Day) and some concerts by major artists (Rammstein, AC/DC) lead to higher host city contact rates than UEFA EURO 2024 matches (Fig. 2). UEFA EURO 2024 matches with German participation and/or heavy rainfall tend toward higher contact rates, presumably due to engagement of the local population and confinement of people in smaller, rain-protected spaces (Fig. 2, Tab. S7). Particularly low contact rates were observed for UEFA EURO 2024 matches with violence among fans (Tab. S7). These findings demonstrate a strong dependence of contacts on the type of MGE and special circumstances (e.g. static/closed configuration in football stadiums versus dynamic/open audiences at festivals and during rainfall). Our results suggest a pivotal role of peripheral activity around MGE venues for the importance and type dependence of MGE contacts; contact rates increase because MGE contacts extend within host cities well beyond MGE venues.

### Dominance of out-of-venue contacts

To investigate the effect of MGEs on host cities more closely, we use the time and position information available for all contacts to infer their precise settings, i.e. the type of location where they take place. We apply this spatio-temporal analysis to representative examples from different types of MGEs mentioned above, namely UEFA EURO 2024 (football), Rammstein (concert) and DoKomi (fair). We compare our results with the contact patterns one week prior to the event as a non-event baseline.

First, we consider the temporal distribution of contacts (Methods, SI Sec. S9). For UEFA EURO 2024 matches, counting contacts by hour relative to the start of the match yields Fig. 3. Contact peaks are observed primarily before and, to a lesser extent, after the end of the matches (Fig. 3, Tab. S8). Approximately 30 % of all contacts on the MGE day occur within two hours prior to a match, while the match accounts for less than 10 % of all contacts. A similar dynamics is also observed for concerts, but not for fairs/festivals (Fig. S7). This finding highlights the relevance of dynamic phases in mass gatherings, where moving crowds generate numerous contacts. For comparatively static events, where individuals show only limited movement during the event (football, concerts), these contacts are concentrated before and after the event, while the event itself represents a minimum. Further, this explains the hierarchy of contact exposures between MGEs (Fig. 2) in which festivals and concerts were dominant due to their dynamic nature, i.e. the entire event involves movement of crowds.

**FIG. 3.**
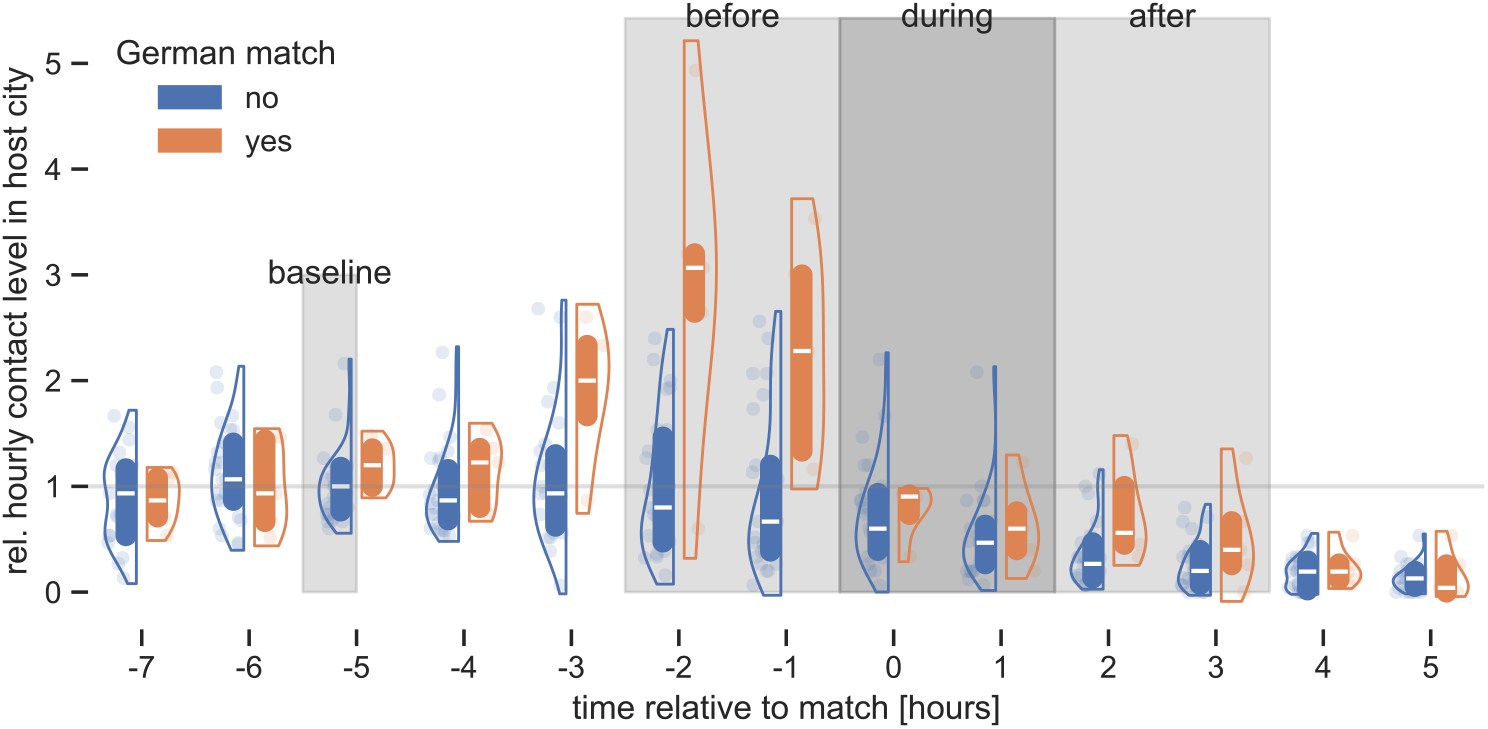
Temporal distribution of contacts in MGE host cities. Relative hourly contacts in host cities of UEFA EURO 2024 matches with German participation (*n* = 5, orange violins, Tab. S5) vs. non-MGE baseline days (*n* = 27, same cities and weekdays, blue violins). Boxes represent center quartiles and white bars inside boxes the median. Data for different matches is aligned relative to the start time of the match (*h* = 0, dark-gray shading). Relative contact levels are defined by city as fold change against *h* = *−*5 on non-MGE days; thus, the median value at *h* = *−*5 on non-MGE days is 1.

Second, we investigate the spatial dimensions of contacts. Using GPS location data and OpenStreetMap, we map contacts to specific settings (Methods, SI Sec. S10, Fig. S8). This allows us to measure contact frequencies in various settings in relation to MGEs (Fig. S9). Using the example of UEFA EURO 2024, we group settings with similar contact dynamics using hierarchical clustering (Methods, SI Sec. S12.1), which leads to three dominant groups of settings with distinct, characteristic contact dynamics, shown in Fig. 4A.

**FIG. 4.**
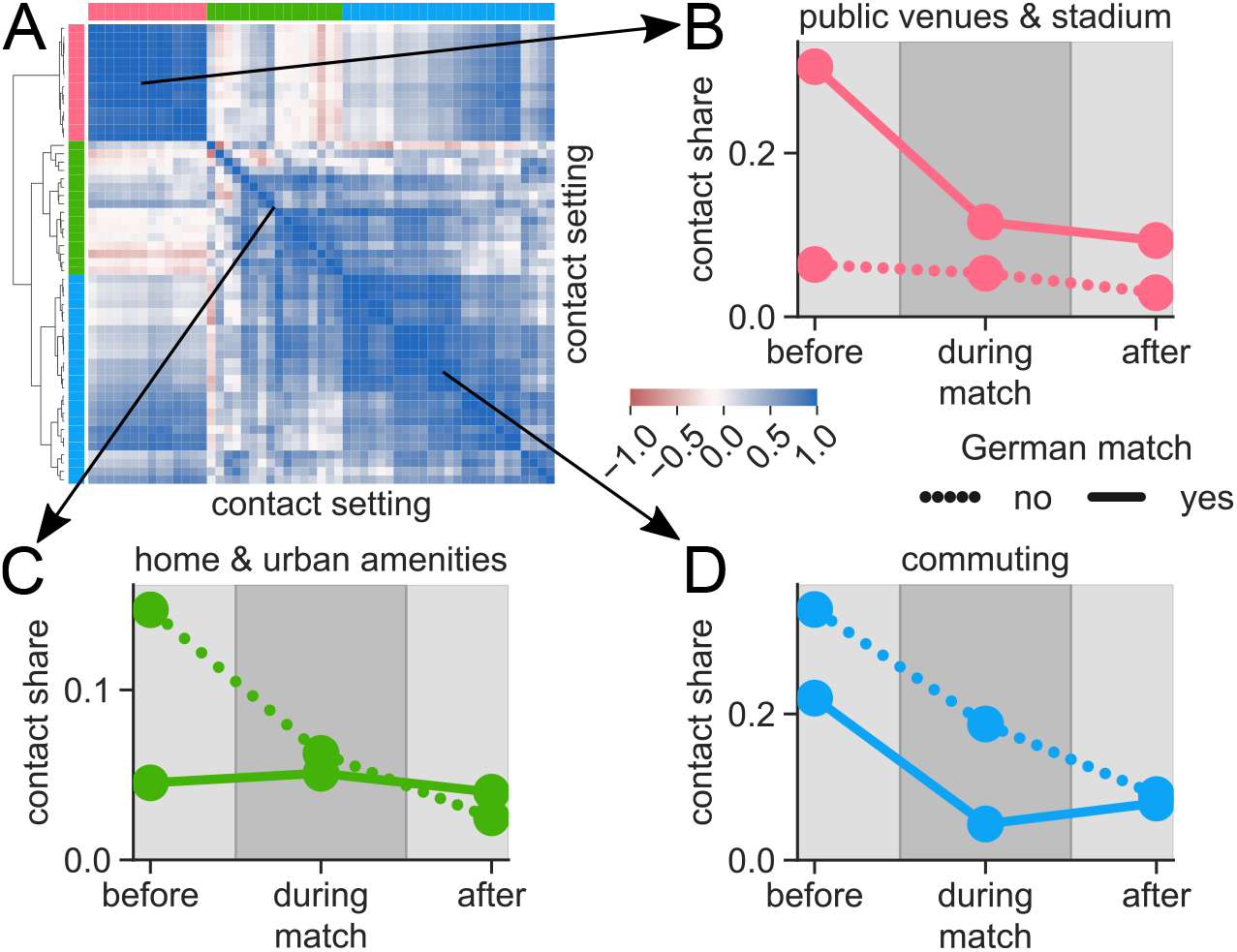
Spatial distribution of contacts in MGE host cities. **(A)** Grouping contact settings with similar contact dynamics in host cities of UEFA EURO 2024 matches with German participation: Each row and column of the correlation matrix corresponds to a single setting (e.g. `leisure:stadium`). The correlation matrix quantifies similarity (+1) or dissimilarity (*−*1) between any pair of settings. Hierarchical clustering (dendrogram) reveals three dominant groups of settings with distinctive contact dynamics (high-lighted in red, green and blue). **(B**,**C**,**D)** Percentage that each group of settings and each time window (2 h before, during and after a match) contributes to all daily contacts in host cities, separately for match days (solid lines) and non-MGE days (dotted lines). For instance, 30 % of contacts on MGE days occur in public venues prior to the event. These three groups are labeled based on the most prominent settings within each group (Tab. S9): **(B)** public venues and stadium, **(C)** home and urban amenities, **(D)** commuting.

Contacts in home, commute and routine settings in host cities are suppressed (Fig. 4C,D), while public venues and host venues (stadium) show increased contacts in particular before matches (Fig. 4B). Settings related to traveling to and from the match also show peaks before and after the matches, but represent a smaller share of overall contacts compared to regular days (Fig. 4D). The MGE venue itself shows the highest number of contacts before the match and lowest after the match, in line with our previously mentioned dynamic and static phases during MGEs. Overall, the share of pre-MGE out-of-venue settings, which concentrate up to 30 % of all contacts in MGE host cities (Fig. 4B), is a surprising feature of MGEs that should be considered in disease modeling and intervention design.

These results demonstrate the pivotal role of peripheral activity around MGE venues for the importance and type dependence of MGE contacts; contact rates increase because MGE contacts extend within host cities well beyond MGE venues. This reveals a substantial fault across the literature on MGEs: MGE studies have been limited to specific venues (e.g. football stadiums or concert halls) [1] or national contact-tracing with coarse resolution [7], which makes host city contacts the blind spot of our current understanding of the role of MGEs.

### Spatial reach and duration of MGE contacts

Since MGEs only locally affect contact rates, we hypothesize other mechanisms, specifically the introduction of long-range contacts in MGEs to be the underlying mechanism leading to widespread spikes in potentially infectious contacts, as demonstrated for the previous UEFA EURO 2020 [7].

Using the home locations of two contact partners (SI Sec. S2), we compute home-to-home distances, corresponding to the distance a virus would travel if transmitted through the contact (Fig. S10C, Methods, SI Sec. S14). Further, we separate contacts into “random” and “recurrent” contacts based on whether or not a given pair of devices had contact on one or multiple days, respectively (Fig. S10A,B, Methods, SI Sec. S13). Comparing contact counts in each class shows that random contacts, which are likely to be underrepresented in other contact datasets [36], are increased on MGE days compared with non-MGE baseline days, while recurrent contacts remain constant (Fig. 5A, Tab. S10, Methods, SI Sec. S13). Fig. 5B plots the distribution of home-to-home distances of contacts, separately for each contact class: Recurrent contacts follow a bimodal distance distribution (home-to-home distances *<* 10 m and *>* 1 km, respectively) and emphasize the role of every-day contact activity in home and other routine settings such as workplaces, regardless of MGEs (Fig. 5B, Tab. S11). Importantly, random contacts during MGEs are commonly long-distance, i.e. co-locations between MGE visitors with home-to-home distances several hundreds of kilometers apart within Germany (Fig. 5B, Tab. S11). This effect is more prominent in the context of low background contact activity, such as in the comparatively small city of Gelsenkirchen which hosted major MGEs such as UEFA EURO 2024 as well as Taylor Swift and Rammstein concerts (Fig. S11A,B, Tabs. S10 and S11). Scatter plots of contact-to-home distances reveal further structure among recurrent contacts (Fig. S11C): They show the existence of different subtypes of recurrent contacts, such as contacts with visitors and contacts between household members away from home, while random contacts appear as an omnipresent feature of both every-day activity and mass gatherings.

**FIG. 5.**
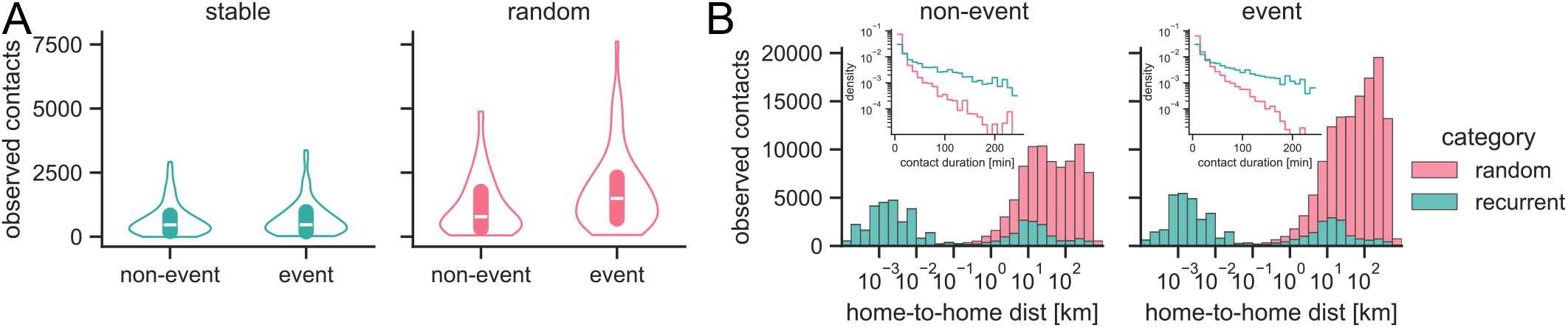
MGEs induce short-lived small-world contacts. **(A)** Distribution of the observed number of recurrent contacts per day (green violin) and random contacts per day (red violin) in host cities for *n* = 67 MGEs (right violins, Tab. S14) and the same number *n* = 67 of non-MGE days 7 days prior to the MGEs as a baseline (left violins). **(B)** Distribution of home-to-home distances and contact durations (inset) for recurrent (green) vs. random (red) contacts on non-MGE (left panel) vs. MGE days (right panel). Home-to-home distances measure how far the virus would travel effectively if the co-location contact had been infectious; note the log scale on the horizontal axis. Distance distributions are shown as stacked histograms, duration distributions as densities. Statistical test results that establish MGE vs. non-MGE differences are provided in Tabs. S10, S11 and S12. Fig. S11A,B extends the results shown in this figure.

Beyond their number and spatial reach, random contacts also change in temporal composition on MGE days: Computing contact duration as the cumulative, not necessarily contiguous time of co-location over a day (Methods, SI Sec. S5) shows that random contacts are more short-lived than recurrent contacts (Fig. 5B), supposedly due to the unplanned nature of random contacts between strangers. However, the duration distribution of random contacts shifts away from the shortest possible contacts (*<* 10 min) toward intermediate durations (10-100 min) typical for MGEs such as football matches and, to a lesser extent, longer (*>* 100 min) durations, both across the 10 host cities combined and in Gelsenkirchen alone (Fig. 5B, Tab. S12). This makes these contacts highly relevant for infectious disease transmission. Recurrent contacts show no comparable shift (Tab. S12).

### Nationwide spatiotemporal reshaping of contacts

We established that increased contact rates are geographically limited to the venue and adjacency of MGE venues (host cities), but that those on-site contacts create long-distance connections relevant to drive infections far from the MGE. However, MGEs with population-wide attention and substantial remote audiences far from the MGE itself may induce additional patterns, e.g. via public viewing events. Even though contact rates at this large geographical scale are dominated by baseline contact activity (Fig. S4), we analyze whether this contact activity shifts toward different settings, e.g. towards pubs as was speculated for the UEFA EURO 2020 tournament [7].

To quantify the spatiotemporal reshaping of contacts at the national scale, we assign OSM settings to all co-location contacts detected across Germany during the data collection period (Methods, SI Sec. S10, Fig. S8). This allows us to quantify the relative contact intensity across settings in Germany on an hourly basis over a period of four months (Figs. S12-S18). Focusing henceforth on the UEFA EURO 2024 quarter-final match between Spain and Germany in Stuttgart on July 5, 2024 (ESP– GER, Tab. S5), we proceed similarly as for the spatiotemporal analysis of host cities (Methods, SI Sec. S12.2) and group settings with similar contact shifts using hierarchical clustering (Fig. 6A). From the most prominent settings within each of the resulting three clusters (Tab. S13), we find that the spatiotemporal contact landscape across Germany decomposes into socializing, shopping, and routine settings. While socializing contacts increase by 22 % compared to an average Friday, shopping contacts decrease by 8 %, and routine contacts remain effectively unchanged, with a 2 % net increase (area under the curve in Fig. 6B). Notably, the nationwide redistribution of contacts across settings is concentrated between two hours before and two hours after the match (Fig. 6B). The marked contact peaks in socializing settings at breakfast and lunch time (Fig. 6B) again emphasize the role of contacts indirectly associated with the MGE. Routine contacts decrease during the match but increase a few hours before and after the match, contributing to a small net increase of 2 % over the day (Fig. 6B). Note that these are percentage changes within each group of settings, so the overall net effect does not need to sum to zero despite the negligible net effect on contact numbers at the national scale (Fig. S4). Aggregated contact heatmaps per group of settings directly show the altered contact activity on July 5, 2024 compared with other days (Fig. S19, SI Sec. S11).

**FIG. 6.**
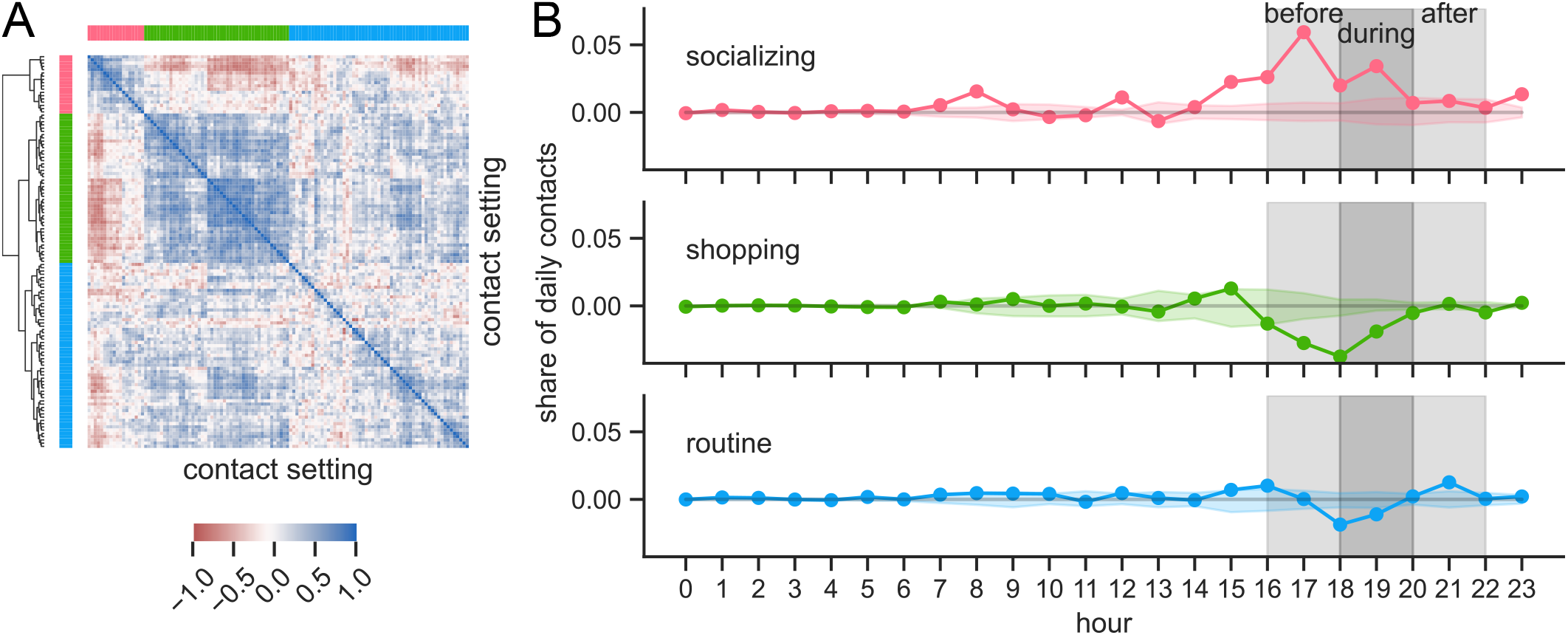
Spatial redistribution of contacts in Germany during major football event. **(A)** Grouping contact settings across Germany with similar hourly contact dynamics on the day of the UEFA EURO 2024 quarter-final ESP–GER on July 5, 2024 in Stuttgart. Hierarchical clustering (dendrogram) on the correlation matrix, similarly to Fig. 4, reveals three dominant groups of contact settings (highlighted in red, green and blue) with distinctive contact dynamics. Based on the most prominent settings within each group (Tab. S13), these groups correspond to “socializing”, “shopping” and “routine” settings. **(B)** Change in contact numbers across Germany in each group of settings at every hour of the day of the match (July 5, 2024), expressed as percentage of all contacts within the same group of settings on a regular non-MGE day. Colored shadings correspond to fluctuations across non-MGE days. The two-hour intervals before, during and after the match are highlighted by gray shading.

## DISCUSSION

In this work, we characterized contacts during MGEs relevant for infectious disease transmission in Germany using large-scale GPS-based mobile phone data covering the time period between April and August 2024. Specifically, we analyzed local and national spatiotemporal contact distributions in relation to various MGEs, and their structural context within the national contact network.

Our analysis reveals four main findings. First, we show strong local contact increases in host cities in connection with MGEs, while any such local net increases are largely diluted on the national scale. Second, while the MGE is typically spatiotemporally confined, contact rate spikes mostly emerge outside the official time and venue of the MGE, particularly in the hours before the event. These contacts are likely to escape from observation by means of established data sources such as surveys or local sensors (i.e. LiDARs or wearables [37]). Third, contact patterns differ substantially by MGE type, with dynamic and open events such as festivals and some concerts producing higher host-city contact rates than football matches. Fourth, MGEs introduce sporadic long-range contacts, i.e. contacts between individuals whose home regions are far apart, thereby creating small-world-like shortcuts in the national contact network. Moreover, these long-range random contacts tend to last longer during MGEs than on non-MGE days, thus compounding their potential to transmit infection across the distances they bridge.

These findings suggest that the epidemiological relevance of MGEs is not determined solely by the total number of contacts generated at the event venue. Instead, MGEs appear to reshape contact networks by concentrating individuals from different regions into shared spaces for short periods of time. This mechanism can explain how locally confined gatherings may contribute to geographically widespread transmission chains even when aggregate national contact numbers remain largely unchanged. In this sense, MGEs may act less as a uniform increase in population-wide contact intensity and more as transient network rewiring events that connect otherwise weakly connected communities.

Our results align with previous findings that out-of-venue activity is the major driver of MGE-related infections and that NPIs targeting out-of-town visitors has measurable effect on reduced incidence in Germany [38] and elsewhere [39, 40], while the event itself has little effect [41, 42]. These insights have practical implications for public health measures during MGEs in times of new emerging threats [43]. Gatherings before the actual events have the potential to become “superspreader events” and could play a critical role in seeding or accelerating the spread of pathogens [44]. Mitigation strategies should therefore consider also the contacts introduced by MGEs before and after the event to effectively reduce the transmission potential.

This study also demonstrates the value of mobile phone location data for infectious disease epidemiology. Compared with surveys, local sensors, wearables, or venue-specific contact-tracing, GPS data provide broad spatial coverage and high temporal resolution across many events and settings. This makes it possible to compare MGEs using a common methodology and to identify contact patterns that would otherwise remain hidden. At the same time, GPS-based co-location data should be viewed as complementary to, rather than a replacement for, other data sources. Their strength lies in detecting population-level exposure patterns across space and time, while other approaches may provide richer information on contact duration, proximity, demography, infection status, or behavior.

Several limitations should be considered, some of which inherent to mobile phone GPS data [45]. First, the dataset represents less than 1 % of the population with regionally varying participation rates, capturing only a fraction of real-world contacts. As a result, this limited sampling poses several potential biases, including under-representation of certain demographic groups and over-detection of contacts during periods of phone activity and inhibit the comparison between different regions. We address part of this issue through a scaling framework, but residual sampling bias is likely.

Second, international visitors as well as minors (below 18 years of age) are likely to be underrepresented or absent from our dataset, restricting our conclusions primarily to the adult local (German) population. This is particularly clear for UEFA EURO 2024 matches and Taylor Swift concerts which attract substantial international and younger audiences. The anonymity of users in our dataset prevents access to their socio-demographic features, preventing us from correcting for these features. As a result, some long-range (international) contacts and the overall contact density for international or young audience events may be underestimated.

Third, mobile phone pings are intermittent, occurring approximately every 15 minutes on average. Only contacts between two panel members whose devices generate pings within the relevant time window can be detected. Consequently, the observed contacts represent only a small fraction of all real-world contacts.

Fourth, GPS precision limits the interpretation of co-location contacts. Our contact definition captures proximity within a spatial threshold of 16 m and a temporal window of 10 minutes, which should be understood as a measure of crowdedness or potential exposure rather than confirmed close-range interpersonal contact. Nevertheless, sensitivity analyses indicate that MGE-specific signals are robust across alternative contact definitions, suggesting that the observed patterns are not driven solely by the chosen threshold.

This study shows how mobile phone data can help us understand how contacts arise during MGEs. While most research focuses on international travel or general crowd management, this work looks at the risks for the local population. We found that additional contact may occur not just at the main event but also in places where people gather before the start and after the end of the event. The type of contacts, not just how many, matters most for respiratory disease spreading. Even with limits like only having data from a biased sample, this method gives useful information to help target public health measures. This can assist public health officials to figure which areas or times are at higher exposure risk and plan better ways to keep people safe during MGEs.

## METHODS

### Data collection

To systematically map contacts across settings of social activity, we build on a panel of about 330,000 anonymous mobile phone users distributed across Germany (ca. 0.4 % of the German population) [18, 31]. Mobile phone users were recruited on an opt-in basis to contribute their individual GPS-based position data and agreed to their usage for research purposes (see Ethics Statement). The dataset consists of frequently generated pings from individual enlisted mobile phones carrying unique and persistent identifiers, timestamps, and GPS-based coordinates, thus informing about the whereabouts of phone users at the time of data generation and their movement over time (Figs. S1 and S2, SI Sec. S1). Over the time period between April 29 and August 4, 2024 (i.e. calendar weeks 19 to 31), we gathered and analyzed around 2.5 billion pings from users across the country. This period covers a wide range of MGEs in Germany, among which most notably the UEFA EURO 2024 football tournament held between June 14 and July 14, 2024, across 10 cities in Germany (Tabs. S1, S5 and S6), as well as regular sport events (e.g. Bundesliga matches), concerts and shows from local and international artists (e.g. Taylor Swift, Rammstein, AC/DC, Peter Maffay, Roland Kaiser, Mario Barth) and festivals and fairs (e.g. Japan Day, DoKomi, Christopher Street Day). A complete list of covered MGEs can be found in Tab. S14. This dataset is exemplarily visualized in Fig. S1 using 5 MGEs of various types as key examples. This dataset thus reunites global reach and local precision over a four-month period, including various MGEs, which represents a unique advantage over other data sources (SI Sec. S7) and allows us to detect close-range co-location contacts, settings and MGE-related effects across Germany.

### Co-location contacts

Using individual position data, we adopt a co-location-based approach to detect contacts possibly relevant for the transmission of infectious disease. A contact is defined as any occurrence in which two devices are co-located within the same place at the same time as per timestamps and coordinates recorded from the mobile devices, modulo some permissible temporal and spatial resolution (Fig. S2, SI Sec. S3). In this study, co-location within a spatial distance of 16 meters and a temporal window of 10 minutes was used to identify contacts. This choice amounts to a trade-off between precision (epidemiological accuracy) and sufficient amount of co-location data (statistical accuracy), as larger distance thresholds lead to more numerous but epidemiologically less relevant co-locations (SI Sec. S4). However, we show from a sensitivity analysis (two-sided Mann-Whitney U test) that the signature of MGEs is stable, regardless of the choice of parameters to define a co-location contact (Tabs. S2 and S3). Furthermore, it is important to note that detecting a contact requires both contact partners to be members of the panel and both mobile phones to generate pings at roughly the same time, which is *per se* unlikely to occur. In consequence, while this procedure entails a large amount of detected contacts, the dataset still represents only a tiny sample of all real contacts.

### Contact rates in MGE host cities

The availability of precise coordinates and timestamps for our co-location contacts on different scales allows us to locate detected co-location contacts on the map and compute contact numbers and rates within specific regions and time frames (SI Secs. S2 and S6). Hence, we could answer the questions: how are contact rates affected by MGEs for individuals within the boundaries of their host venue, host city or host country? Focusing on cities first, we denote by 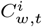 the number of detected contacts within city *i* on weekday *t ∈ {*1, 2, …, 7*}* in week *w* and by 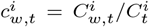 a relative, detrended contact level centered around a baseline value of one. A value of 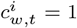 indicates contact rates as expected on a regular day without mass gatherings, while 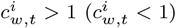 indicates higher (lower) contact rates than usual. The long-term weekday-specific (*t*) and city-dependent (*i*) baseline value is given by 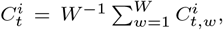, where the sum runs over all *W* weeks between June 1st and August 4th, 2024, and allows us to reduce the effects of public holidays and weekly periodicity of contact rates (Fig. S4). Figs. 1 and S3 show daily contact rates 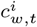 for major cities in Germany.

### Contact rate comparison across MGEs and host cities

To achieve a quantitative comparison of various MGEs, we need to account for the incomplete nature of the contact dataset when measuring contact rates. Mere counts of detected contacts 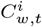 are not sufficient, as they represent a small sample of all real contacts and sampling rates are not uniform across Germany. Here, we use a scaling argument to correct for the sampling biases across different cities in Germany. Contact numbers scale with the size of the studied population *N*, which varies across cities and MGEs, as well as with population shares represented in the user panel *p* and rates of ping generation per user *r*, which both vary across Germany as well (Fig. S5B, Tab. S4). Intuitively, the number of observed contacts increases with population size *N*, the observed population share *p*, and the ping generation rate *r*. This is confirmed empirically via juxtaposition of contact numbers from different cities and MGE of different sizes (Fig. S5B). In consequence, we introduced individualized mean contact rates *C/N* (or “mean degree” in network terminology) which quantifies the mean rate of potentially infectious contacts per person per day associated with a person being located inside the host venue, host city or host country of MGEs. Furthermore, we use a scaling argument to relate the true mean contact rate *C/N* to the equivalent quantity *c/n* measured within the observed sample as a function of *N, p* and *r* (SI Sec. S8 and Eq. (S1) therein).

### Contact settings from reverse geocoding

Stratification of contacts by the settings in which they occur (e.g. home, work, school) is important as it allows to model the relative role of different types of social activity for disease transmission dynamics and their relevance for targeted interventions. In contact surveys such as COVIMOD [10–12, 17], the setting for every recorded contact is directly inquired from the participants. For GPS-based co-location contacts, we infer contact settings from reverse geocoding of the coordinates of any observed co-location event, i.e. determine the map features (setting) at the location of the co-location event (Fig. S8). We performed a mapping of the entire co-location dataset to the map features in which the co-location occurred based on a PostgreSQL database [46] and its PostGIS extension for geospatial data [47]. We utilized an OpenStreetMap (OSM) [48] data extract for Germany downloaded from the OSM online mirror Geofabrik [49], which contains a practically complete set of all buildings, streets, public spaces, businesses, amenities etc. in Germany. **Spatio-temporal contact dynamics**. To study the spatio-temporal distribution of contacts, we group co-location events by their timing relative to the event on hourly basis (before, during or after the event; SI Sec. S9) and by their settings (OpenStreetMap map features found at the location of contact). This allows us to compute setting-specific contact frequency trajectories over the course of MGE and non-MGE days, both for host cities and Germany at-large (Figs. S9 and S12-S18, SI Sec. S11). By contact frequency, we refer to the percentage of all contacts in a day that fall into a designated setting and time window. Different map features may describe settings with similar purpose and contact patterns, e.g. “bench” and “park” or “light rail” and “platform”. Therefore, we group settings by similarity of their contact frequency profiles using hierarchical clustering (using the Pearson correlation matrix between settings, Euclidean metric, and Ward’s linkage; SI Sec. S12).

### Duration and small-worldness of contacts

Not all contacts contribute equally to transmission dynamics. Certain properties such as contact duration [6, 50] and small-worldness [51, 52] can influence disease transmission on a individual and population-wide-scale, respectively. While contact duration determines transmission efficiency per contact, small-worldness implies that disease can be carried over long distances, potentially being introduced into new, yet unaffected communities. To reveal the effects of MGEs on these contact properties, we compare the small-worldness, duration and recurrence of contacts between MGE and non-MGE days in host cities and for Germany at-large.

In fact, the concept of “small-worldness” of interactions, founded in network theory, states that few long-range interactions can have outstanding effects across the entire population and greatly facilitate the spread by creating “shortcuts” for potential transmission chains which would otherwise be subject to the constraints of spatial/geographical distance and community boundaries [53–56]. To study small-worldness of contacts, we compute the home-to-home distances of persons having a contact (Fig. S10C) as the effective “travel distance” of a virus in case of transmission.

The temporal dynamics of contacts can be considered on different timescales: Within the day, we consider the contact duration. A contact can last from minutes to hours, which we captured by weighing contacts by the cumulative time of co-location between the same pair of users within the day in multiples of 10 min (Figs. 5B, S3 and S11B,C, SI Sec. S5). On longer timescales, we consider the concept of contact repetition, i.e. whether or not the same contact repeatedly occurs on different days, which is likely for e.g. household and work contacts but unlikely e.g. between strangers visiting the same MGE. To distinguish incidental one-time encounters and routine contacts, we check for each pair of users, identifiable through their persistent IDs, whether or not contact between them was observed only on a single day or repeatedly on multiple days within the study period (Fig. S10A,B). Accordingly, all pairs and contacts are classified as either “random” contacts (single-day occurrence) or “recurrent” contacts (multiple-day occurrence).

## Data Availability

Scripts and data to reproduce all figures and results in this manuscript have been deposited in public GitHub (https://github.com/st-sch/nc-euro24.git) and Zenodo (https://doi.org/10.5281/zenodo.19726144) repositories.

https://doi.org/10.5281/zenodo.19726144

## ACKNOWLEDGMENT

S.S., A.R.H., A.T., and R.P. were supported by the German Federal Ministry for Economic Affairs and Climate Action (BMWK) via the project DAKI-FWS (grant number 01MK21009A). A.K.J., M.Z., J.S., H.T.P., R.M., V.K.J., A.K., and V.B. were supported by the German Federal Ministry for Education and Research (BMBF) via the project OptimAgent (grant number 031L0299A). Additionally, A.K.J., M.Z., and V.B. were partially supported by “Pandemic non-pharmaceutical interventions to flatten the curve: needs, effectiveness and impact in the global South – the example of Ghana”, primarily funded by the Berlin University Alliance (BUA) as part of the Excellence Strategy of the German Federal and State Governments (grant number 113 MC GlobalHealth). M.Z. and V.B. were also partially supported by the German Federal Ministry of Research, Technology and Space (BMFTR) via the project DREAM-EP (“Data-informed Responsive Epidemic Analysis and Multiscale-Modelling for Epidemic Preparedness”) within the MONID – Phase II – Consortium (grant number 031L0323E). The authors would like to thank Prof. Markus Löffler for helpful discussions. Attribution for maps and map data: © OpenStreetMap contributors.

## ETHICS STATEMENT

Participation in this opt-in study was voluntary, and an informed consent to data collection was obtained from each of the participants. All analyses were carried out on anonymized data. The GPS location data are collected via a software development kit developed for the primary purpose of assessing the quality of cell phone networks. The general terms and conditions of this data collection (www.netcheck.de/datenschutz) also cover the use for research purposes through a broad consent. The Ethics Committee of the Medical Board Westfalen-Lippe and the University of Münster gave ethical approval for this work (reference number 2020-473-f-s). Data collection and usage in this work was additionally reviewed and approved by law firm GvW Graf von Westphalen (gvw. com), deemed compliant with regulations under Federal German Law with regard to protection of privacy and personal information (DSGVO).

## AUTHOR CONTRIBUTIONS

Conceptualization: S.S., A.R.H., A.K.J., R.M., V.K.J., A.K., V.B. Data curation: S.S., A.R.H., A.K.J., M.Z., A.T., R.P. Formal analysis: S.S., A.R.H., A.K.J., M.Z., A.T., H.T.P., C.X. Funding acquisition: S.S., R.M., R.P., V.K.J., A.K., V.B. Investigation: S.S., A.R.H., A.K.J., M.Z., A.T., R.M., R.P., V.K.J., A.K., V.B. Methodology: S.S., A.R.H., A.K.J., M.Z., A.T., V.B. Project administration: S.S., R.M., R.P., V.K.J., A.K., V.B. Software: S.S., A.R.H., A.K.J., M.Z., A.T. Resources: S.S., R.M., R.P., V.K.J., A.K., V.B. Supervision: S.S., R.M., V.K.J., A.K., V.B. Validation: S.S., A.R.H., A.K.J., M.Z., A.T. Visualization: S.S., A.R.H., M.Z., A.T., V.B. Writing – original draft: S.S., A.R.H., A.K.J., M.Z., J.S., V.K.J., A.K., V.B. Writing – review & editing: S.S., A.K.J., A.R.J., M.Z., J.S., A.T., L.F., H.T.P., C.X., R.M., R.P., V.K.J., A.K., V.B.

## COMPETING INTERESTS

S.S., A.R.H., A.T., and R.P. are employees of NET CHECK GmbH. M.Z. is an intern at NET CHECK GmbH. All other authors declare no competing interests.

## Notes

### Author Declarations

Participation in this opt-in study was voluntary, and an informed consent to data collection was obtained from each of the participants. All analyses were carried out on anonymized data. The GPS location data are collected via a software development kit developed for the primary purpose of assessing the quality of cell phone networks. The general terms and conditions of this data collection (www.netcheck.de/datenschutz) also cover the use for research purposes through a broad consent. The Ethics Committee of the Medical Board Westfalen-Lippe and the University of Münster gave ethical approval for this work (reference number 2020-473-f-s). Data collection and usage in this work was additionally reviewed and approved by law firm GvW Graf von Westphalen (gvw.com), deemed compliant with regulations under Federal German Law with regard to protection of privacy and personal information (DSGVO).

## REFERENCES

[1] Stefan Moritz et al. The risk of indoor sports and culture events for the transmission of covid-19. Nature Communications, 12(1), August 2021.

[2] Ziad A Memish et al. Mass gatherings medicine: public health issues arising from mass gathering religious and sporting events. The Lancet, 393(10185):2073–2084, May 2019.

[3] Jehad Feras AlSamhori et al. Implications of the covid-19 pandemic on athletes, sports events, and mass gathering events: Review and recommendations. Sports Medicine and Health Science, 5(3):165–173, September 2023.

[4] European Centre for Disease Prevention and Control. Mass gathering events and communicable diseases: Considerations for public health authorities. Technical report, European Centre for Disease Prevention and Control, Stockholm, 2024.

[5] Amaia Artazcoz Glaria et al. Mass-gathering decision making and its implementation during the covid-19 pandemic. The Lancet Public Health, 8(12):e912–e913, December 2023.

[6] Luca Ferretti et al. Digital measurement of sars-cov-2 transmission risk from 7 million contacts. Nature, 626(7997):145–150, December 2023.

[7] Michelle Kendall et al. Drivers of epidemic dynamics in real time from daily digital covid-19 measurements. Science, 385(6710), August 2024.

[8] Philip Rutten et al. Modelling the dynamic relationship between spread of infection and observed crowd movement patterns at large scale events. Scientific Reports, 12(1), September 2022.

[9] Frederik Verelst et al. Socrates-comix: a platform for timely and open-source contact mixing data during and in between covid-19 surges and interventions in over 20 european countries. BMC Medicine, 19(1), September 2021.

[10] Damilola Victoria Tomori et al. Individual social contact data and population mobility data as early markers of sars-cov-2 transmission dynamics during the first wave in germany—an analysis based on the covimod study. BMC Medicine, 19(1), October 2021.

[11] Jasmin Walde et al. Effect of risk status for severe covid-19 on individual contact behaviour during the sars-cov-2 pandemic in 2020/2021—an analysis based on the german covimod study. BMC Infectious Diseases, 23(1), April 2023.

[12] Huynh Thi Phuong et al. Changes in social contact patterns in germany during the sars-cov-2 pandemic – an analysis based on the covimod study. BMC Infectious Diseases, 25(1), April 2025.

[13] Stephen M. Kissler et al. Sparking “the bbc four pandemic”: Leveraging citizen science and mobile phones to model the spread of disease. November 2018.

[14] Rachael Pung et al. Using high-resolution contact networks to evaluate sars-cov-2 transmission and control in large-scale multi-day events. Nature Communications, 13(1), April 2022.

[15] Piotr Sapiezynski et al. Interaction data from the copenhagen networks study. Scientific Data, 6(1), December 2019.

[16] Forrest W. Crawford et al. Impact of close interpersonal contact on covid-19 incidence: Evidence from 1 year of mobile device data. Science Advances, 8(1), January 2022.

[17] Huynh Thi Phuong et al. Social contact patterns derived from an epidemiological survey and gps-based co-location data – a systematic comparison using parallel data collections during the covid-19 pandemic in germany. Epidemics, 54:100886, March 2026.

[18] Sten Rüdiger et al. Predicting the sars-cov-2 effective reproduction number using bulk contact data from mobile phones. Proceedings of the National Academy of Sciences, 118(31), July 2021.

[19] Sebastian A. Mueller et al. Comparing gps and cell-based mobile phone data to identify activity participation during the covid-19 pandemic. EPJ Data Science, 13(1), November 2024.

[20] Stefania Rubrichi, Zbigniew Smoreda, and Mirco Musolesi. A comparison of spatial-based targeted disease mitigation strategies using mobile phone data. EPJ Data Science, 7(1):17, 2018.

[21] Kyra H. Grantz et al. The use of mobile phone data to inform analysis of covid-19 pandemic epidemiology. Nature Communications, 11(1), September 2020.

[22] Michele Tizzoni et al. On the use of human mobility proxies for modeling epidemics. PLoS Computational Biology, 10(7):e1003716, July 2014.

[23] WHO Novel Coronavirus-19 Mass Gatherings Expert Group. Mass gathering events and reducing further global spread of covid-19: a political and public health dilemma. The Lancet, 395(10230):1096–1099, 2020.

[24] World Health Organization. Risk communication and community engagement readiness and response toolkit: Mass gatherings. World Health Organization, 2025.

[25] Marta C. González et al. Understanding individual human mobility patterns. Nature, 453(7196):779–782, June 2008.

[26] Chaoming Song et al. Limits of predictability in human mobility. Science, 327(5968):1018–1021, February 2010.

[27] Francesco Calabrese et al. Understanding individual mobility patterns from urban sensing data: A mobile phone trace example. Transportation Research Part C: Emerging Technologies, 26:301–313, January 2013.

[28] Caterina Balzotti et al. Understanding human mobility flows from aggregated mobile phone data. IFAC-PapersOnLine, 51(9):25–30, 2018.

[29] Christian M. Schneider et al. Unravelling daily human mobility motifs. Journal of The Royal Society Interface, 10(84):20130246, July 2013.

[30] Zhihua Zhong et al. Human mobility description by physical analogy of electric circuit network based on gps data. Scientific Reports, 14(1), June 2024.

[31] Steven Schulz et al. Real-time dissection and forecast of infection dynamics during a pandemic. March 2023.

[32] Serina Chang et al. Mobility network models of covid-19 explain inequities and inform reopening. Nature, 589(7840):82–87, November 2021.

[33] Dr Alejandra Hidalgo et al. Leveraging real-time population-scale gps data to forecast and identify components of epidemic dynamics. International Journal of Infectious Diseases, 152:107651, March 2025.

[34] Abigail L. Horn et al. Population mobility data provides meaningful indicators of fast food intake and diet-related diseases in diverse populations. npj Digital Medicine, 6(1), November 2023.

[35] Laura Alessandretti. What human mobility data tell us about covid-19 spread. Nature Reviews Physics, 4(1):12–13, December 2021.

[36] Rossana Mastrandrea et al. Contact patterns in a high school: A comparison between data collected using wearable sensors, contact diaries and friendship surveys. PLOS ONE, 10(9):e0136497, September 2015.

[37] Deepak Sharma et al. A review on technological advancements in crowd management. Journal of Ambient Intelligence and Humanized Computing, 9(3):485–495, November 2016.

[38] Kai Fischer. Thinning out spectators: Did football matches contribute to the second covid-19 wave in germany? German Economic Review, 23(4):595–640, April 2022.

[39] Matthew Olczak et al. Mass outdoor events and the spread of an airborne virus: English football and covid-19. SSRN Electronic Journal, 2020.

[40] Kimberly Marsh et al. Contributions of the euro 2020 football championship events to a third wave of sars-cov-2 in scotland, 11 june to 7 july 2021. Eurosurveillance, 26(31), August 2021.

[41] Jonas Dehning et al. Impact of the euro 2020 championship on the spread of covid-19. Nature Communications, 14(1), January 2023.

[42] Helena Heese et al. Results of the enhanced covid-19 surveillance during uefa euro 2020 in germany. Epidemiology and Infection, 150, 2022.

[43] Andrzej Jarynowski et al. Sentiment analysis, topic modelling and social network analysis, page 210–224. Routledge, January 2022.

[44] J. O. Lloyd-Smith et al. Superspreading and the effect of individual variation on disease emergence. Nature, 438(7066):355–359, November 2005.

[45] Francisco Barreras et al. The exciting potential and daunting challenge of using gps human-mobility data for epidemic modeling. Nature Computational Science, 4(6):398–411, June 2024.

[46] The PostgreSQL Global Development Group. PostgreSQL database management system. www.postgresql.org, 2025. Version 17. Accessed: 2025-06-10.

[47] PostGIS Project Steering Committee. PostGIS – spatial and geographic objects for PostgreSQL. postgis.net, 2025. Version 3.5. Accessed: 2025-06-10.

[48] M. Haklay et al. Openstreetmap: User-generated street maps. IEEE Pervasive Computing, 7(4):12–18, October 2008.

[49] Geofabrik GmbH et al. OpenStreetMap data extract: Germany. download.geofabrik.de, 2025. Accessed: 2025-06-10.

[50] Joël Mossong et al. Social contacts and mixing patterns relevant to the spread of infectious diseases. PLoS Medicine, 5(3):e74, March 2008.

[51] Duncan J. Watts et al. Collective dynamics of ‘small-world’ networks. Nature, 393(6684):440–442, June 1998.

[52] M. E. J. Newman et al. Scaling and percolation in the small-world network model. Physical Review E, 60(6):7332–7342, December 1999.

[53] Duygu Balcan et al. Multiscale mobility networks and the spatial spreading of infectious diseases. Proceedings of the National Academy of Sciences, 106(51):21484–21489, December 2009.

[54] Sten Rüdiger et al. Epidemics with mutating infectivity on small-world networks. Scientific Reports, 10(1), April 2020.

[55] Frank Schlosser et al. Covid-19 lockdown induces disease-mitigating structural changes in mobility networks. Proceedings of the National Academy of Sciences, 117(52):32883–32890, December 2020.

[56] Dirk Brockmann et al. The hidden geometry of complex, network-driven contagion phenomena. Science, 342(6164):1337–1342, December 2013.

