## Supplementary Information for "Sporadic long-range contacts in out-of-venue settings dominate mass gathering events in Germany"

(Dated: August 18, 2026)

### S1. DATA COLLECTION

Each of the roughly 2.5 billion data points, or “pings”, carries information about the ID $i$ of the mobile phone by which it was generated, as well as the exact time $t$ and the exact GPS position $(x, y)$ in terms of geographical longitude $x$ and latitude $y$ of the mobile phone during data generation. The ID $i$ is unchanged for all participating mobile phones over the course of the four-month time period covered in this study and data generation is independently triggered on mobile phones on average every 15 minutes, with variations towards more or less frequent data at times of higher or lower activity of phone users on their mobile devices. This data collection provides us with a discretized trajectory $\{x_i(t_k), y_i(t_k)\}_{k=1,2,...}$ for each mobile phone $i$ over this time period, as schematized in Fig. S2.

### S2. HOME LOCATIONS

We require knowledge about home locations of mobile phone users for multiple occasions: First, assigning phone users to their places of usual stay is required to determine local biases in population sampling and ping generation rates. Second, identifying contacts as home contacts and estimating the small-worldness of non-home contacts requires geographical distances between locations of contact and locations of stay. Since home locations are not directly available from the phone users, we adopt a method based on spatial data clustering to estimate this information [1, 2]: In short, it assumes that a substantial amount of pings is generated at or near an individual’s home location. All pings from a given individual over the study period are therefore projected on the spatial dimensions only (i.e. dropping the timestamps). Spatial clusters of pings are then identified using Density-Based Spatial Clustering of Applications with Noise (DBSCAN) and the weighted centroid coordinates of pings within the most prominent cluster, where the phone user spends the most cumulative dwell time, is taken as the individual’s home location.

### S3. CO-LOCATION CONTACTS

To determine co-location contacts, exact GPS positions $(x, y)$ are mapped onto a static, pre-defined regular square grid with a cell size of 16 m (more precisely: 15.625 m) covering all of Germany (using the `EPSG:32632` local coordinate system). The timestamps $t$ are binned into time windows of length 10 min, of which there are 144 within a day (1,440 min). Using these discretized spatio-temporal coordinates for the whereabouts of mobile devices, we identify co-locations by matching pings with identical coordinates (i.e. same grid cell and same time window) generated from different mobile phones: $x_i = x_j$, $y_i = y_j$ and $t_i = t_j$ for two distinct mobile phones $i$ and $j$ (Fig. S2). Co-locations may involve more than two devices whenever coordinates are shared between more than two devices. From these co-locations, a contact is established between any two mobile phones found within the co-location event. This means, when $n \geq 2$ mobile phones are involved in the co-location, we count $n(n-1)$ contacts, which corresponds to twice the amount of pairs within a group of $n$ individuals (the factor $1/2$ is dropped in accordance with the handshake lemma which states that a contact is counted once for each individual participating in the contact).

### S4. SPATIO-TEMPORAL RESOLUTION

To study the effect of the spatial and temporal granularity, we repeat this procedure for alternative choices of the grid cell size and the time window size: 8 m (7.8125 m), 16 m (15.625 m), 31 m (31.25 m) and 62 m (62.5 m) as well as 10 min and 60 min. Higher resolution (e.g. smaller grid cell size and smaller time windows) leads to fewer, but epidemiologically more relevant contacts. The spatial grids are obtained by starting off with a grid of cell size 1,000 m and then iteratively subdividing cells into 4 equally sized cells at each iteration, thus leading to grids of cell sizes 500 m, 250 m, etc., down to 7.8125 m. This ensures that contacts at higher resolution are strictly contained within the contacts at coarser resolution. The offset of the spatial grid – which reflects the translational invariance of positioning the grid onto the map of Germany – is randomly chosen and expected to play a negligible role.

### S5. DURATION OF CONTACTS

Our definition of contact is based on co-location within fixed 10 min time windows, of which there are 144 within a day (the length of one day corresponds to 1,440 min). We define the contact duration for a device pair as the number of time windows within a day (out of 144) in which they were found to be co-located, regardless of whether the contact arises from a single contiguous or multiple discontinuous co-locations. Accordingly, the contact duration is therefore a multiple of 10 min: 10 min, 20 min, etc. up to the maximal value of 1,440 min. Note that contact duration is defined based on the time of co-location within the day while contact recurrence is defined based on repeated co-location on different days.

### S6. COUNTING CONTACTS

Each co-location contact (involving two individuals) carries information about the exact location and time of the contact and the anonymous IDs of both contact partners. We can thus count contacts that fall within temporal and spatial delimitations of interest, e.g. contacts within a given day, hour as well as within a given city, venue, contact setting, or Germany at-large. Additionally, we can choose to count contacts involving the same two individuals once ("unique contacts") or as many times as it recurs within a given day ("weighted contacts"). The latter choice amounts to weighing contacts by their effective duration, which is known to play a role for the infectiousness of contacts. Long-duration contacts typically occur between household members, co-workers and leisure/travel companions (Fig. S11C). Contact rates are computed based on unique contacts (Figs. 1 and 2), while spatio-temporal contact distributions (Figs. 3, 4 and 6) count every contact regardless of the involved contact partners. Daily contact counts across all 10 cities considered here, namely the host cities of UEFA EURO 2024, are shown in Fig. S3. 169 MGEs are identified from this data in these cities within the study period and are summarized in Tab. S14. Counting unique vs. weighted contacts does not substantially alter the contact footprint of MGEs (Fig. S3).

### S7. DATA QUALITY AND COMPARISON TO OTHER DATASETS

For the purpose of studying the role of MGEs in contact dynamics, crowd-based GPS location data as used in this study has multiple advantages over established methodology: First, contact surveys [3–6] require active input from participants and therefore suffer from fatigue, awareness and memory biases [7]. Also, in spite of their representativeness, surveys are hardly scalable to cohorts large enough to cover MGE audiences within the population at-large. Second, contact-tracing approaches in experimental settings (e.g. concerts, cruises, university campus) [8–11] are strictly limited to contacts within the venue and time of the MGE and thus neglect any dynamics outside the strict perimeter of the MGE. At the opposite extreme, mobile phone-based contact-tracing rolled out to full populations during the SARS-CoV-2 pandemic, using e.g. Bluetooth [12–14] or call records (CDR) [15, 16], builds on relative and approximate positions between exposed individuals and thus lack context information about contacts (precise contact settings, structural contact network context). By contrast, the GPS-derived contact dataset in this study combines the scalability of contact-tracing with the possibility to passively inquire contextual information about contacts (settings via precise GPS coordinates, contact network via persistent individual identifiers), thus allowing to study contact dynamics both inside and outside of MGEs.

### S8. SCALING OF CONTACT RATES ACROSS CITIES AND MGES

The contacts reflected in our dataset represent a small sample of all real contacts, depending on two sampling parameters $p$ (population sampling) and $r$ (temporal sampling), which in turn can vary across different regions or MGE types in Germany. In order to be able to compare contact rates across various MGEs, scaling observed contact rates by these sampling parameters $p$ and $r$ is required. The population share $p = n/N$ denotes the fraction of the full population (for cities) or audience (for MGEs) that participates in the data panel within a particular area of interest. Here, $N$ is the population size or audience size and $n$ the number of data panel participants among the population of $N$ (for cities: Tab. S4). For MGE venues, we use for $N$ the actual audience size reported from the news or the capacity of the venue, assuming actual audiences are close to capacity (Tabs. S1 and S14), and for $n$ the number of unique phone users who generated pings within the MGE venue on the day of the MGE. The ping generation rate per phone user is defined as $r \sim s/n$ where $s$ is the number of pings generated by the $n$ participants over a certain time period (for cities: Tab. S4).

How the number of contacts depends on the number and sampling of participants is a widely studied problem in network theory. From a sampling perspective (Horvitz-Thompson network sampling theory) [17–19] in which an original contact network with $N$ participants and $C$ contacts is thinned by retaining them with probabilities $p$ and $q$, respectively, the contact number per individual (mean degree) scales as $c/n = C/N \cdot pq$. From a network construction viewpoint in contrast, Random Geometric Graphs (RGG) [20, 21] are a model for spatially embedded network growth (such as real-world contact networks) which gives rise to a scaling of the form $c/n = C/N \cdot p^{\beta-1}q$ with the exponent $1 \leq \beta \leq 2$ modulating the strength of spatial constraint on contacts. It mediates between the extreme cases of nearest-neighbor contacts only ($\beta = 1$; Fig. S5A upper panel) and all-to-all mixing/mean-field contacts ($\beta = 2$; Fig. S5A lower panel). These represent two extreme cases of mass-action dynamics typically found together in MGEs: lattice-like contacts during static phases vs. small-world contacts during dynamic phases [22]. The contact detection probability $q$ itself depends on the ping generation rate $r$, i.e. $q \sim r^\gamma$. An exponent $\gamma = 2$ ($\gamma = 1$) is expected under the assumption of independent (correlated) user activity and ping generation on mobile phones. The true individual mean contact rate $C/N$ of MGEs can thus be estimated from knowledge of all other parameters ($c/n$, $p$ and $r$) using the overall scaling law

$$C/N = c/n \cdot p^{1-\beta} r^{-\gamma}. \tag{S1}$$

The strength of spatial constraint and correlation between mobile phone activity is likely to be different at host city scale compared with host venues alone. Therefore, we determine the missing scaling exponents $\beta$ and $\gamma$ separately for cities and for MGE venues. In both cases, we exploit large variations of $p$, $r$ and $c$ across cities or MGEs to find the best fit to the growth scaling law $c \sim p^\beta r^\gamma$ using linear regression on log-transformed data (Fig. S5B,C). We find $\beta = 1.009$ $(0.970; 1.049)$ for host cities and $\beta = 1.447$ $(1.286; 1.607)$ for host venues, where the values in brackets indicate 95 % confidence intervals. This provides empirical evidence for the relevance of geographical constraints ($\beta < 2$) and validity of the RGG scaling. It also demonstrates that contact dynamics within host venues are closer to the all-to-all mixing scenario, while host cities are indistinguishable from the nearest-neighbor contact scenario. Furthermore, we estimate $\gamma = 2.225$ $(1.886; 2.564)$ for host cities and $\gamma = 1.084$ $(-0.493; 2.662)$ for host venues, thus indicating highly correlated mobile phone activity among MGE attendees and essentially uncorrelated city-wide activity, which is expected. Extending the result for host cities to Germany at-large, we assume $\beta = 1$ and $\gamma = 2$ for the scaling of nationwide contact rates.

### S9. TEMPORAL DISTRIBUTION OF MGE CONTACTS

Precise timestamps of co-location contacts enable us to study at what times MGE-induced contacts occur in relation to the MGE itself. Given the dependence on MGE type, we compute time distributions separately for sporting events, concerts and festivals/fairs. From each MGE type, we choose a representative MGE with multiple occurrences on different days and/or in different cities to maximize statistics: UEFA EURO 2024 matches with participation of the German national team (5 occurrences: 2 in Stuttgart, 1 each in Munich, Frankfurt and Dortmund; Fig. 3), Rammstein (8 occurrences: 5 in Gelsenkirchen, 3 in Frankfurt; Fig. S7A) and the DoKomi fair (3 occurrences in Düsseldorf; Fig. S7B), see Tab. S14. Since each occurrence of the same event can be held at different hours of the day (Tab. S5), we introduce a relative time axis centered around the start of the MGE, which defines $h = 0$. We count hourly contacts $C^i_{w,t,h}$, $h = \ldots, -2, -1, 0, 1, 2, \ldots$ for each occurrence $i, w, t$ of the MGEs, both on MGE days and on non-MGE baseline days. As baseline data for each MGE, we choose the contacts on the same weekday in other weeks, as long as they do not have other MGEs or are public holidays, including only the host cities of these MGEs. To account for the different sizes of host cities before superposing data from different occurrences of the same MGE, we normalize contact numbers by their median value at $h = -5$ (for UEFA EURO 2024 matches and Rammstein concert) or $h = 4$ (for DoKomi fair) on non-MGE baseline days.

### S10. IDENTIFICATION OF CONTACT SETTINGS USING OPENSTREETMAP

We use OpenStreetMap (OSM) data to map co-location contacts to contact settings using their precise GPS coordinates. Each co-location contact can map to one or several map features based on spatial overlap, as demonstrated in Fig. S8. Map features in OSM are tagged by one or several key-value pairs describing the purpose or nature of the location, such as `amenity:parking`, `leisure:stadium`, `railway:platform` and `building:retail` (Fig. S8) [23]. Consequently, we define the contact setting of co-location contacts as the key-value pair(s) of OSM map feature(s) they are mapped to. For instance, a contact located in a parking lot per its GPS coordinates would thus be associated with the setting `amenity:parking` (Fig. S8B). Importantly, this definition of contact setting differs from established terminology in that contacts are not necessarily uniquely assigned to a single setting and that different settings are not disjoint: A given co-location contact can be mapped to multiple settings, notably when multiple map features overlap at the location of contact (e.g. `leisure:stadium` and `building:grandstand`; Fig. S8A) or multiple map features fall within the spatial precision of 16 m around the location of contact (e.g. `railway:platform` and `railway:light_rail`; Fig. S8C).

The OSM map data consists of different types of map features: polygons as 2D map features (e.g. buildings), lines as 1D map features (e.g. streets) and nodes as 0D map features (e.g. named entities such as restaurants). The procedure to match OSM map features with contacts, which we represent as 2D squares with dimensions 16 m-by-16 m on the map, depends on the dimension: (i) For 2D polygon-type and 1D line-type map features, a contact is matched based on non-empty intersection between the 2D contact and the map feature (Fig. S8). (ii) 0D node-type map features are first mapped to 2D map features which contain them, e.g. shops inside a mall. Contacts matched with these encompassing 2D map features are also matched with the contained 0D map features, e.g. the contact is matched with both the mall and the shops inside the mall (Fig. S8D). In addition to the OSM map features, selected map features in UEFA EURO 2024 host cities have been additionally tagged as fan zones (`fanzone:yes`; Tab. S1) and on an individual basis as home (`home:yes`) if the location of contact coincided with the home location of at least one of the contact partners within 100 m distance.

The one-to-many nature of this OSM-based contact setting inference has implications for contact counts: When counting contacts by setting, such as in Figs. 4 and 6, co-location contacts are possibly counted more than once, once for each setting to which they had been mapped.

### S11. CONTACT HEATMAPS

After matching co-location contacts with contact settings, we can group the entire co-location dataset by setting, day and hour of the contacts. This allows us to count contacts $C^i_{w,t,h}$ per setting $i$, day $w, t$ and hour $h$ within the study period. Normalized contact counts $c^i_{w,t,h}$ can be conveniently represented as heatmaps (Figs. S12-S18) to reveal setting-specific contact dynamics that match general intuition about everyday life: weekly periodicity, high contacts in leisure settings on weekends, absence of contacts in shopping settings on Sundays and public holidays, evening/night contacts in nightlife venues, etc.

### S12. IDENTIFYING SETTINGS WITH SIMILAR CONTACT DYNAMICS

Contact settings are expected to show characteristic contact dynamics in everyday life and under the influence of MGEs. Different contact settings, e.g. settings with similar purpose such as railway and platform or stadium and grandstand, may show similar contact dynamics and MGE-induced variations. Here, we group contact settings together based on similarity of contact dynamics in the presence and absence of MGEs, i.e. in such a way that settings within a group have similar contact patterns while settings in different groups have distinct contact patterns. A small number of groups of settings defines a higher-level classification of contact settings based on contact patterns under the influence of MGEs.

To achieve this, we perform hierarchical clustering on a standard Pearson correlation matrix $\mathrm{corr}_{i,j} = \frac{1}{H}\sum_h c^i_h c^j_h$ computed from a setting-specific ($i$) and daytime-dependent ($h$) contact signal $c^i_h$ which quantifies the similarity of contact dynamics between any two contact settings $i$ and $j$ (Figs. 4A and 6A). We apply a standard scaler prior to correlation and clustering in order to normalize hourly contact counts $c^i_h$ independently for each contact setting $i$. This ensures that the input data for the clustering analysis has mean $\mu = \langle c^i_h \rangle_h = 0$ and variance $\sigma^2 = \langle c^{i2}_h \rangle_h - \langle c^i_h \rangle^2_h = 1$ for each setting $i$ and captures only the shape (and not the scale) of contact dynamics. The precise contact signals $c^i_h$ used for host cities and Germany at-large is different because nationwide contact variations are dominated by weekday-dependent patterns and require subtraction of baseline contacts prior to the clustering analysis (see below).

Visual inspection of the correlation matrices and dendrograms of the hierarchical clustering reveal the existence of three prominent groups of contact settings for both host cities and Germany at-large (Figs. 4A and 6A). In each case, we identify the settings which fall into each of the groups (Tabs. S9 and S13) and assign higher-level designations based on the most prominent settings within each group: In the Germany-wide analysis, we find that MGE-induced contact dynamics delineate settings associated with socializing, shopping and routine activities (Fig. 6B, Tab. S13). At host city level for UEFA EURO 2024 matches, these groups correspond to stadiums, home and urban amenities and commuting settings (Fig. 4B, Tab. S9).

#### S12.1. MGE host cities

In the case of host cities of UEFA EURO 2024 matches with German participation, we group contacts by setting $i$ and time bin $h$ (within two hours before the start of the match, during the match or within two hours after the match), separately for match days and baseline days. From the contact counts $C^i_{w,t,h}$, we define

$c_h^i = \langle C^i_{w,t,h} \rangle_{w,t} / C_{w,t}$ as the share of all daily contacts associated with setting $i$ and time bin $h$ (Fig. S9). Here, $C_{w,t} = \sum_{i,h} \langle C^i_{w,t,h} \rangle_{w,t}$ represents the average daily contact count. We compute $c_h^i$ separately for match days and baseline days and use it as input for hierarchical clustering of contact settings $i$ as described above. For each setting $i$, $c_h^i$ defines vectors of length 6, hence $H = 6$ in the correlation formula above (three time bins by two categories: match days vs. baseline days).

After clustering of contact settings, we compute contact shares per time bin and group of settings as $\sum_{i \in \text{cluster}} c_h^i$ (Fig. 4B-D).

#### S12.2. Germany at-large

For the Germany-wide analysis of the UEFA EURO 2024 quarter final ESP–GER, we use a different approach because MGE-related contact variations are essentially masked by weekday-dependent contact variations (Fig. S4). First, we define weekday-dependent ($t$) and daytime-dependent ($h$) baseline contacts $C^i_{t,h} = \frac{1}{W} \sum_{w=1}^{W} C^i_{w,t,h}$ for each setting $i$ by averaging over all contact counts after June 10, 2024 and excluding the targeted MGE day (July 5, 2024). Data before June 10, 2024 is excluded from the baseline due to the special patterns from various public holidays which do not follow weekday-specific patterns (Fig. S19). We then use hourly deviations from the baseline on the targeted MGE day ($w'$, $t'$), $c_h^i = C^i_{w',t',h} - C^i_{t',h}$, which capture setting- and daytime-dependent MGE effects, as input for the clustering step. This implies $H = 24$ for this case in correlation formula above. We only include contact settings $i$ with sufficient contact counts in this clustering analysis, specifically, settings with at least 3,000 contacts over the study period (Figs. S12-S18). Further, we restrict the Germany-wide analysis to include only settings represented as nodes in OSM (shops, food places, public spaces, etc.).

To quantify hourly MGE-related contact variations in different higher-level settings after the clustering step, we sum all contact variations within groups of settings and relate this total difference to the total baseline contacts, $\sum_{i \in \text{cluster}} c_h^i / \sum_{h, i \in \text{cluster}} C^i_{t',h}$. This quantifies by how much each group of settings increases or decreases daily baseline contacts at each hour of the MGE day (in terms of percentage change compared to daily baseline contacts; Fig. 6B). Further summing over the hours of the MGE day, $\sum_{h, i \in \text{cluster}} c_h^i / \sum_{h, i \in \text{cluster}} C^i_{t',h}$, then quantifies the MGE-related net contact increase or decrease in each higher-level setting over the MGE day (area under the curve in Fig. 6B). We thus find that socializing contacts across Germany were 22 % increased, shopping contacts decreased by 8 % and routine contacts overall unchanged (2 % net increase) while being redistributed across the hours before, during and after the match.

### S13. RECURRENCE OF CONTACTS

Invariable device IDs in the dataset allow us to follow the contact of any given pair of devices over time, i.e. whether or not the same contact is repeated at different times and on different days (Fig. S10A). Recurrent contacts between e.g. household members and coworkers would manifest in the data by the recurrence of co-locations on different days involving the same pair of device IDs. By contrast, one-time encounters e.g. between strangers in the MGE audience would correspond to co-locations involving device ID pairs which are otherwise completely absent from the co-location dataset. To classify device ID pairs $ij$ as "recurrent" or "random" contacts, we compute for each pair a binary timeseries $b_{ij,t}$ (of length 96 because the data covers a period of 96 days; Fig. S10B). These timeseries indicate for each day $t$ whether or not there was a co-location contact between the two devices, $b_{ij,t} = 1$ if there was a contact (regardless of duration or repeat within the day) and $b_{ij,t} = 0$ if not. We classify a device ID pair $ij$ as "random" if the timeseries was active only on a single day ($\sum_t b_{ij,t} = 1$) or as "recurrent" if it was active on multiple days ($\sum_t b_{ij,t} > 1$), regardless of the contact location. This is a simplification that could tend to misclassify some recurrent contacts as random contacts because of the temporal sampling in the data collection (Figs. S1 and S2), which makes the majority of true co-locations invisible to us.

To reveal MGE-related trends in recurrent and random contacts, we count contacts in each category in host cities for 67 MGEs (Tab. S14) versus the same number of baseline contact counts, which are taken 7 days prior to each of the MGEs to eliminate weekday-dependent effects. In most cases (80 %), these baseline days were indeed non-MGE days.

### S14. SMALL-WORLDNESS OF CONTACTS

While respiratory disease transmissions are the result of close-range contacts, a virus can effectively travel over long distances, depending on the individuals involved in the contact: While transmission between household members effectively keep the virus in place, infections between MGE visitors who live hundreds of kilometers apart can carry the virus over long distances and seed infection clusters in previously unaffected regions and communities. This defines another dimension of contacts beyond mere numbers, duration and spatio-temporal distribution, namely the so-called small-worldness, which is expected to be a critical driver of epidemic spread.

To characterize the small-worldness of contacts, we introduce the home-to-home distance of contacts. It quantifies the effective travel distance of a virus if a given contact had been infectious. From the coordinates of home locations available for most mobile phones, we compute the home-to-home distance between any pair of mobile phones involved in co-location contacts as the 2D planar distance between the two home locations, regardless of the location of the contact (Fig. S10C). Additionally, we compute contact-to-home distances using contact locations and home locations to measure how far from home the contact occurred for each of the involved individuals (Fig. S10C). This provides additional information about possible infection paths of recurrent and random contacts. For instance, recurrent contacts can occur between household members away from home or between household members and visitors inside the household. Also, random contacts typically occur far from the home locations of all involved individuals, e.g. at or nearby MGE venues.

### REFERENCES


[1] Christian M. Schneider et al. Unravelling daily human mobility motifs. *Journal of The Royal Society Interface*, 10(84):20130246, July 2013.

[2] Z. Tao. *Hurricane Evacuation Analysis based on Smartphone Location Data: A Case Study of Hurricane Florence*. phdthesis, 2021.

[3] Frederik Verelst et al. Socrates-comix: a platform for timely and open-source contact mixing data during and in between covid-19 surges and interventions in over 20 european countries. *BMC Medicine*, 19(1), September 2021.

[4] Damilola Victoria Tomori et al. Individual social contact data and population mobility data as early markers of sars-cov-2 transmission dynamics during the first wave in germany—an analysis based on the covimod study. *BMC Medicine*, 19(1), October 2021.

[5] Jasmin Walde et al. Effect of risk status for severe covid-19 on individual contact behaviour during the sars-cov-2 pandemic in 2020/2021—an analysis based on the german covimod study. *BMC Infectious Diseases*, 23(1), April 2023.

[6] Huynh Thi Phuong et al. Changes in social contact patterns in germany during the sars-cov-2 pandemic – an analysis based on the covimod study. *BMC Infectious Diseases*, 25(1), April 2025.

[7] Timo Smieszek et al. Contact diaries versus wearable proximity sensors in measuring contact patterns at a conference: method comparison and participants' attitudes. *BMC Infectious Diseases*, 16(1), July 2016.

[8] Stefan Moritz et al. The risk of indoor sports and culture events for the transmission of covid-19. *Nature Communications*, 12(1), August 2021.

[9] Stephen M. Kissler et al. Sparking "the bbc four pandemic": Leveraging citizen science and mobile phones to model the spread of disease. November 2018.

[10] Rachael Pung et al. Using high-resolution contact networks to evaluate sars-cov-2 transmission and control in large-scale multi-day events. *Nature Communications*, 13(1), April 2022.

[11] Piotr Sapiezynski et al. Interaction data from the copenhagen networks study. *Scientific Data*, 6(1), December 2019.

[12] Michelle Kendall et al. Drivers of epidemic dynamics in real time from daily digital covid-19 measurements. *Science*, 385(6710), August 2024.

[13] Luca Ferretti et al. Digital measurement of sars-cov-2 transmission risk from 7 million contacts. *Nature*, 626(7997):145–150, December 2023.

[14] Forrest W. Crawford et al. Impact of close interpersonal contact on covid-19 incidence: Evidence from 1 year of mobile device data. *Science Advances*, 8(1), January 2022.

[15] Michele Tizzoni et al. On the use of human mobility proxies for modeling epidemics. *PLoS Computational Biology*, 10(7):e1003716, July 2014.

[16] Sebastian A. Mueller et al. Comparing gps and cell-based mobile phone data to identify activity participation during the covid-19 pandemic. *EPJ Data Science*, 13(1), November 2024.

[17] Sten Rüdiger et al. Predicting the sars-cov-2 effective reproduction number using bulk contact data from mobile phones. *Proceedings of the National Academy of Sciences*, 118(31), July 2021.

[18] Steven Schulz et al. Real-time dissection and forecast of infection dynamics during a pandemic. March 2023.

[19] Eric D. Kolaczyk. *Statistical Analysis of Network Data: Methods and Models*. Springer New York, 2009.

[20] Jesper Dall et al. Random geometric graphs. *Physical Review E*, 66(1), July 2002.

[21] Mathew Penrose. *Random Geometric Graphs*. Oxford University Press, May 2003.

[22] Philip Rutten et al. Modelling the dynamic relationship between spread of infection and observed crowd movement patterns at large scale events. *Scientific Reports*, 12(1), September 2022.

[23] OpenStreetMap contributors. Map features – OpenStreetMap wiki. wiki.openstreetmap.org, 2025. Accessed: 2025-06-10.

[24] Wikipedia contributors. Fußball-europameisterschaft 2024. de.wikipedia.org, 2024. Wikipedia, The Free Encyclopedia.

[25] Berlin.de. Fanmeile am brandenburger tor. Berlin.de – Das offizielle Hauptstadtportal, 2024.

[26] Stadt Dortmund. Euro 2024 festival & public viewing. dortmund.de, 2024.

[27] Polizei Düsseldorf. Fan zones und public viewing. Polizei Düsseldorf, 2024.

[28] Tourismus- und Congress GmbH Frankfurt am Main. UEFA EURO 2024 Fan Zone Mainufer. visitfrankfurt.travel, 2024.

[29] Stadt Gelsenkirchen. Festival & events – Fan Zone & public viewing. UEFA EURO 2024 Host City Gelsenkirchen, 2024.

[30] Hamburg Tourismus GmbH. Fan zone hamburg. hamburg-tourism.de, 2024.

[31] koeln.de. Em 2024 in köln: Public viewing. koeln.de, 2024.

[32] RB Leipzig. Leipzig fan zone: Public viewing & events zur UEFA europameisterschaft 2024. rbleipzig.com, 2024.

[33] Referat für Bildung und Sport, Landeshauptstadt München. Fan zone im olympiapark zur UEFA EURO 2024 in München mit public viewing. muenchen.de – Das offizielle Stadtportal, 2024.

[34] Landeshauptstadt Stuttgart. Host city stuttgart – UEFA EURO 2024. uefaeuro2024.stuttgart.de, 2024.

[35] Wikipedia contributors. Spielplan der em 2024: Wann sie urlaub nehmen sollten. www.fr.de, 2024. Frankfurter

Rundschau.
[36] Wikipedia contributors. List of FIFA country codes. Wikipedia, The Free Encyclopedia, 2024.
[37] Die Welt. Deutschland–Dänemark: Diese bilder gingen um die welt. welt.de, 2024.
[38] Sportschau. Regenfälle in Dortmund vor EM-halbfinale. sportschau.de, 2024.
[39] Die Welt. Türkei–Georgien: Das bislang größte spektakel der EM 2024. welt.de, 2024.
[40] YouTube. AUT–TUR heavy rainfall. youtube.com, 2024.
[41] Hessenschau. EM-fanmeile in Frankfurt wegen unwetter geschlossen. hessenschau.de, 2024.
[42] Die Welt. EM 2024: Fans von England und Serbien geraten aneinander. welt.de, 2024.
[43] Hessenschau. Dänemark–England: Römerberg in Frankfurt in britischer hand. hessenschau.de, 2024.
[44] WDR. EM-fanmärsche in Düsseldorf: Belgien und Frankreich. wdr.de, 2024.
[45] RP Online. Datenrekord bei Spanien gegen Albanien in der Düsseldorfer Arena. rp-online.de, 2024.
[46] Sportschau. Nach messerangriff in EM-fanzone in Stuttgart: Tatverdächtiger in U-haft. sportschau.de, 2024.
[47] Waldbühne Berlin. Mario barth – Waldbühne Berlin. waldbuehne-berlin.de, 2024.
[48] Rolling Stone Deutschland. Green day live in Berlin. rollingstone.de, 2024.
[49] mix1.de. Berliner rundfunk Open Air 2024. mix1.de, 2024.
[50] WAZ. Apache 207 in Dortmund. waz.de, 2024.
[51] Rhein-Ruhr Aktuell. Schlagerfest XXL in Dortmund. rheinruhraktuell.de, 2024.
[52] Dogs & Fun. Dogs & Fun – Messe Dortmund. dogs-and-fun.com, 2024.
[53] Ruhr24. Dortmund OLE – Revierpark Wischlingen. ruhr24.de, 2024.
[54] Japanisches Generalkonsulat Düsseldorf. Japan day Düsseldorf 2024. dus.emb-japan.go.jp, 2024.
[55] DoKomi. Dokomi 2024 – Messe Düsseldorf. dokomi.de, 2024.
[56] tonight.de. Rheinkirmes Düsseldorf 2024. tonight.de, 2024.
[57] Time For Metal. Five finger death punch in Frankfurt. time-for-metal.eu, 2024.
[58] Football Aktuell. American football in Frankfurt – PSD Bank Arena. football-aktuell.de, 2024.
[59] Hessenschau. Rammstein in Frankfurt – Waldstadion. hessenschau.de, 2024.
[60] Football Aktuell. American football in Frankfurt – PSD Bank Arena. football-aktuell.de, 2024.
[61] Cityguide Rhein-Neckar. Peter maffay in Frankfurt – Deutsche Bank Park. cityguide-rhein-neckar.de, 2024.
[62] Cityguide Rhein-Neckar. Roland kaiser in Frankfurt – Deutsche Bank Park. cityguide-rhein-neckar.de, 2024.
[63] WAZ. AC/DC in Gelsenkirchen – Veltins-Arena. waz.de, 2024.
[64] Radio Emscher Lippe. AC/DC in Gelsenkirchen – zweites konzert. radioemscherlippe.de, 2024.
[65] Die Welt. Taylor swift in Gelsenkirchen – Veltins-Arena. welt.de, 2024.
[66] Schalke 04. Schalke tach – 75.000 fans in Gelsenkirchen. schalke04.de, 2024.
[67] Festivals United. Rammstein in Gelsenkirchen – Veltins-Arena. festivalsunited.com, 2024.
[68] NDR. Hafengeburtstag Hamburg 2024. ndr.de, 2024.
[69] Sounds and Books. Marius Müller-Westernhagen live in Hamburg 2024. soundsandbooks.com, 2024.
[70] ARD Mediathek. Taylor swift in Hamburg – Volksparkstadion. ardmediathek.de, 2024.
[71] Express Köln. Peter maffay in Köln – Rhein-Energie-Stadion. express.de, 2024.
[72] Der Westen. Roland kaiser in Köln – Rhein-Energie-Stadion. derwesten.de, 2024.
[73] WDR. Christopher street day Köln 2024. wdr.de, 2024.
[74] Leipziger Volkszeitung. Apache 207 in Leipzig – QUARTERBACK Immobilien ARENA. lvz.de, 2024.
[75] Bildlexikon Leipzig. Leipziger weinfest. bildlexikon-leipzig.de, 2024.
[76] Stadt Leipzig. Leipziger stadtfest 2024. leipzig.de, 2024.
[77] QUARTERBACK Immobilien ARENA. Andre rieu in Leipzig. quarterback-immobilien-arena.de, 2024.
[78] Radio SAW. P!NK in Leipzig – Red Bull Arena. radio-saw.de, 2024.
[79] Leipziger Volkszeitung. Roland kaiser in Leipzig – Red Bull Arena. lvz.de, 2024.
[80] RND. Peter maffay in Leipzig – Red Bull Arena. rnd.de, 2024.
[81] QUARTERBACK Immobilien ARENA. SDP in Leipzig – QUARTERBACK Immobilien ARENA. quarterback-immobilien-arena.de, 2024.
[82] Burn Your Ears. Metallica in München – Olympiastadion. burnyourears.de, 2024.
[83] Abendzeitung München. AC/DC in München – Olympiastadion. abendzeitung-muenchen.de, 2024.
[84] Merkur. Andreas gabalier in München – Olympiastadion. merkur.de, 2024.
[85] ANTENNE BAYERN. ANTENNE BAYERN Open Air – UEFA EURO 2024 fanzone München. antenne.group, 2024.
[86] München Ticket. Tollwood sommerfestival München. muenchenticket.de, 2024.
[87] Werner Consult. B2Run München. wernerconsult.com, 2024.
[88] Allgäuer Zeitung. Taylor swift in München – Olympiastadion. allgaeuer-zeitung.de, 2024.
[89] Hallenduo. Bushido in Stuttgart – Porsche-Arena. hallenduo.de, 2024.
[90] Zeitungsverlag Waiblingen. Fußball-EM 2024: Fan zones in Stuttgart mit public viewing, konzert und gaming. zvw.de, 2024.
[91] Stuttgarter Nachrichten. Peter maffay in Stuttgart – Cannstatter Wasen. stuttgarter-nachrichten.de, 2024.
[92] SWR. AC/DC in Stuttgart – Cannstatter Wasen. swr.de, 2024.
[93] Stuttgarter Nachrichten. P!NK in Stuttgart – MHP Arena. stuttgarter-nachrichten.de, 2024.
[94] Stuttgarter Nachrichten. SDP in Stuttgart – Cannstatter Wasen. stuttgarter-nachrichten.de, 2024.

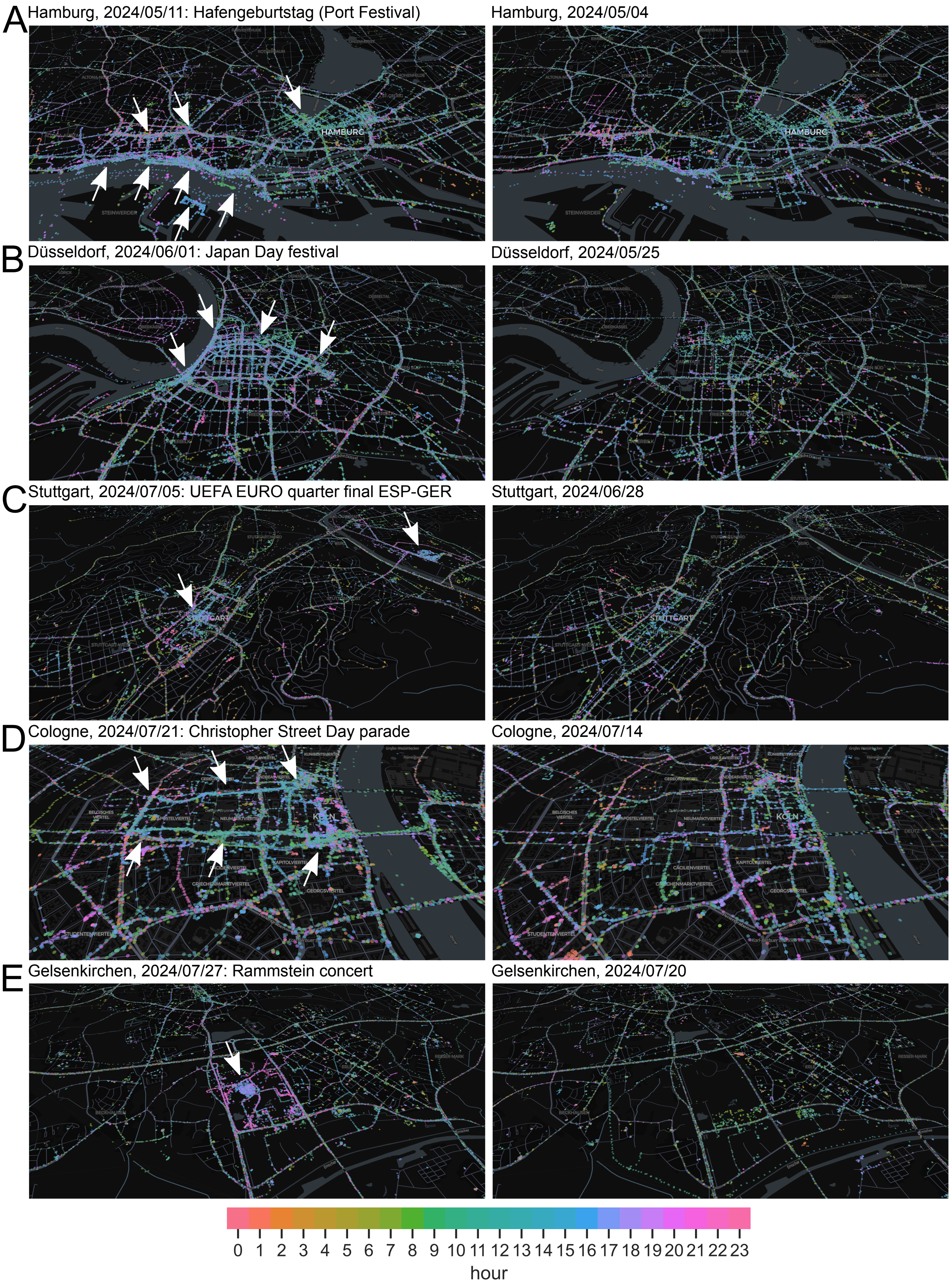
A
Hamburg, 2024/05/11: Hafengeburtstag (Port Festival)
Hamburg, 2024/05/04
B
Düsseldorf, 2024/06/01: Japan Day festival
Düsseldorf, 2024/05/25
C
Stuttgart, 2024/07/05: UEFA EURO quarter final ESP-GER
Stuttgart, 2024/06/28
D
Cologne, 2024/07/21: Christopher Street Day parade
Cologne, 2024/07/14
E
Gelsenkirchen, 2024/07/27: Rammstein concert
Gelsenkirchen, 2024/07/20
0 1 2 3 4 5 6 7 8 9 10 11 12 13 14 15 16 17 18 19 20 21 22 23
hour

FIG. S1. **GPS location data collected from mobile phone users.** The raw data for this contact study consists of data points, or "pings", collected from mobile phones. Each ping is endowed with a unique and persistent device ID, a GPS location (here represented as dots on the map) and a timestamp (here represented by color). Each dot in the map corresponds to one such ping and indicates that the phone user was located at the shown location at the specified time. Left panels show the pings recorded in host cities on the day of a MGE (as indicated), whereas right panels show pings on a regular day as a baseline (7 days prior to the indicated MGE). White arrows indicate locations with increased data creation on the MGE day.

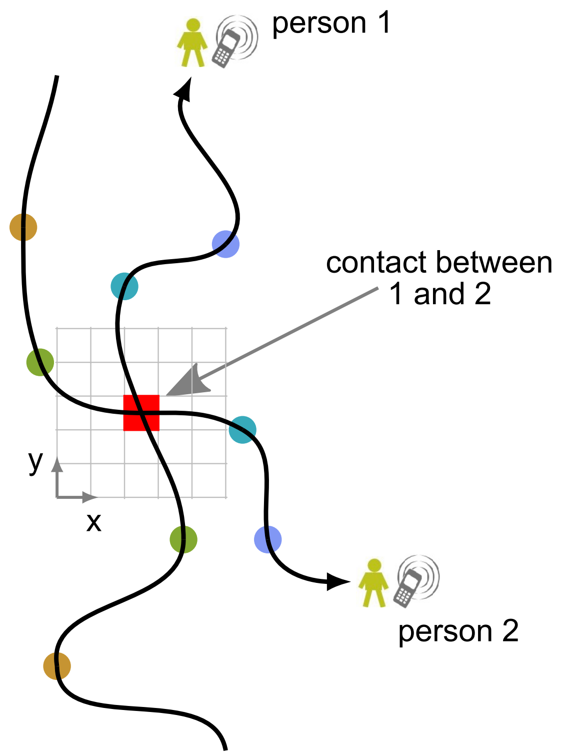


FIG. S2. **Definition of co-location contact.** The trajectories of mobile phone users (black arrows) are sampled at discrete timepoints (colored dots) and the timestamps and GPS locations of samples are recorded alongside persistent device IDs. A contact is defined as the co-location of two (or more) mobile devices within the same 16 m-by-16 m cell (red square) on a pre-defined lattice (gray grid) at the same time modulo 10 min.

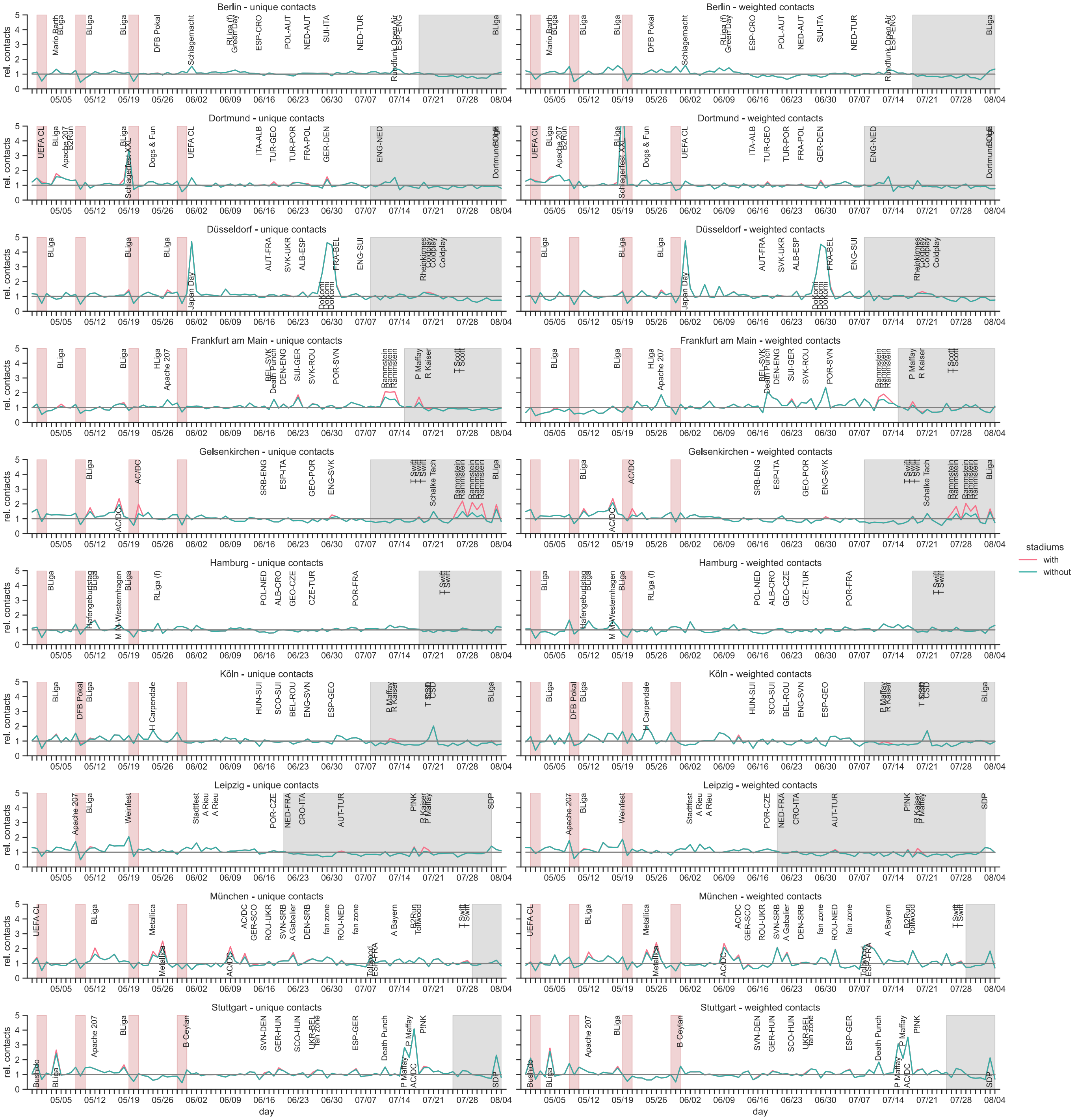


FIG. S3. **Contact variations in MGE host cities.** Extended version of Fig. 1. Relative detrended daily contact levels $c^i_{w,t} = C^i_{w,t}/C^i_t$ between April 29 and August 4, 2024 in all 10 host cities $i$ of UEFA EURO 2024 (rows). Contacts $C^i_{w,t}$ are counted either as "unique" contacts (left panels) or as "weighted" contacts (right panels). The distinction resides in counting repeatedly co-located devices within a day once or as many times as they were observed. The weekday-dependent baseline is given by $C^i_t = \frac{1}{W}\sum_{w=1}^{W} C^i_{w,t}$. Data is shown when contacts inside the host stadiums of UEFA EURO 2024 are included (red line) or excluded (green line) from the contact count $C^i_{w,t}$ of city $i$. Public holidays and school holidays are indicated by red and gray shading, respectively (note the regional differences in public holiday observance and school holiday schedules). Major MGEs in these 10 cities are indicated (Tab. S14).

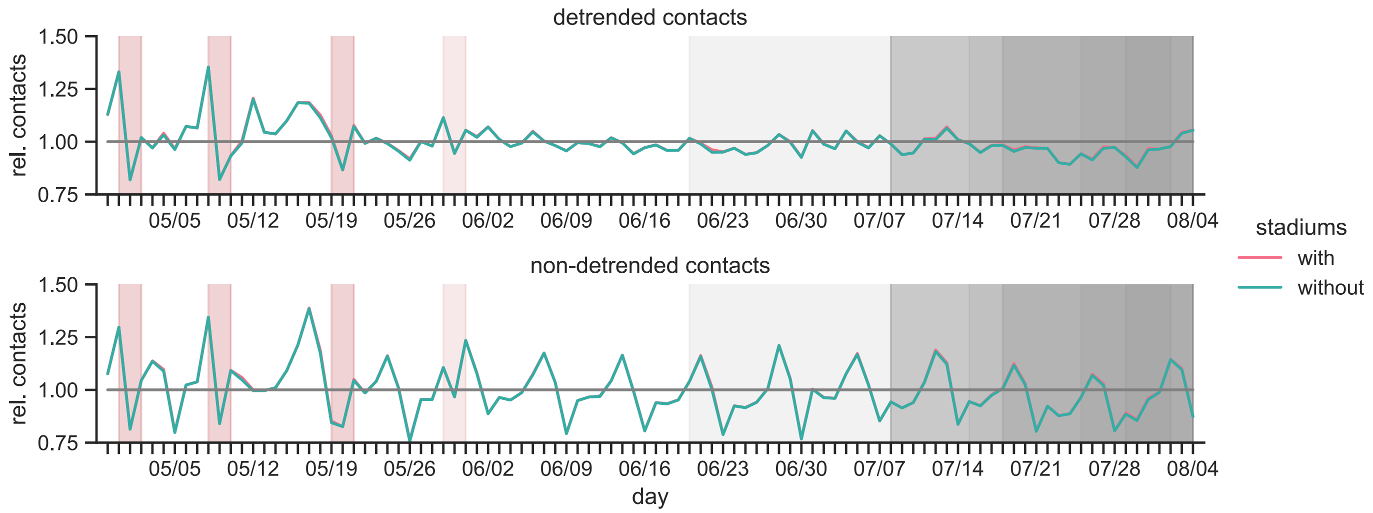


FIG. S4. **Nationwide contact variations.** Relative daily contact levels between April 29 and August 4, 2024 across Germany at-large. The data is shown as **(A)** detrended signal $c^i_{w,t} = C^i_{w,t}/C^i_t$ with $C^i_t = \frac{1}{W}\sum_{w=1}^{W} C^i_{w,t}$ and as **(B)** non-detrended signal $c^i_{w,t} = C^i_{w,t}/C^i$ with $C^i = \frac{1}{7W}\sum_{w=1}^{W}\sum_{t=1}^{7} C^i_{w,t}$. Contacts $C^i_{w,t}$ are counted as "unique" contacts. Data is shown when contacts inside the host stadiums of UEFA EURO 2024 are included (red line) or excluded (green line) from the contact count $C^i_{w,t}$ of city $i$. Public holidays and school holidays are indicated by red and gray shading, respectively (note the regional differences in public holiday observance and school holiday schedules which is reflected in the different shadings).

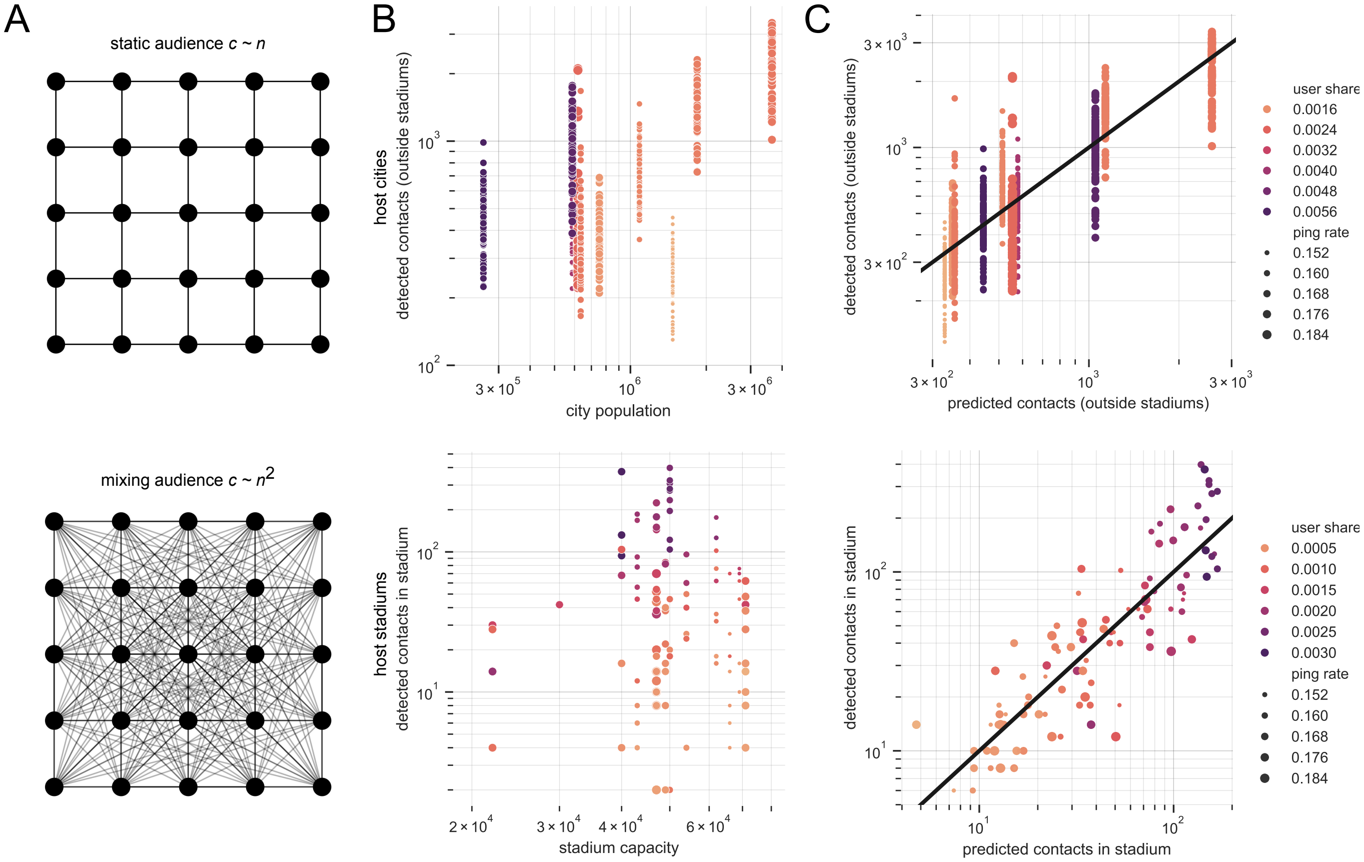


FIG. S5. **Scaling of contact numbers with cohort size.** **(A)** Schematic network representation of individuals in the nearest-neighbor (top panel) and fully mixed (bottom panel) contact scenarios: Every individual in the cohort (nodes) is in contact (lines) only with its nearest neighbors or with every other individual [22]. These scenarios represent opposite extreme cases in which contact numbers $c$ scale linearly ($\beta = 1$) and quadratically ($\beta = 2$) with the number of individuals $n$. **(B)** Relationship between observed contact numbers $c$ (vertical axes), cohort sizes $N$ (horizontal axes), user percentage $p = n/N$ (color) and data generation activity $r = s/n$ (dot size). This relationship is plotted separately for host cities (top panel) and host stadiums (bottom panel) of UEFA EURO 2024. Values for $N$, $p$ and $r$ for host cities are provided in Tabs. S1 and S4. Note the log-log scale. **(C)** Fit of a scaling model (Eq. (S1)) to relate $c$, $N$, $p$ and $r$. The plot shows observed contacts $c$ (vertical axes) vs. contacts predicted from $N$, $p$ and $r$ using the scaling law (horizontal axes) along with the diagonal $y = x$ (black line). The scaling model is fitted separately for host cities (top panel) and host stadiums alone (bottom panel) using Ordinary Least Squares (OLS) on log-transformed data.

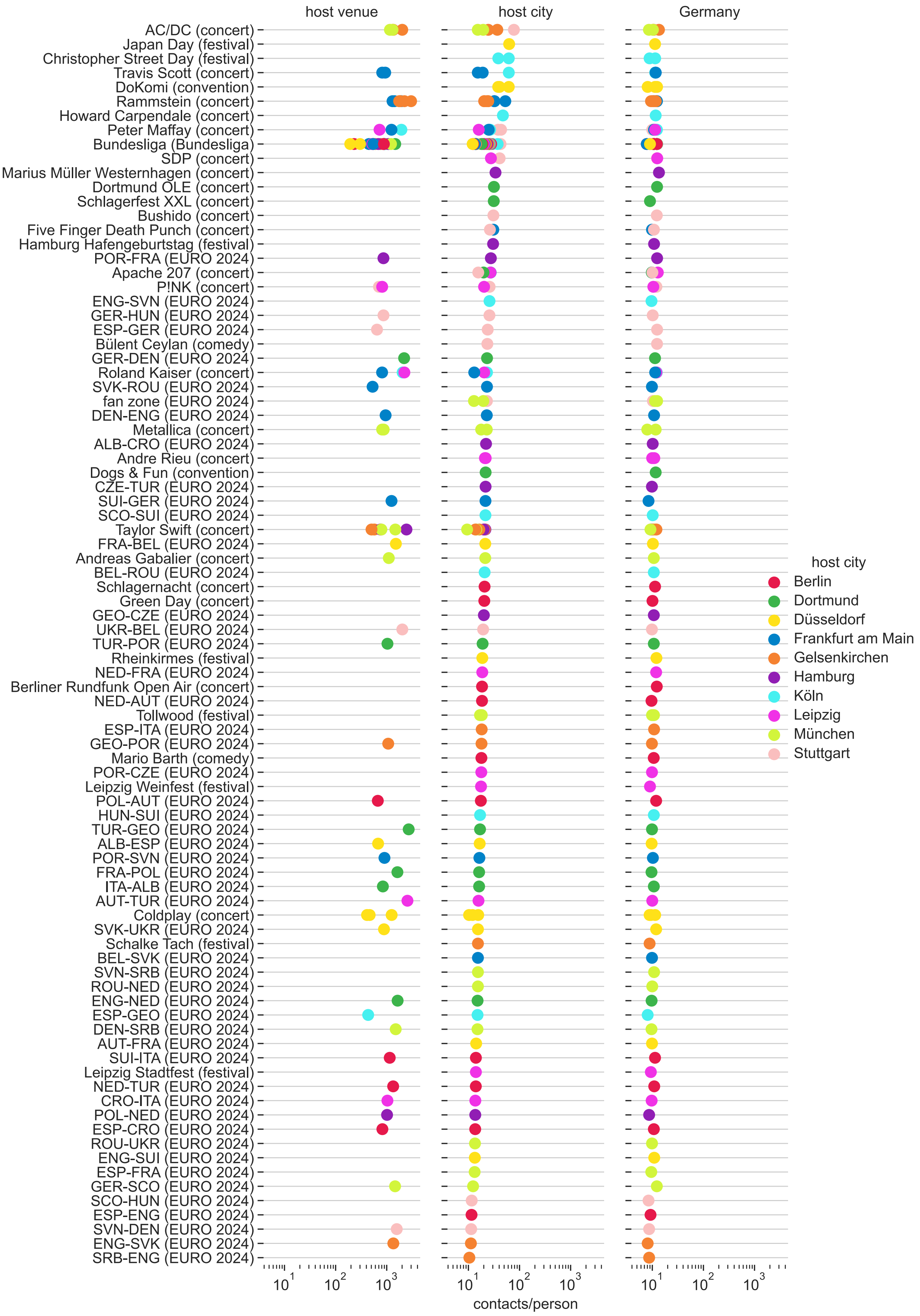
host venue
host city
Germany
AC/DC (concert)
Japan Day (festival)
Christopher Street Day (festival)
Travis Scott (concert)
DoKomi (convention)
Rammstein (concert)
Howard Carpendale (concert)
Peter Maffay (concert)
Bundesliga (Bundesliga)
SDP (concert)
Marius Müller Westernhagen (concert)
Dortmund OLE (concert)
Schlagerfest XXL (concert)
Bushido (concert)
Five Finger Death Punch (concert)
Hamburg Hafengeburtstag (festival)
POR-FRA (EURO 2024)
Apache 207 (concert)
P!NK (concert)
ENG-SVN (EURO 2024)
GER-HUN (EURO 2024)
ESP-GER (EURO 2024)
Bülent Ceylan (comedy)
GER-DEN (EURO 2024)
Roland Kaiser (concert)
SVK-ROU (EURO 2024)
fan zone (EURO 2024)
DEN-ENG (EURO 2024)
Metallica (concert)
ALB-CRO (EURO 2024)
Andre Rieu (concert)
Dogs & Fun (convention)
CZE-TUR (EURO 2024)
SUI-GER (EURO 2024)
SCO-SUI (EURO 2024)
Taylor Swift (concert)
FRA-BEL (EURO 2024)
Andreas Gabalier (concert)
BEL-ROU (EURO 2024)
Schlagernacht (concert)
Green Day (concert)
GEO-CZE (EURO 2024)
UKR-BEL (EURO 2024)
TUR-POR (EURO 2024)
Rheinkirmes (festival)
NED-FRA (EURO 2024)
Berliner Rundfunk Open Air (concert)
NED-AUT (EURO 2024)
Tollwood (festival)
ESP-ITA (EURO 2024)
GEO-POR (EURO 2024)
Mario Barth (comedy)
POR-CZE (EURO 2024)
Leipzig Weinfest (festival)
POL-AUT (EURO 2024)
HUN-SUI (EURO 2024)
TUR-GEO (EURO 2024)
ALB-ESP (EURO 2024)
POR-SVN (EURO 2024)
FRA-POL (EURO 2024)
ITA-ALB (EURO 2024)
AUT-TUR (EURO 2024)
Coldplay (concert)
SVK-UKR (EURO 2024)
Schalke Tach (festival)
BEL-SVK (EURO 2024)
SVN-SRB (EURO 2024)
ROU-NED (EURO 2024)
ENG-NED (EURO 2024)
ESP-GEO (EURO 2024)
DEN-SRB (EURO 2024)
AUT-FRA (EURO 2024)
SUI-ITA (EURO 2024)
Leipzig Stadtfest (festival)
NED-TUR (EURO 2024)
CRO-ITA (EURO 2024)
POL-NED (EURO 2024)
ESP-CRO (EURO 2024)
ROU-UKR (EURO 2024)
ENG-SUI (EURO 2024)
ESP-FRA (EURO 2024)
GER-SCO (EURO 2024)
SCO-HUN (EURO 2024)
ESP-ENG (EURO 2024)
SVN-DEN (EURO 2024)
ENG-SVK (EURO 2024)
SRB-ENG (EURO 2024)
$10^1$
$10^2$
$10^3$
contacts/person
host city
Berlin
Dortmund
Düsseldorf
Frankfurt am Main
Gelsenkirchen
Hamburg
Köln
Leipzig
München
Stuttgart

FIG. S6. **Locally contained and type-dependent contact increases in MGE host cities.** Extended version of Fig. 2 including daily mean number of contacts per person $C/N$ for host venues (left column) while showing the same data as in Fig. 2 for host cities and Germany (center and right columns). These additional values represent contact numbers per person associated with a person being inside the MGE venue. Note that, unlike in Fig. 2, data is shown on a logarithmic scale.

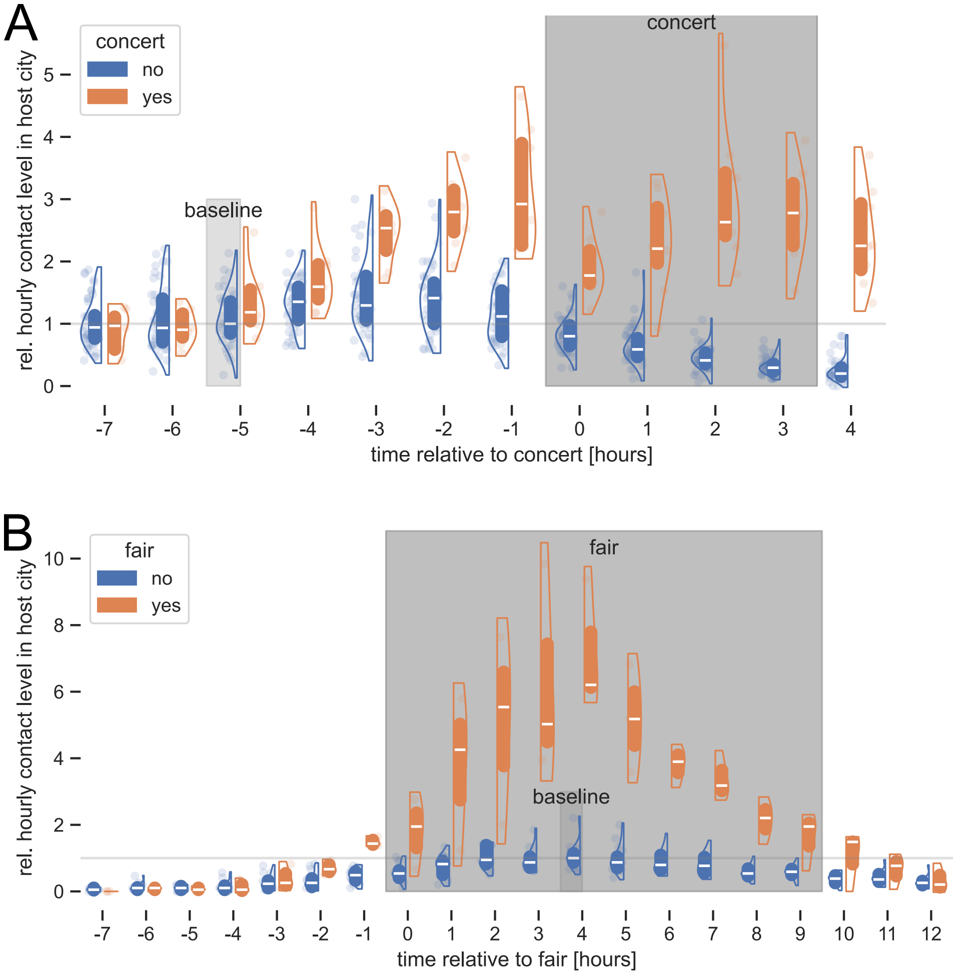


FIG. S7. **Temporal distribution of contacts in MGE host cities.** Relative hourly contact levels in host cities of **(A)** Rammstein concerts ($n = 8$; Tab. S14) and **(B)** DoKomi fair ($n = 3$; Tab. S14) on MGE days (orange violins) vs. non-MGE days (blue violins; same weekday), relative to the start time of the MGE. Relative contact levels are given as fold change against 2 pm on non-MGE days, where the median value is defined to be 1 (light gray shading). The time covered by the MGE is highlighted by dark gray shading. Boxes represent center quartiles (50 % of data) and white bars inside boxes represent the median, dots represent individual data points.

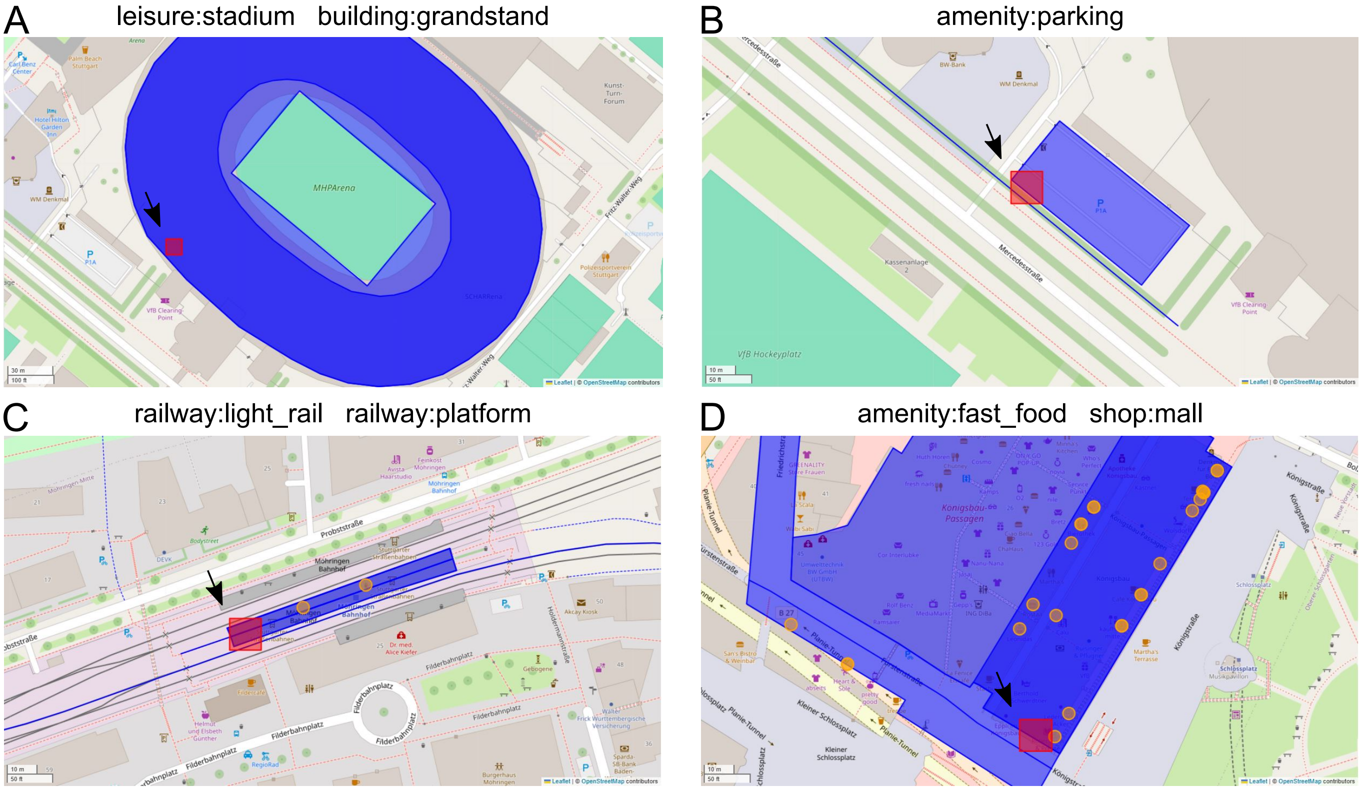


FIG. S8. **OpenStreetMap projection of GPS contacts.** Examples of contact setting inference by matching the location of GPS contacts (black arrows and red square showing the 16 m-by-16 m cell of the co-location) and OpenStreetMap map features (blue contours, blue lines and orange dots). The OSM key-value pairs characterizing the matched map features are indicated: **(A)** The observed stadium contact is ascribed to settings `leisure:stadium` and `building:grandstand`. In the other examples, the contact settings are: **(B)** `amenity:parking`, **(C)** `railway:light_rail` and `railway:platform`, **(D)** `amenity:fast_food` and `shop:mall`.

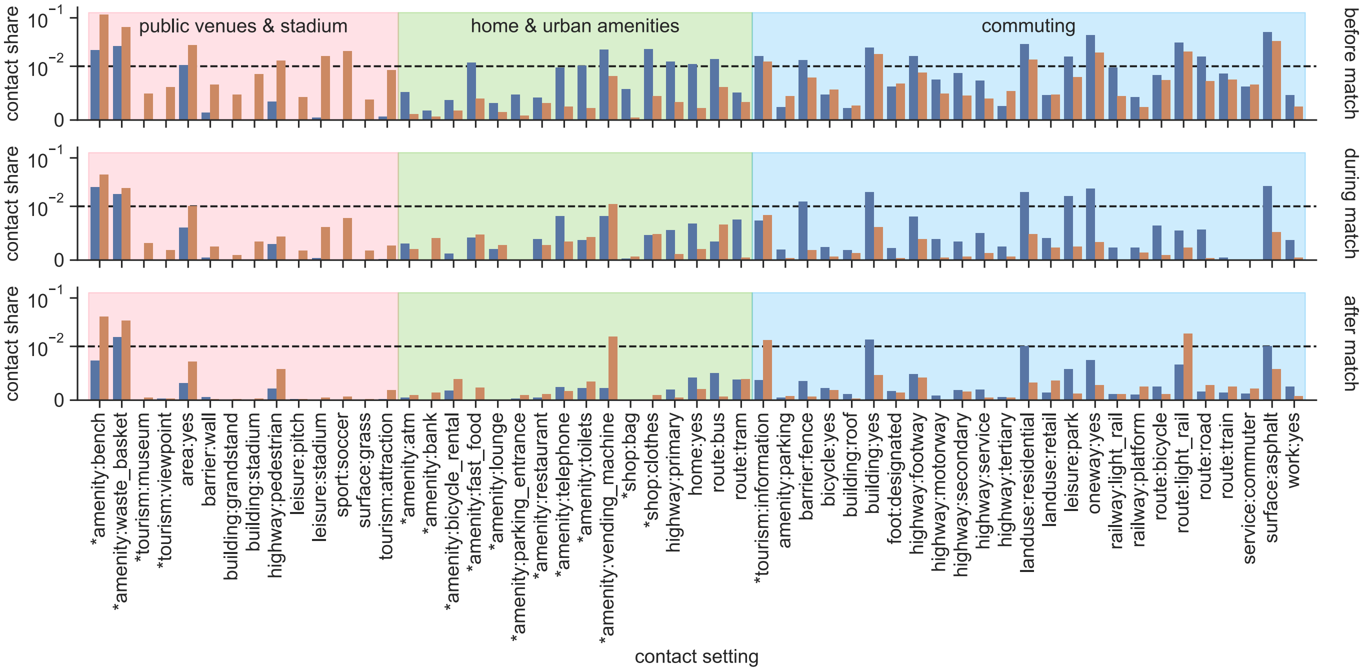


FIG. S9. **Distribution of contact settings in MGE host cities.** Spatio-temporal distribution of contacts in host cities of UEFA EURO 2024 matches with German participation on match days (orange bars) and baseline non-MGE days (blue bars). The time bins are defined relative to the start of the matches: 2 hours before (top panel), during (center panel) and after (bottom panel) the match start time. Contact settings correspond to key-value descriptors of OpenStreetMap map features. Bars indicate the percentage of all daily contacts in each contact setting and time bin. Only contact settings with at least 0.1 % in at least one of the time bins are shown. Contact settings are grouped by similarity of contact dynamics during MGEs (colored shading; Fig. 4). Note that contact shares do not necessarily sum to 1, as contacts may be associated with multiple settings (Fig. S8). Setting highlighted by asterisk ($*$) correspond to 0D OSM map features (nodes), while others correspond to 2D and 1D map features (polygons and lines). The plot uses a log-linear scale on vertical axes; the dashed black line delineates the logarithmic and linear scales.

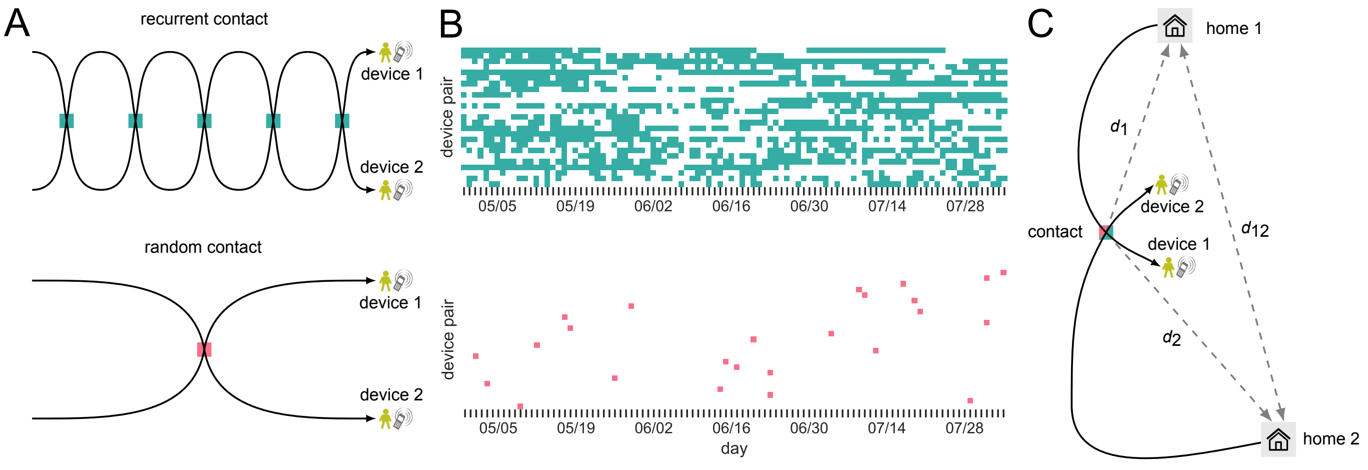


FIG. S10. **Topological features of GPS contacts.** **(A)** Schematic representation of “recurrent” (top panel) vs. “random” (bottom panel) contacts: A recurrent contact is repeated on distinct days between the same individuals (e.g. partners, co-workers), identified through repeated co-location of their individual trajectories. A random contact is a one-time encounter between otherwise non-interacting individuals (e.g. strangers attending the same MGE), identified through a one-time co-location of otherwise distant trajectories. **(B)** Temporal footprint of recurrent vs. random contacts: Each row represents a fixed pair of devices in the data. The time series then indicates on daily basis for each pair whether or not a co-location was observed (colored dots). **(C)** The home-to-home distance $d_{12}$ of a contact is defined as the distance between the home locations of the two individuals involved in the co-location. The contact-to-home distances $d_1$ and $d_2$ of a contact are defined as the distance of the contact location to each of the home locations of the two individuals involved in the co-location.

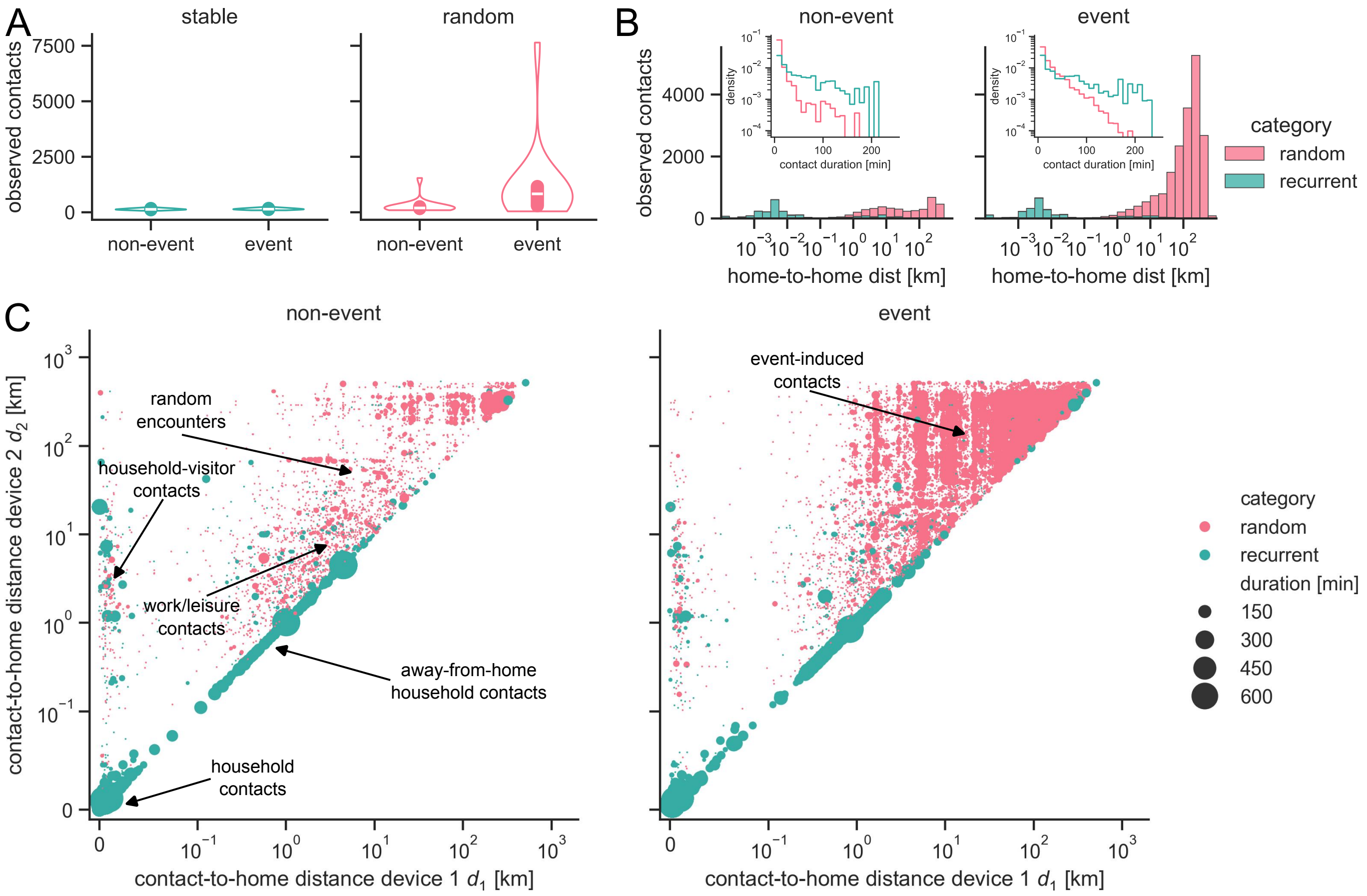


FIG. S11. **Topological effects of MGE contacts in Gelsenkirchen. (A)** Same as Fig. 5A, but showing the distribution of daily "recurrent" and "random" contact numbers for the UEFA EURO 2024 host city of Gelsenkirchen alone. Gelsenkirchen is an unusually small host city for MGE of this scale, thus showing low baseline non-MGE contact levels. **(B)** Same as Fig. 5B, but showing home-to-home distance ($d_{12}$) and contact duration distributions for the UEFA EURO 2024 host city of Gelsenkirchen alone. **(C)** Scatter plots combining information about contact-to-home distances $d_1$ and $d_2$ and cumulative contact duration of device pairs, separately for MGE days (right panel) and non-MGE days (left panel) in the UEFA EURO 2024 host city of Gelsenkirchen. Each dot represents a single pair of devices and indicates how far the contact location is from the home of each of the contact partners (vertical and horizontal axes; note the log-log scale), for how long the contact lasted (dot size; observed cumulative duration) and whether or not the contact was repeated on distinct days (dot color). Different contact modes are highlighted and labeled. Note that pairs are arbitrarily ordered to satisfy $d_1 \leq d_2$, which is why only the upper triangle in the scatter plots is populated.

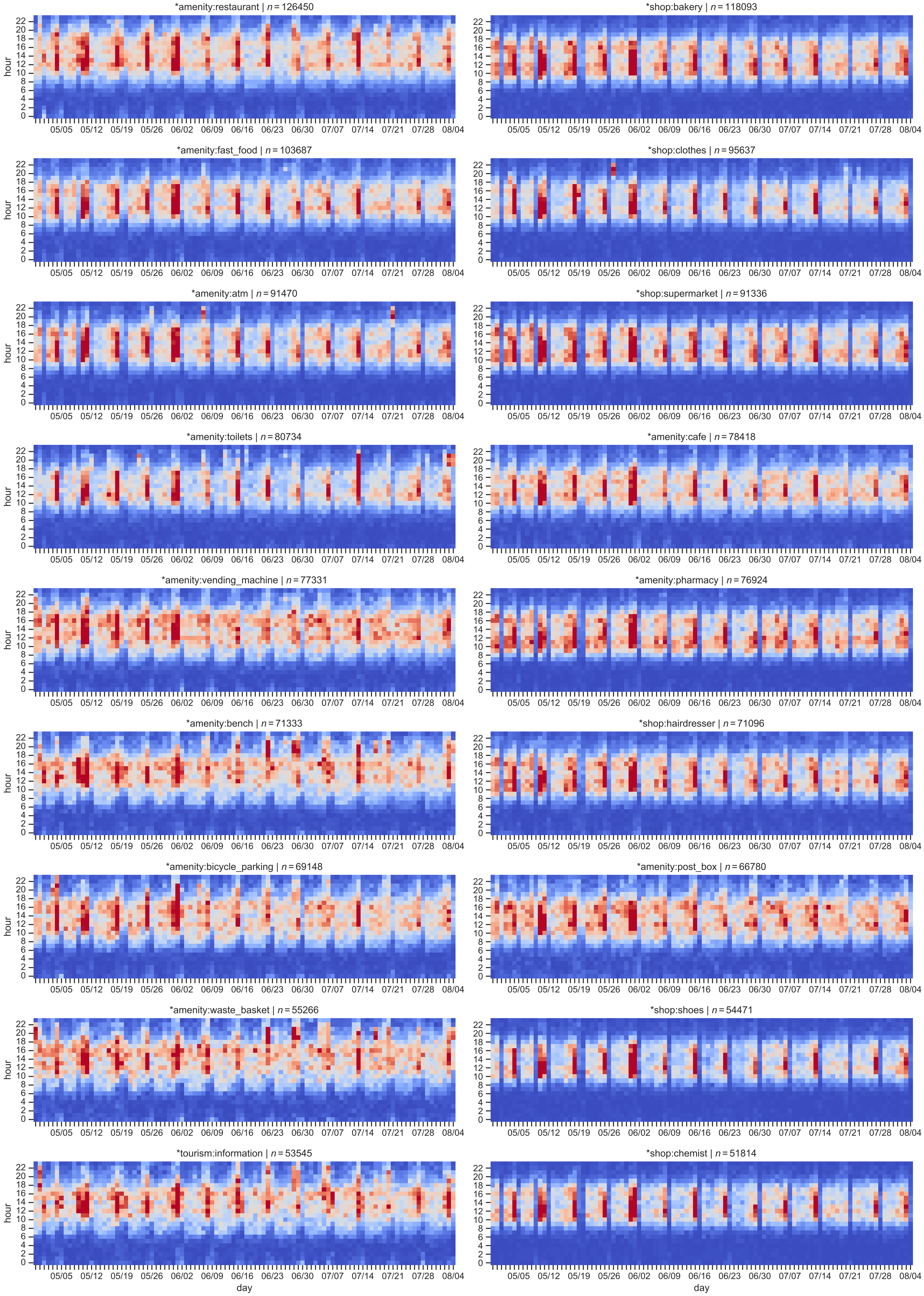


FIG. S12. **Germany-wide spatiotemporal distribution of contacts.** Heatmaps showing relative contact intensities across Germany by contact setting (different panels), day (horizontal axis) and hour (vertical axis) over the entire study period. Red (blue) color indicates high (low) intensity of contacts. The colorbar is scaled to represent the central 95 % of data within each panel (thus avoiding domination by outliers). Heatmaps are only shown for contact settings represented as node-like map features in OpenStreetMap and to which $n \geq 3{,}000$ co-locations have been associated.

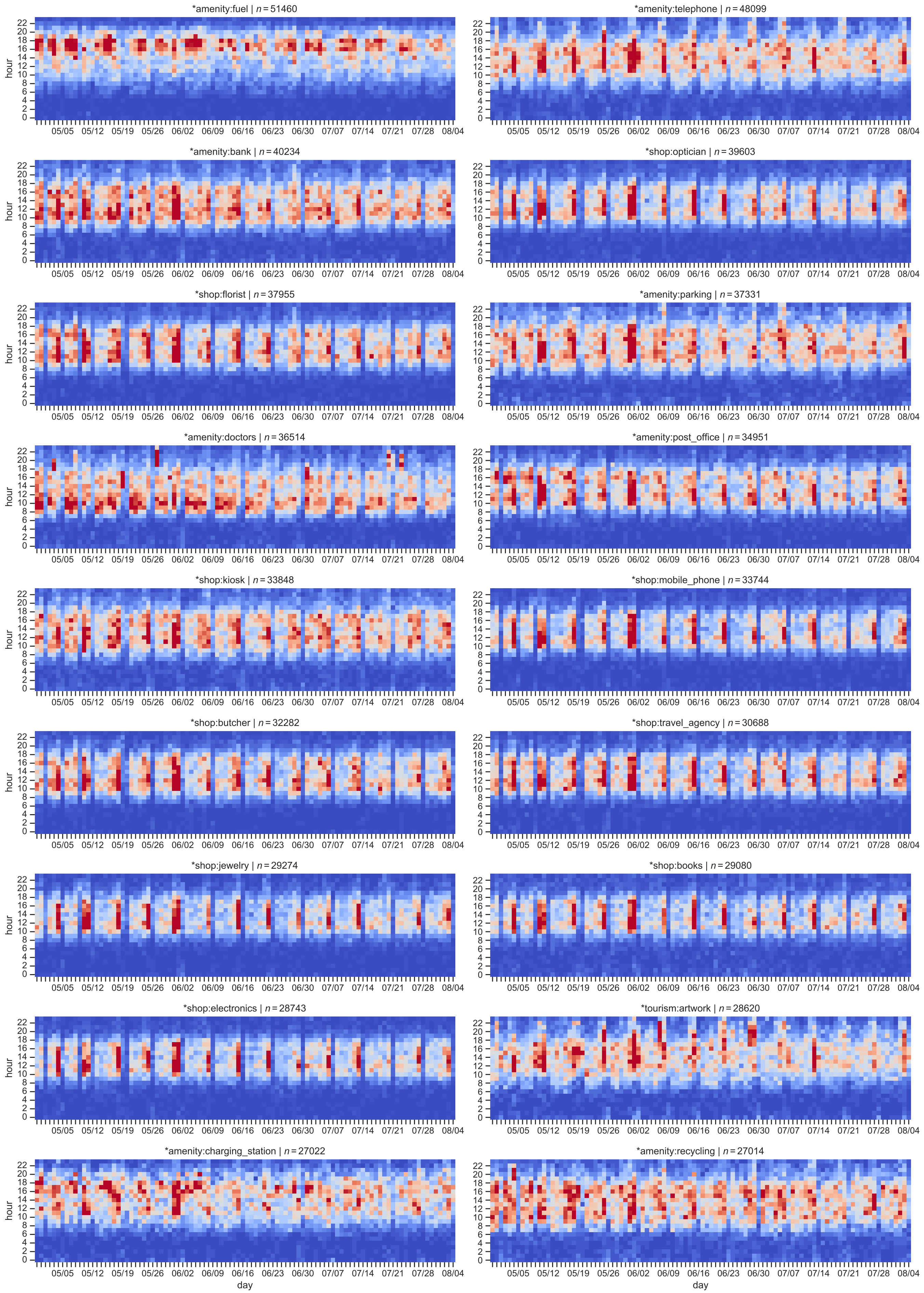


FIG. S13. **Germany-wide spatiotemporal distribution of contacts.** Continuation of Fig. S12.

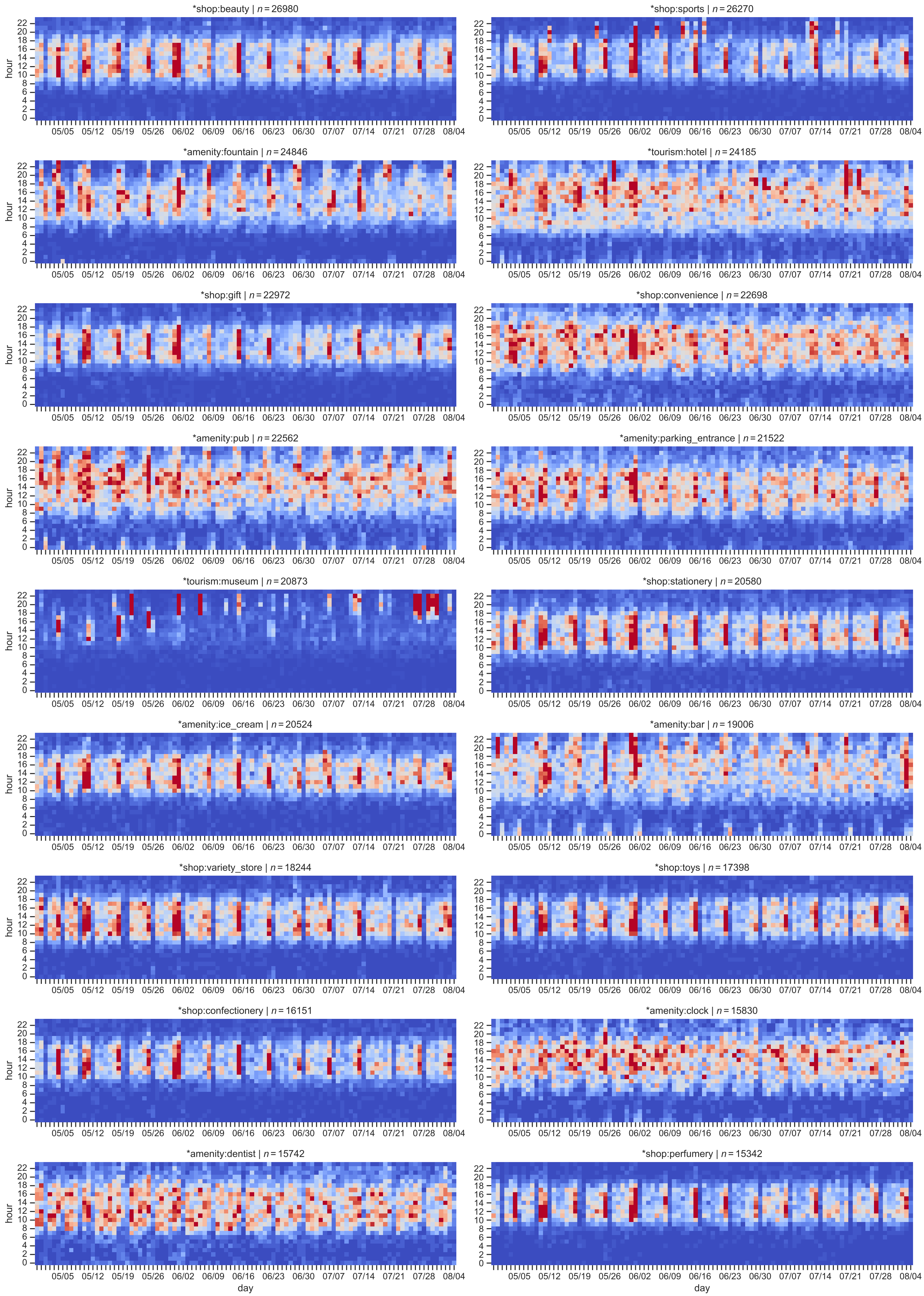


FIG. S14. **Germany-wide spatiotemporal distribution of contacts.** Continuation of Fig. S12.

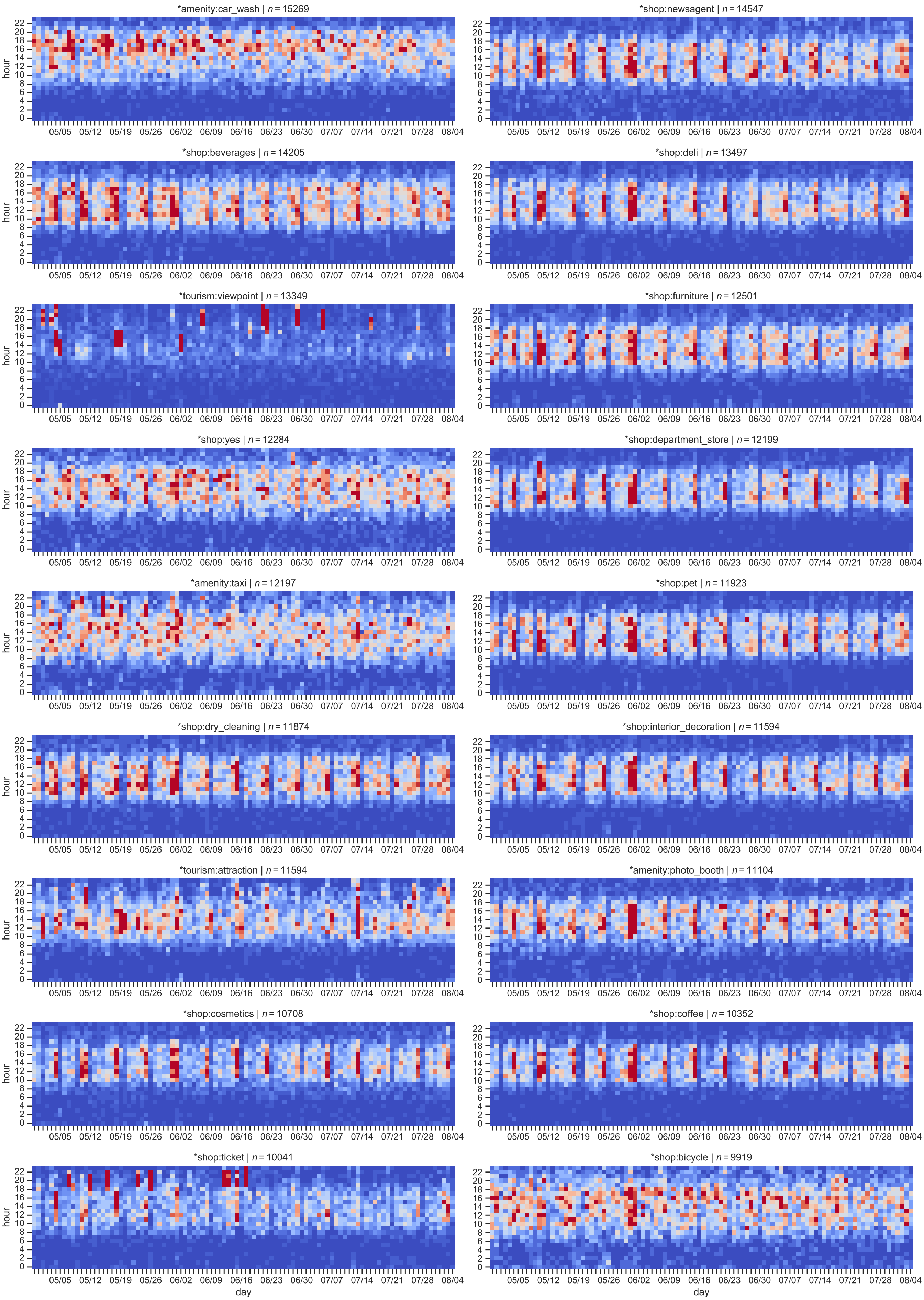


FIG. S15. **Germany-wide spatiotemporal distribution of contacts.** Continuation of Fig. S12.

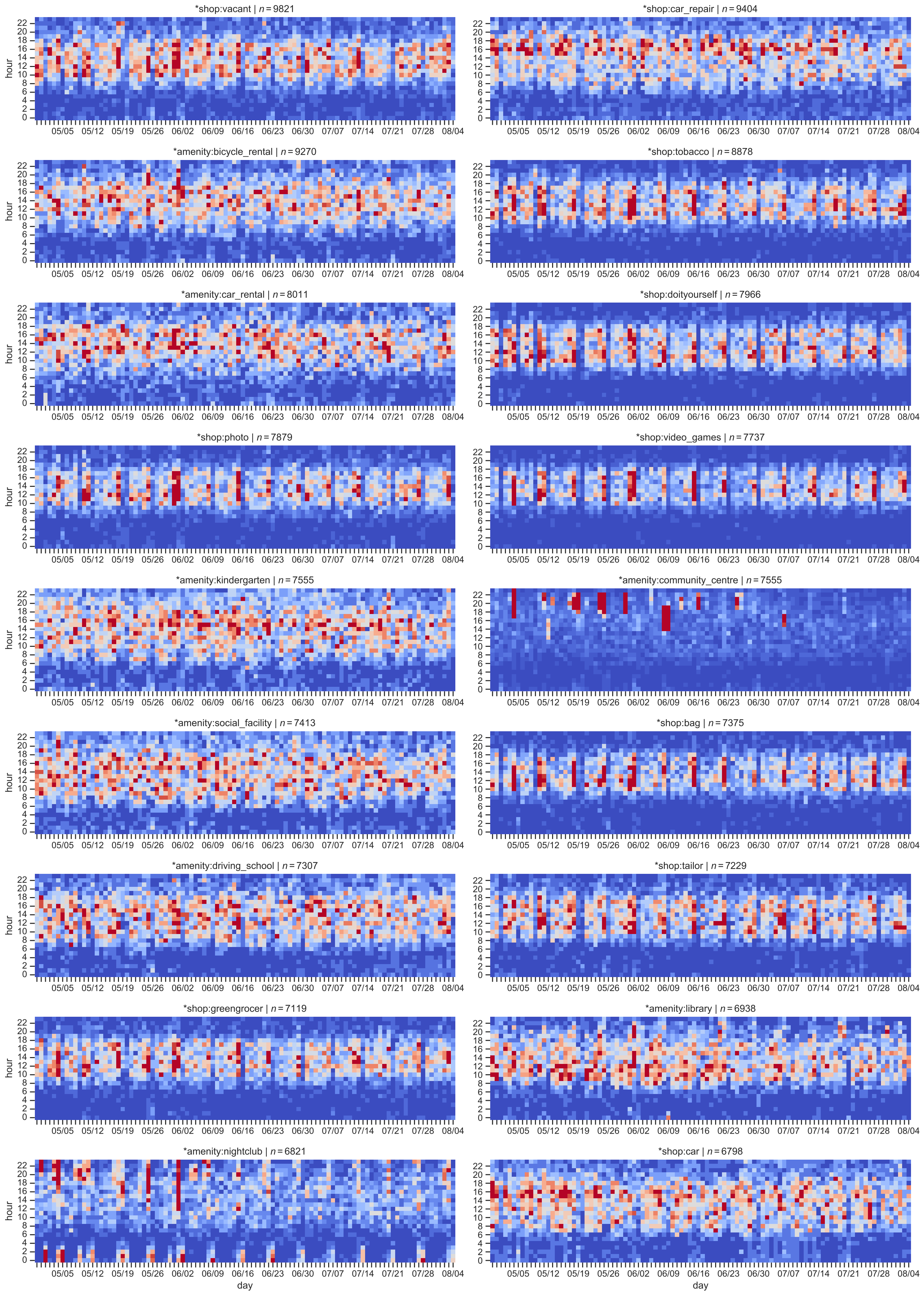


FIG. S16. **Germany-wide spatiotemporal distribution of contacts.** Continuation of Fig. S12.

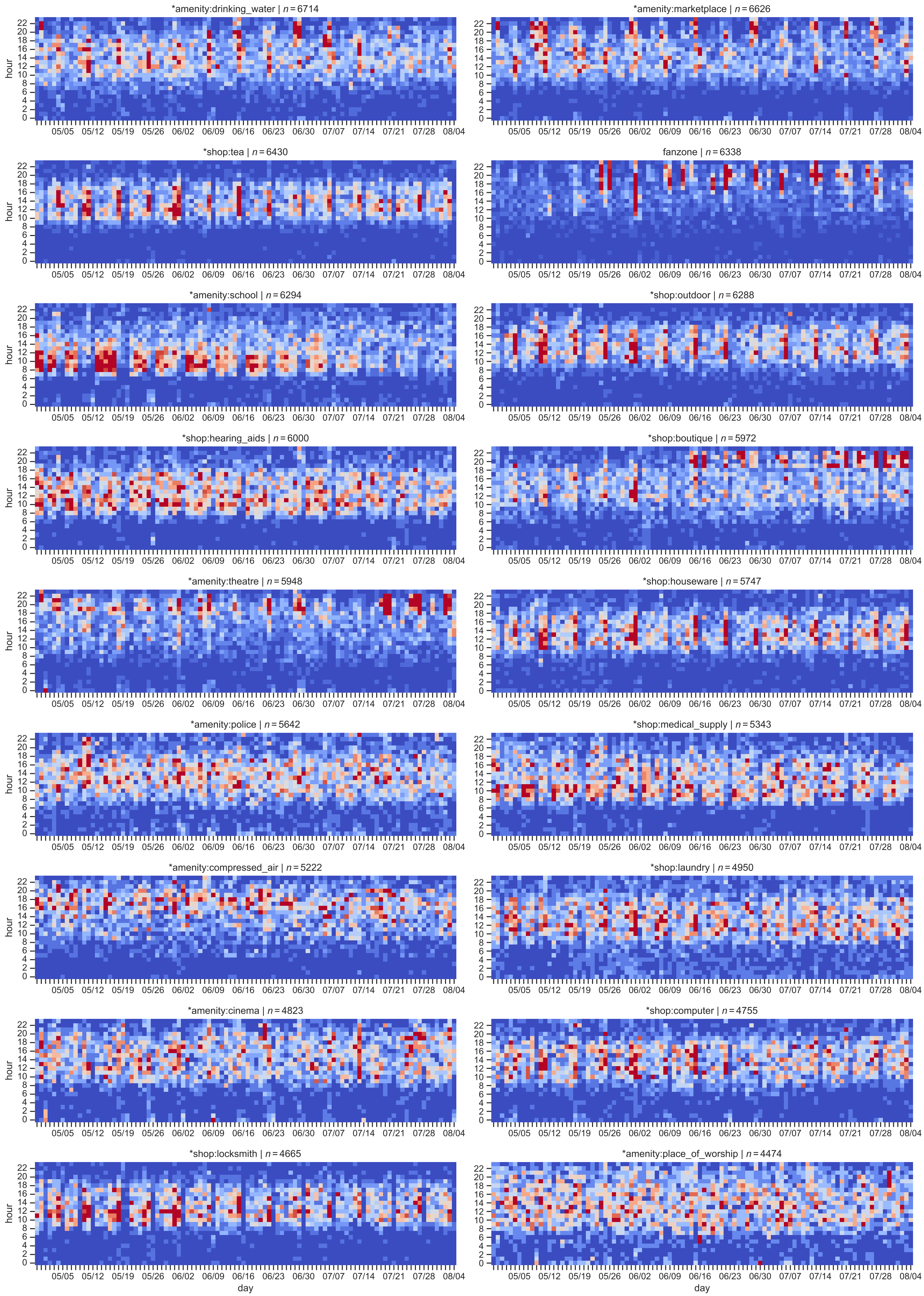


FIG. S17. **Germany-wide spatiotemporal distribution of contacts.** Continuation of Fig. S12.

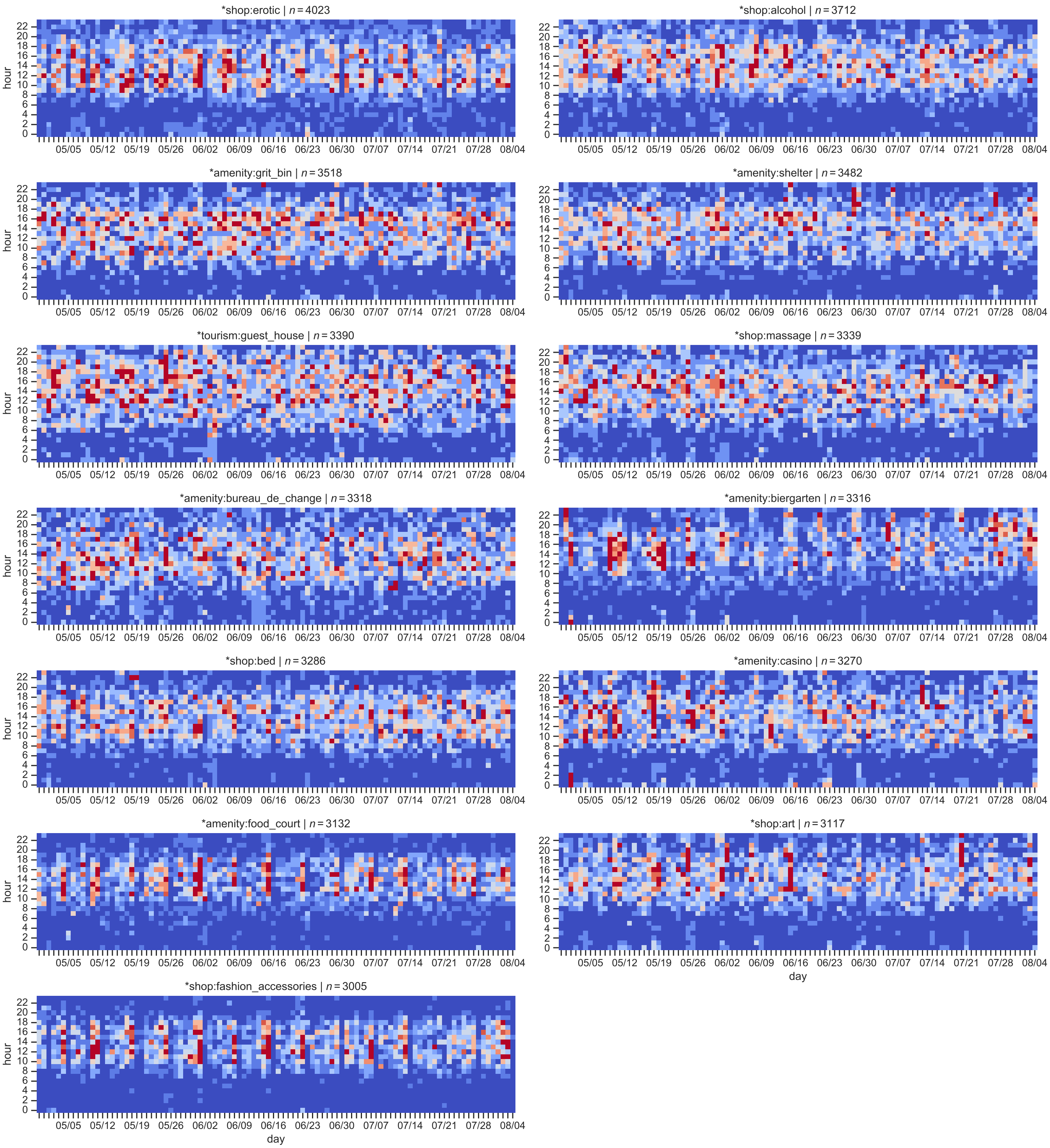


FIG. S18. **Germany-wide spatiotemporal distribution of contacts.** Continuation of Fig. S12.

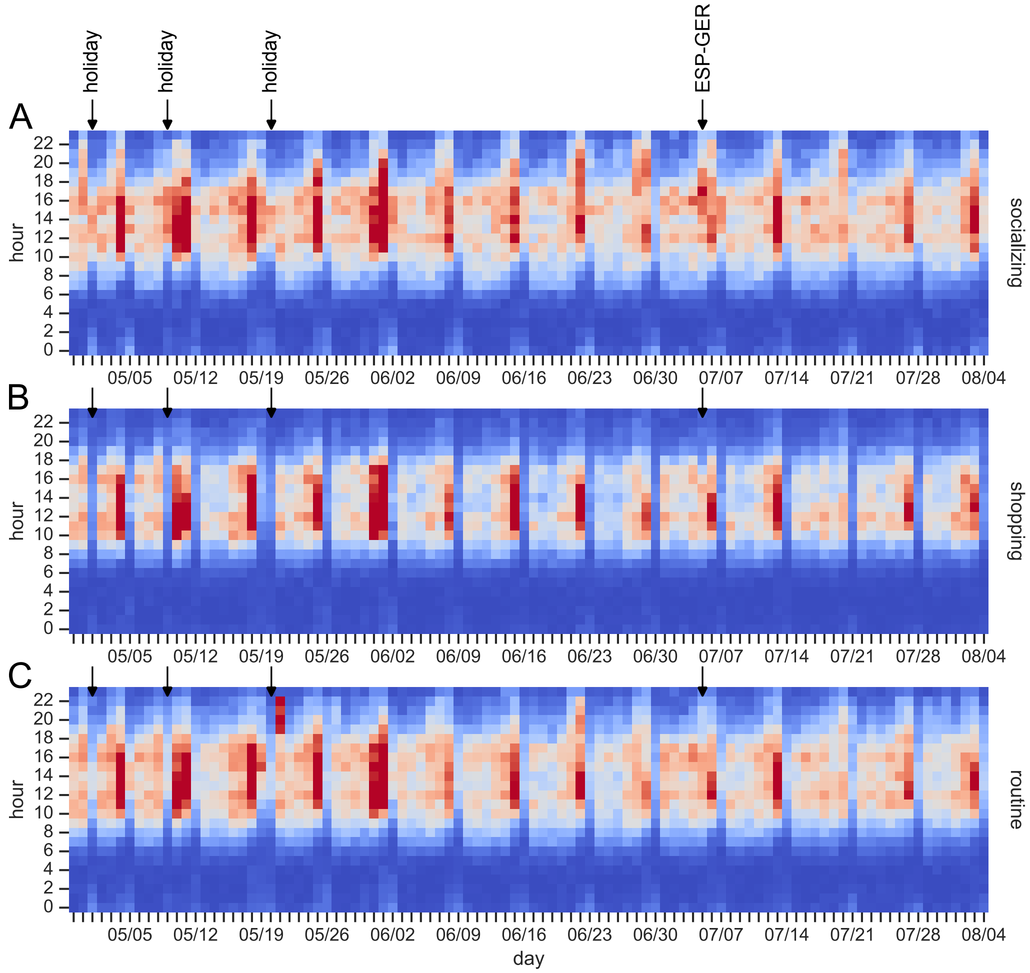


FIG. S19. **Temporal footprint of contacts in socializing, shopping and routine settings across Germany.** Heatmaps showing relative contact intensities across Germany by day (horizontal axis), hour (vertical axis) and category of settings: **(A)** socializing settings, **(B)** shopping settings, **(C)** routine settings as defined in Fig. 6 and Tab. S13. Red (blue) color indicates high (low) intensity of contacts. The colorbar is scaled to represent the central 95 % of data within each panel (thus avoiding domination by outliers). Nationwide public holidays and the day of the UEFA EURO 2024 quarter-final match ESP–GER are highlighted by black arrows.

| host city | population | host stadium | capacity | fan zones |
|---|---|---|---|---|
| Berlin | 3,645,000 | Olympiastadion | 71,000 | Brandenburg Gate<br>Reichstag |
| Dortmund | 587,000 | Signal Iduna Park | 62,000 | Westfalenpark<br>Friedensplatz |
| Düsseldorf | 619,000 | Merkur Spiel-Arena | 47,000 | Burgplatz<br>Gustaf-Gründgens-Platz<br>Untere Rheinwerft<br>Rheinpark |
| Frankfurt am Main | 753,000 | Deutsche Bank Park | 47,000 | Mainufer<br>Nizza |
| Gelsenkirchen | 261,000 | Veltins-Arena | 50,000 | Nordsternplatz<br>Amphitheater Gelsenkirchen |
| Hamburg | 1,841,000 | Volksparkstadion | 49,000 | Heiligengeistfeld |
| Cologne | 1,086,000 | RheinEnergieStadion | 43,000 | Heumarkt<br>Aachener Weiher |
| Leipzig | 588,000 | Red Bull Arena | 40,000 | Augustusplatz<br>Wilhelm-Leuschner-Platz<br>Villa Sack<br>Moritzbastei |
| Munich | 1,472,000 | Allianz Arena | 66,000 | Olympiapark |
| Stuttgart | 635,000 | MHP Arena | 54,000 | Schlossplatz<br>Marktplatz<br>Karlsplatz<br>Schillerplatz |

TABLE S1. **Host cities, host stadiums and fan zones of the UEFA EURO 2024 tournament.** The 10 host cities and host stadiums of the UEFA EURO 2024 tournament in Germany alongside their population sizes and stadium capacities [24]. The places of "fan zones" for public viewing of matches in these cities [25–34] are also listed.

| resolution | detrended daily rel. contacts $c^i_{w,t}$ (city scale) median (25th percentile; 75th percentile) | | $p$-value (MW) | |
|---|---|---|---|---|
| | non-MGE | MGE | MGE vs. non-MGE | non-MGE vs. non-MGE |
| 62 m, 10 min | 1.006 (0.902; 1.103) | 1.245 (1.069; 1.569) | $< 10^{-10}$ * | 0.69 |
| 31 m, 10 min | 0.999 (0.893; 1.109) | 1.256 (1.054; 1.570) | $< 10^{-10}$ * | 0.52 |
| 16 m, 10 min | 0.998 (0.882; 1.118) | 1.254 (1.039; 1.562) | $< 10^{-10}$ * | 0.85 |
| 8 m, 10 min | 0.996 (0.863; 1.110) | 1.267 (1.030; 1.554) | $< 10^{-10}$ * | 0.25 |
| 8 m, 60 min | 1.006 (0.872; 1.114) | 1.249 (1.002; 1.496) | $< 10^{-10}$ * | 0.64 |

TABLE S2. **Significance and sensitivity analysis of MGE-induced contacts in host cities.** Relative detrended daily contacts $c^i_{w,t} = C^i_{w,t}/C^i_t$ across 10 cities in Germany on days without ("non-MGE") vs. with ("MGE") mass gathering events. Contacts are determined using multiple alternative definitions ("resolution"). Median, 25th (Q1) and 75th (Q3) percentiles of $c^i_{w,t}$ on MGE and non-MGE days and cities are given alongside $p$-values from two-sided Mann-Whitney U tests for the hypothesis that MGE and non-MGE contact levels $c^i_{w,t}$ are equivalent ("MGE vs. non-MGE"). $p$-values for a negative control in which MGE data is randomly replaced by non-MGE data prior to the statistical test ("non-MGE vs. non-MGE") is also provided. Statistically significant differences ($p < 0.05$) are highlighted by asterisk ($*$).

| resolution | detrended daily rel. contacts $c_{w,t}$ (Germany at-large) median (25th percentile; 75th percentile) | | $p$-value (MW) | |
|---|---|---|---|---|
| | non-MGE | MGE | MGE vs. non-MGE | non-MGE vs. non-MGE |
| 62 m, 10 min | 0.993 (0.956; 1.053) | 0.991 (0.941; 0.991) | 0.57 | 0.45 |
| 31 m, 10 min | 0.994 (0.957; 1.032) | 0.984 (0.941; 0.987) | 0.56 | 0.79 |
| 16 m, 10 min | 0.989 (0.962; 1.034) | 0.976 (0.948; 0.982) | 0.67 | 0.46 |
| 8 m, 10 min | 0.990 (0.955; 1.030) | 0.973 (0.934; 0.985) | 0.62 | 0.75 |
| 8 m, 60 min | 0.989 (0.953; 1.035) | 0.980 (0.931; 0.988) | 0.51 | 0.90 |

TABLE S3. **Non-significance of MGE-induced contacts across Germany at-large.** Relative detrended daily contacts $c_{w,t} = C_{w,t}/C_t$ across Germany at-large on days without ("non-MGE") vs. with ("MGE") mass gathering events. Contacts are determined using multiple alternative definitions ("resolution"). Median, 25th (Q1) and 75th (Q3) percentiles of $c_{w,t}$ on MGE and non-MGE days are given alongside $p$-values from two-sided Mann-Whitney U tests for the hypothesis that MGE and non-MGE contact levels $c_{w,t}$ are equivalent ("MGE vs. non-MGE"). $p$-values for a negative control in which MGE data is randomly replaced by non-MGE data prior to the statistical test ("non-MGE vs. non-MGE") is also provided. Statistically significant differences ($p < 0.05$) are highlighted by asterisk ($*$).

| host city | device number based in city $n$ | population share [%] $p = n/N$ | ping number $s$ | ping rate [%] $q = a^{-1} \cdot s/n$ |
|---|---|---|---|---|
| Berlin | 7,355 | 0.202 | 18,239,313 | 17.6 |
| Dortmund | 2,265 | 0.386 | 4,895,086 | 15.3 |
| Düsseldorf | 1,418 | 0.229 | 3,720,448 | 18.6 |
| Frankfurt am Main | 1,080 | 0.143 | 2,606,480 | 17.1 |
| Gelsenkirchen | 1,489 | 0.571 | 3,455,363 | 16.4 |
| Hamburg | 3,466 | 0.188 | 8,363,145 | 17.1 |
| Cologne | 1,908 | 0.176 | 4,227,072 | 15.7 |
| Leipzig | 3,128 | 0.532 | 7,637,571 | 17.3 |
| Munich | 1,399 | 0.095 | 2,921,822 | 14.8 |
| Stuttgart | 1,241 | 0.195 | 2,832,907 | 16.2 |
| Germany | 328,817 | 0.388 | 1,599,792,503 | 18.9 |

TABLE S4. **Parameters of population sampling and activity sampling.** Numbers of data panel participants $n$ per city (by home location in June 2024) and across Germany at-large are given. Local population shares $p = n/N$ reflected in the data panel is calculated using values for population sizes $N$ from Tab. S1; for Germany, we assumed $N = 84{,}669{,}000$. Furthermore, the total ping numbers $s$ generated by the $n$ phone users from each city is given (not counting multiple pings per 10 min interval) and used to calculate local ping rates $q = a^{-1} \cdot s/n$ where $a$ represents the number of 10 min time windows in the studied time period (April 29 to August 4, 2024), $a = 98\,\text{days} \cdot 1{,}440\,\text{min/day} \cdot (10\,\text{min})^{-1} = 14{,}112$.

| day (mm/dd) | start time | phase | host city | match | result |
|---|---|---|---|---|---|
| 06/14 | 21:00 | group A, 1st round | Munich | GER–SCO | 5–1 |
| 06/15 | 15:00 | group A, 1st round | Cologne | HUN–SUI | 1–3 |
| 06/15 | 18:00 | group B, 1st round | Berlin | ESP–CRO | 3–0 |
| 06/15 | 21:00 | group B, 1st round | Dortmund | ITA–ALB | 2–1 |
| 06/16 | 15:00 | group D, 1st round | Hamburg | POL–NED | 1–2 |
| 06/16 | 18:00 | group C, 1st round | Stuttgart | SVN–DEN | 1–1 |
| 06/16 | 21:00 | group C, 1st round | Gelsenkirchen | SRB–ENG | 0–1 |
| 06/17 | 15:00 | group E, 1st round | Munich | ROU–UKR | 3–0 |
| 06/17 | 18:00 | group E, 1st round | Frankfurt am Main | BEL–SVK | 0–1 |
| 06/17 | 21:00 | group D, 1st round | Düsseldorf | AUT–FRA | 0–1 |
| 06/18 | 18:00 | group F, 1st round | Dortmund | TUR–GEO | 3–1 |
| 06/18 | 21:00 | group F, 1st round | Leipzig | POR–CZE | 2–1 |
| 06/19 | 15:00 | group B, 2nd round | Hamburg | CRO–ALB | 2–2 |
| 06/19 | 18:00 | group A, 2nd round | Stuttgart | GER–HUN | 2–0 |
| 06/19 | 21:00 | group A, 2nd round | Cologne | SCO–SUI | 1–1 |
| 06/20 | 15:00 | group C, 2nd round | Munich | SVN–SRB | 1–1 |
| 06/20 | 18:00 | group C, 2nd round | Frankfurt am Main | DEN–ENG | 1–1 |
| 06/20 | 21:00 | group B, 2nd round | Gelsenkirchen | ESP–ITA | 1–0 |
| 06/21 | 15:00 | group E, 2nd round | Düsseldorf | SVK–UKR | 1–2 |
| 06/21 | 18:00 | group D, 2nd round | Berlin | POL–AUT | 1–3 |
| 06/21 | 21:00 | group D, 2nd round | Leipzig | NED–FRA | 0–0 |
| 06/22 | 15:00 | group F, 2nd round | Hamburg | GEO–CZE | 1–1 |
| 06/22 | 18:00 | group F, 2nd round | Dortmund | TUR–POR | 0–3 |
| 06/22 | 21:00 | group E, 2nd round | Cologne | BEL–ROU | 2–0 |
| 06/23 | 21:00 | group A, 3rd round | Frankfurt am Main | SUI–GER | 1–1 |
| 06/23 | 21:00 | group A, 3rd round | Stuttgart | SCO–HUN | 0–1 |
| 06/24 | 21:00 | group B, 3rd round | Leipzig | CRO–ITA | 1–1 |
| 06/24 | 21:00 | group B, 3rd round | Düsseldorf | ALB–ESP | 0–1 |
| 06/25 | 18:00 | group D, 3rd round | Berlin | NED–AUT | 2–3 |
| 06/25 | 18:00 | group D, 3rd round | Dortmund | FRA–POL | 1–1 |
| 06/25 | 21:00 | group C, 3rd round | Cologne | ENG–SVN | 0–0 |
| 06/25 | 21:00 | group C, 3rd round | Munich | DEN–SRB | 0–0 |
| 06/26 | 18:00 | group E, 3rd round | Frankfurt am Main | SVK–ROU | 1–1 |
| 06/26 | 18:00 | group E, 3rd round | Stuttgart | UKR–BEL | 0–0 |
| 06/26 | 21:00 | group F, 3rd round | Gelsenkirchen | GEO–POR | 2–0 |
| 06/26 | 21:00 | group F, 3rd round | Hamburg | CZE–TUR | 1–2 |
| 06/29 | 18:00 | round of 16 | Berlin | SUI–ITA | 2–0 |
| 06/29 | 21:00 | round of 16 | Dortmund | GER–DEN | 2–0 |
| 06/30 | 18:00 | round of 16 | Gelsenkirchen | ENG–SVK | 2–1 (AET) |
| 06/30 | 21:00 | round of 16 | Cologne | ESP–GEO | 4–1 |
| 07/01 | 18:00 | round of 16 | Düsseldorf | FRA–BEL | 1–0 |
| 07/01 | 21:00 | round of 16 | Frankfurt am Main | POR–SVN | 3–0 (PEN) |
| 07/02 | 18:00 | round of 16 | Munich | ROU–NED | 0–3 |
| 07/02 | 21:00 | round of 16 | Leipzig | AUT–TUR | 1–2 |
| 07/05 | 18:00 | quarter-finals | Stuttgart | ESP–GER | 2–1 (AET) |
| 07/05 | 21:00 | quarter-finals | Hamburg | POR–FRA | 3–5 (PEN) |
| 07/06 | 18:00 | quarter-finals | Berlin | NED–TUR | 2–1 |
| 07/06 | 21:00 | quarter-finals | Düsseldorf | ENG–SUI | 5–3 (PEN) |
| 07/09 | 21:00 | semi-finals | Munich | ESP–FRA | 2–1 |
| 07/10 | 21:00 | semi-finals | Dortmund | NED–ENG | 1–2 |
| 07/14 | 21:00 | final | Berlin | ESP–ENG | 2–1 |

TABLE S5. **Timetable of UEFA EURO 2024.** For all 51 matches of the UEFA EURO 2024 tournament, the date of the match, time of kick-off, phase, host city, competing national teams (see Tab. S6 for a lookup of FIFA country codes) and the result are indicated. Results tagged with (AET) are achieved after extra time and those tagged with (PEN) after extra time and penalties. Assembled from [35].

| FIFA country code | country |
| --- | --- |
| ALB | Albania |
| AUT | Austria |
| BEL | Belgium |
| CRO | Croatia |
| CZE | Czech Republic |
| DEN | Denmark |
| ENG | England |
| ESP | Spain |
| FRA | France |
| GEO | Georgia |
| GER | Germany |
| HUN | Hungary |
| ITA | Italy |
| NED | Netherlands |
| POL | Poland |
| POR | Portugal |
| ROU | Romania |
| SCO | Scotland |
| SRB | Serbia |
| SUI | Switzerland |
| SVK | Slovakia |
| SVN | Slovenia |
| TUR | Turkey |
| UKR | Ukraine |

TABLE S6. **FIFA country codes.** List of FIFA country codes for participating countries of UEFA EURO 2024 used throughout this manuscript. Assembled from [36].

| match | remark | reference |
|---|---|---|
| GER–DEN | heavy rainfall | [37] |
| ENG–NED | heavy rainfall | [38] |
| TUR–GEO | heavy rainfall | [39] |
| AUT–TUR | heavy rainfall | [40] |
| SVK–ROU | heavy rainfall, fan zone visitors sent home | [41] |
| ENG–SRB | riots in city center at first match in the city | [42] |
| DEN–ENG | crowded city center | [43] |
| FRA–BEL | fan march outside stadium | [44] |
| ALB–ESP | record mobile data usage inside stadium | [45] |
| CZE–TUR | stabbing and temporary evacuation of fan zone | [46] |

TABLE S7. **News articles on notable MGEs covered in the dataset.** Synopsis of relevant news articles reporting on aspects of MGEs covered in this study relevant for actual contact rates.

| | football | | concert | |
|---|---|---|---|---|
| relative time | p-value (MW) | relative change to baseline | p-value (MW) | relative change to baseline |
| −7 | 0.98 | | 0.47 | |
| −6 | 0.66 | | 0.65 | |
| −5 | 0.11 | | 0.18 | |
| −4 | 0.25 | | 0.059 | |
| −3 | 0.035 $*$ | 114.3% (−13.3%; 218.2%) | $2.0 \cdot 10^{-4}$ $*$ | 95.9% (40.4%; 131.6%) |
| −2 | $8.8 \cdot 10^{-3}$ $*$ | 283.3% (−38.0%; 566.7%) | $3.4 \cdot 10^{-5}$ $*$ | 97.9% (64.1%; 158.0%) |
| −1 | $8.1 \cdot 10^{-3}$ $*$ | 242.0% (25.0%; 581.8%) | $7.5 \cdot 10^{-6}$ $*$ | 161.4% (86.3%; 306.3%) |
| 0 | 0.60 | | $1.2 \cdot 10^{-5}$ $*$ | 121.8% (88.9%; 192.3%) |
| 1 | 0.39 | | $1.8 \cdot 10^{-5}$ $*$ | 275.0% (160.7%; 440.0%) |
| 2 | 0.025 $*$ | 110.0% (2.6%; 525.0%) | $7.3 \cdot 10^{-6}$ $*$ | 539.0% (377.8%; 779.9%) |
| 3 | 0.27 | | $6.8 \cdot 10^{-6}$ $*$ | 844.7% (588.2%; 1139.7%) |
| 4 | 0.60 | | $6.8 \cdot 10^{-6}$ $*$ | 1,025.5% (723.5%; 1,595.6%) |
| 5 | 0.75 | | | |

TABLE S8. **Significance of pre- and post-event contacts in host cities.** The table summarizes $p$-values from Mann-Whitney U tests (MW) testing on hourly basis relative to the start time if contact levels in host cities are equivalent on days with UEFA EURO 2024 matches with German participation and non-MGE days ("football"; data from Fig. 3) or on days with Rammstein concerts and non-MGE days ("concert"; data from Fig. S7). In hours with statistically significant differences on MGE days (highlighted by asterisk $*$), the median relative increase in contacts due the MGE and 95 % confidence intervals from bootstrapping MGE and non-MGE data are given.

| public venues & stadium | | home & urban amenities | | commuting | |
|---|---|---|---|---|---|
| map feature | contacts | map feature | contacts | map feature | contacts |
| *amenity:bench | 1904 | *amenity:vending_machine | 588 | surface:asphalt | 1261 |
| *amenity:waste_basket | 1451 | *shop:clothes | 372 | oneway:yes | 998 |
| area:yes | 505 | route:bus | 340 | building:yes | 826 |
| highway:pedestrian | 242 | home:yes | 286 | landuse:residential | 798 |
| sport:soccer | 184 | *amenity:telephone | 278 | route:light_rail | 741 |
| leisure:stadium | 154 | highway:primary | 256 | *tourism:information | 525 |
| tourism:attraction | 99 | *amenity:fast_food | 250 | leisure:park | 498 |
| barrier:wall | 85 | *amenity:toilets | 246 | highway:footway | 437 |
| building:stadium | 78 | route:tram | 234 | barrier:fence | 399 |
| *tourism:viewpoint | 57 | *amenity:restaurant | 141 | route:road | 325 |
| *tourism:museum | 55 | *amenity:atm | 123 | route:bicycle | 254 |
| leisure:pitch | 41 | *amenity:bicycle_rental | 111 | highway:secondary | 201 |
| building:grandstand | 37 | *amenity:lounge | 85 | highway:service | 196 |
| surface:grass | 37 | *shop:bag | 75 | railway:light_rail | 183 |
| | | *amenity:parking_entrance | 67 | route:train | 182 |
| | | *amenity:bank | 58 | landuse:retail | 182 |
| | | | | highway:motorway | 172 |
| | | | | foot:designated | 168 |
| | | | | bicycle:yes | 157 |
| | | | | work:yes | 146 |
| | | | | service:commuter | 140 |
| | | | | railway:platform | 126 |
| | | | | highway:tertiary | 104 |
| | | | | amenity:parking | 89 |
| | | | | building:roof | 86 |

TABLE S9. **Groups of contact settings with similar contact dynamics in host cities of UEFA EURO 2024.** Lists of contact settings in each of three host city contact dynamics profiles (public venues and stadium, home and urban amenities, and commuting) identified for UEFA EURO 2024 matches with German participation (Fig. 4). Numbers of recorded contacts within host cities and within the 6-hour time window around these matches are indicated for all settings. Contact settings are listed in decreasing order of contact numbers. Map features highlighted with asterisk ($^*$) correspond to 0D OSM map features (nodes), while the others correspond to 2D and 1D map features (polygons and lines).

| coverage | category | p-value (MW) | p-value (KS) |
|---|---|---|---|
| 10 cities | recurrent | 0.28 | 0.86 |
| | random | $1.5 \cdot 10^{-3}$ ∗ | 0.043 ∗ |
| Gelsenkirchen | recurrent | 0.16 | 0.47 |
| | random | $1.4 \cdot 10^{-4}$ ∗ | $1.2 \cdot 10^{-3}$ ∗ |

TABLE S10. **MGEs induce random contacts.** The table summarizes $p$-values from one-sided Mann-Whitney U tests (MW) for the hypothesis that contact numbers in the specified category (recurrent, random) are increased on MGE days compared with non-MGE days, separately for the 10 host cities of UEFA EURO 2024 combined (Fig. 5A) and the city of Gelsenkirchen alone (Fig. S11A). Additionally, $p$-values of two-sided Kolmogorov-Smirnov tests (KS) for the hypothesis that contact numbers are equivalent on MGE and non-MGE days are reported. Statistical significance ($p < 0.05$) is highlighted by asterisk (∗).

| coverage | category | p-value (MW) | p-value (KS) |
|---|---|---|---|
| 10 cities | recurrent | 0.48 | $4.4 \cdot 10^{-10}$ ∗ |
| | random | $< 10^{-10}$ ∗ | $< 10^{-10}$ ∗ |
| Gelsenkirchen | recurrent | 0.44 | 0.070 |
| | random | $< 10^{-10}$ ∗ | $< 10^{-10}$ ∗ |

TABLE S11. **MGEs induce small-world contacts.** The table summarizes $p$-values from one-sided Mann-Whitney U tests (MW) for the hypothesis that home-to-home distances in the specified contact category (recurrent, random) are increased on MGE days compared with non-MGE days, separately for the 10 host cities of UEFA EURO 2024 combined (Fig. 5B) and the city of Gelsenkirchen alone (Fig. S11B). Additionally, $p$-values of two-sided Kolmogorov-Smirnov tests (KS) for the hypothesis that home-to-home distances are equivalent on MGE and non-MGE days are reported. Statistical significance ($p < 0.05$) is highlighted by asterisk (∗).

| coverage | category | duration | alternative (MW) | p-value (MW) | p-value (KS) |
|---|---|---|---|---|---|
| 10 cities | recurrent | <10 min | less for MGE | 0.45 | 0.86 |
| | | 10-100 min | greater for MGE | 0.67 | 0.95 |
| | | >100 min | greater for MGE | 0.36 | 0.99 |
| | random | <10 min | less for MGE | $2.9 \cdot 10^{-7}$ ∗ | $7.6 \cdot 10^{-7}$ ∗ |
| | | 10-100 min | greater for MGE | $1.6 \cdot 10^{-7}$ ∗ | $7.6 \cdot 10^{-7}$ ∗ |
| | | >100 min | greater for MGE | $5.4 \cdot 10^{-3}$ ∗ | 0.043 ∗ |
| Gelsenkirchen | recurrent | <10 min | less for MGE | 0.46 | 0.96 |
| | | 10-100 min | greater for MGE | 0.84 | 0.47 |
| | | >100 min | greater for MGE | 0.13 | 0.75 |
| | random | <10 min | less for MGE | $1.8 \cdot 10^{-5}$ ∗ | $4.0 \cdot 10^{-5}$ ∗ |
| | | 10-100 min | greater for MGE | $1.5 \cdot 10^{-5}$ ∗ | $4.0 \cdot 10^{-5}$ ∗ |
| | | >100 min | greater for MGE | 0.046 ∗ | 0.75 |

TABLE S12. **MGEs redistribute random contacts toward intermediate durations.** The table summarizes $p$-values of one-sided Mann-Whitney U tests (MW) for the hypothesis that contacts of the specified category and duration class (<10 min, 10-100 min, >100 min) have a higher or lower relative share on MGE days than on non-MGE days, separately for the 10 host cities of UEFA EURO 2024 combined (Fig. 5B) and the city of Gelsenkirchen alone (Fig. S11B). Additionally, $p$-values of two-sided Kolmogorov-Smirnov tests (KS) for the hypothesis that contact shares per category and duration class are equivalent on MGE and non-MGE days are reported. Statistical significance ($p < 0.05$) is highlighted by asterisk ($*$).

| socializing | | shopping | | routine | |
|---|---|---|---|---|---|
| map feature | contacts | map feature | contacts | map feature | contacts |
| *amenity:bench | 1002 | *shop:bakery | 1348 | *amenity:restaurant | 1449 |
| *amenity:vending_machine | 929 | *shop:supermarket | 1155 | *amenity:atm | 1077 |
| *amenity:bicycle_parking | 888 | *amenity:fast_food | 1126 | *amenity:cafe | 867 |
| *tourism:information | 768 | *shop:clothes | 1037 | *amenity:post_box | 841 |
| *amenity:telephone | 639 | *amenity:pharmacy | 941 | *amenity:waste_basket | 712 |
| *amenity:parking | 565 | *amenity:toilets | 825 | *tourism:viewpoint | 610 |
| *tourism:artwork | 369 | *shop:hairdresser | 803 | *amenity:fuel | 506 |
| *amenity:pub | 321 | *shop:shoes | 614 | *shop:kiosk | 443 |
| *amenity:fountain | 319 | *shop:chemist | 609 | *amenity:doctors | 363 |
| *amenity:ice_cream | 310 | *shop:optician | 454 | *amenity:recycling | 339 |
| fanzone | 237 | *amenity:bank | 447 | *tourism:hotel | 236 |
| *amenity:bar | 220 | *shop:florist | 411 | *shop:convenience | 227 |
| *shop:yes | 153 | *amenity:post_office | 385 | *amenity:clock | 199 |
| *amenity:drinking_water | 127 | *shop:mobile_phone | 376 | *shop:variety_store | 194 |
| *amenity:marketplace | 71 | *shop:butcher | 371 | *shop:beverages | 169 |
| *amenity:biergarten | 65 | *shop:travel_agency | 336 | *amenity:taxi | 159 |
| *amenity:nightclub | 51 | *shop:electronics | 310 | *amenity:car_wash | 154 |
| *shop:massage | 48 | *shop:books | 310 | *shop:deli | 152 |
| | | *shop:jewelry | 302 | *shop:bicycle | 147 |
| | | *shop:beauty | 283 | *amenity:photo_booth | 145 |
| | | *amenity:charging_station | 256 | *shop:department_store | 135 |
| | | *shop:sports | 248 | *shop:furniture | 130 |
| | | *shop:gift | 226 | *shop:tobacco | 121 |
| | | *shop:stationery | 220 | *shop:car_repair | 115 |
| | | *amenity:parking_entrance | 206 | *amenity:bicycle_rental | 111 |
| | | *shop:newsagent | 195 | *tourism:attraction | 106 |
| | | *shop:perfumery | 182 | *shop:tailor | 103 |
| | | *shop:toys | 175 | *shop:interior_decoration | 101 |
| | | *amenity:dentist | 162 | *shop:coffee | 96 |
| | | *shop:confectionery | 162 | *shop:photo | 93 |
| | | *shop:dry_cleaning | 123 | *amenity:driving_school | 92 |
| | | *shop:pet | 117 | *shop:ticket | 92 |
| | | *shop:vacant | 108 | *amenity:kindergarten | 89 |
| | | *shop:cosmetics | 98 | *amenity:library | 89 |
| | | *shop:car | 93 | *amenity:compressed_air | 86 |
| | | *shop:doityourself | 86 | *amenity:social_facility | 84 |
| | | *amenity:car_rental | 85 | *shop:medical_supply | 76 |
| | | *shop:greengrocer | 77 | *tourism:museum | 76 |
| | | *shop:tea | 72 | *amenity:school | 72 |
| | | *shop:video_games | 72 | *amenity:place_of_worship | 70 |
| | | *shop:bag | 70 | *shop:hearing_aids | 68 |
| | | *shop:outdoor | 62 | *amenity:police | 61 |
| | | *shop:fashion_accessories | 35 | *amenity:theatre | 59 |
| | | *shop:alcohol | 31 | *shop:houseware | 56 |
| | | *amenity:bureau_de_change | 23 | *shop:boutique | 56 |
| | | *tourism:guest_house | 23 | *shop:laundry | 55 |
| | | | | *shop:computer | 53 |
| | | | | *shop:erotic | 47 |
| | | | | *amenity:grit_bin | 46 |
| | | | | *amenity:cinema | 44 |
| | | | | *shop:locksmith | 41 |
| | | | | *amenity:community_centre | 38 |
| | | | | *shop:bed | 37 |
| | | | | *amenity:food_court | 33 |
| | | | | *amenity:shelter | 33 |
| | | | | *amenity:casino | 22 |
| | | | | *shop:art | 20 |

TABLE S13. **Groups of contact settings with similar contact dynamics in Germany on the day of UEFA EURO 2024 quarter-final ESP–GER.** Lists of contact settings in each of three nationwide contact dynamics profiles (socializing, shopping, and routine) identified for the UEFA EURO 2024 quarter-final match ESP–GER (Fig. 6). Numbers of recorded contacts across Germany on the day of the match (July 5, 2024) are indicated for all settings. Contact settings are listed in decreasing order of contact numbers. This analysis takes only 0D OSM map features (nodes; highlighted with asterisk $*$) into account.

| city | day (mm/dd) | MGE | capacity/audience | reference |
|---|---|---|---|---|
| Berlin | 05/04 | Mario Barth | 22,000 | Waldbühne [47] |
| Berlin | 05/05 | S25 Berlin | Tab. S1 | |
| Berlin | 05/05 | Bundesliga | 22,000 | Stadion An der Alten Försterei |
| Berlin | 05/11 | Bundesliga | Tab. S1 | |
| Berlin | 05/18 | Bundesliga | 22,000 | Stadion An der Alten Försterei |
| Berlin | 05/25 | DFB Pokal | Tab. S1 | |
| Berlin | 06/01 | Schlagernacht | 17,000 | Uber Arena |
| Berlin | 06/09 | Regionalliga (female) | 22,000 | Stadion An der Alten Försterei |
| Berlin | 06/10 | Green Day | 22,000 | Waldbühne [48] |
| Berlin | 06/15 | ESP–CRO | Tab. S1 | |
| Berlin | 06/21 | POL–AUT | Tab. S1 | |
| Berlin | 06/25 | NED–AUT | Tab. S1 | |
| Berlin | 06/29 | SUI–ITA | Tab. S1 | |
| Berlin | 07/06 | NED–TUR | Tab. S1 | |
| Berlin | 07/13 | Berliner Rundfunk Open Air | 17,000 | Parkbühne Wuhlheide [49] |
| Berlin | 07/14 | ESP–ENG | Tab. S1 | |
| Berlin | 07/27 | friendly match | 22,000 | Stadion An der Alten Försterei |
| Berlin | 08/03 | Bundesliga | Tab. S1 | |
| Berlin | 08/03 | friendly match | 22,000 | Stadion An der Alten Försterei |
| Dortmund | 05/01 | UEFA Champions League | Tab. S1 | |
| Dortmund | 05/04 | Bundesliga | Tab. S1 | |
| Dortmund | 05/06 | Apache 207 | 13,000 | Westfalenhalle [50] |
| Dortmund | 05/07 | B2Run | Tab. S1 | |
| Dortmund | 05/18 | Bundesliga | Tab. S1 | |
| Dortmund | 05/19 | Schlagerfest XXL | 9,000 | Westfalenhalle [51] |
| Dortmund | 05/24 | Dogs & Fun | 25,000 | Messe Dortmund [52] |
| Dortmund | 06/01 | UEFA Champions League | Tab. S1 | |
| Dortmund | 06/15 | ITA–ALB | Tab. S1 | |
| Dortmund | 06/18 | TUR–GEO | Tab. S1 | |
| Dortmund | 06/22 | TUR–POR | Tab. S1 | |
| Dortmund | 06/25 | FRA–POL | Tab. S1 | |
| Dortmund | 06/29 | GER–DEN | Tab. S1 | |
| Dortmund | 07/10 | ENG–NED | Tab. S1 | |
| Dortmund | 08/03 | Bundesliga (3rd) | Tab. S1 | |
| Dortmund | 08/03 | Dortmund OLE | 20,000 | Revierpark Wischlingen [53] |
| Düsseldorf | 05/03 | Bundesliga | Tab. S1 | |
| Düsseldorf | 05/19 | Bundesliga | Tab. S1 | |
| Düsseldorf | 05/27 | Bundesliga (relegation) | Tab. S1 | |
| Düsseldorf | 06/01 | Japan Day | 630,000 | downtown Düsseldorf [54] |
| Düsseldorf | 06/17 | AUT–FRA | Tab. S1 | |
| Düsseldorf | 06/21 | SVK–UKR | Tab. S1 | |
| Düsseldorf | 06/24 | ALB–ESP | Tab. S1 | |
| Düsseldorf | 06/28 | DoKomi | 60,000 | Messe Düsseldorf [55] |
| Düsseldorf | 06/29 | DoKomi | 60,000 | Messe Düsseldorf [55] |
| Düsseldorf | 06/30 | DoKomi | 60,000 | Messe Düsseldorf [55] |
| Düsseldorf | 07/01 | FRA–BEL | Tab. S1 | |
| Düsseldorf | 07/06 | ENG–SUI | Tab. S1 | |
| Düsseldorf | 07/19 | Rheinkirmes | 360,000 | Oberkassler Festwiesen [56] |
| Düsseldorf | 07/20 | Coldplay | Tab. S1 | |
| Düsseldorf | 07/21 | Coldplay | Tab. S1 | |
| Düsseldorf | 07/23 | Coldplay | Tab. S1 | |
| Frankfurt am Main | 05/05 | Bundesliga | Tab. S1 | |
| Frankfurt am Main | 05/18 | Bundesliga | Tab. S1 | |
| Frankfurt am Main | 05/25 | Hessenpokal | 12,000 | PSD Bank Arena |
| Frankfurt am Main | 05/27 | Apache 207 | 13,500 | Festhalle Frankfurt |
| Frankfurt am Main | 06/17 | BEL–SVK | Tab. S1 | |
| Frankfurt am Main | 06/18 | Five Finger Death Punch | 15,000 | Festhalle [57] |
| Frankfurt am Main | 06/20 | DEN–ENG | Tab. S1 | |
| Frankfurt am Main | 06/23 | SUI–GER | Tab. S1 | |
| Frankfurt am Main | 06/26 | SVK–ROM | Tab. S1 | |

| city | day (mm/dd) | MGE | capacity/audience | reference |
|---|---|---|---|---|
| Frankfurt am Main | 06/30 | American Football | 6,700 | PSD Bank Arena [58] |
| Frankfurt am Main | 07/01 | POR–SLO | Tab. S1 | |
| Frankfurt am Main | 07/11 | Rammstein | 40,000 | Waldstadion [59] |
| Frankfurt am Main | 07/12 | Rammstein | 40,000 | Waldstadion [59] |
| Frankfurt am Main | 07/13 | Rammstein | 40,000 | Waldstadion [59] |
| Frankfurt am Main | 07/13 | American Football | 6,700 | PSD Bank Arena [60] |
| Frankfurt am Main | 07/18 | Peter Maffay | 39,000 | Deutsche Bank Park [61] |
| Frankfurt am Main | 07/20 | Roland Kaiser | 25,000 | Deutsche Bank Park [62] |
| Frankfurt am Main | 07/26 | Travis Scott | Tab. S1 | |
| Frankfurt am Main | 07/27 | Travis Scott | Tab. S1 | |
| Gelsenkirchen | 05/11 | Bundesliga | Tab. S1 | |
| Gelsenkirchen | 05/17 | AC/DC | 55,000 | Veltins-Arena [63] |
| Gelsenkirchen | 05/21 | AC/DC | 54,000 | Veltins-Arena [64] |
| Gelsenkirchen | 06/16 | SRB–ENG | Tab. S1 | |
| Gelsenkirchen | 06/20 | ESP–ITA | Tab. S1 | |
| Gelsenkirchen | 06/26 | GEO–POR | Tab. S1 | |
| Gelsenkirchen | 06/30 | ENG–SVK | Tab. S1 | |
| Gelsenkirchen | 07/17 | Taylor Swift | 60,000 | Veltins-Arena [65] |
| Gelsenkirchen | 07/18 | Taylor Swift | 60,000 | Veltins-Arena [65] |
| Gelsenkirchen | 07/19 | Taylor Swift | 60,000 | Veltins-Arena [65] |
| Gelsenkirchen | 07/21 | Schalke Tach | 75,000 | Veltins-Arena [66] |
| Gelsenkirchen | 07/26 | Rammstein | 60,000 | Veltins-Arena [67] |
| Gelsenkirchen | 07/27 | Rammstein | 60,000 | Veltins-Arena [67] |
| Gelsenkirchen | 07/29 | Rammstein | 60,000 | Veltins-Arena [67] |
| Gelsenkirchen | 07/30 | Rammstein | 60,000 | Veltins-Arena [67] |
| Gelsenkirchen | 07/31 | Rammstein | 60,000 | Veltins-Arena [67] |
| Gelsenkirchen | 08/03 | Bundesliga | Tab. S1 | |
| Hamburg | 05/03 | Bundesliga | Tab. S1 | |
| Hamburg | 05/11 | Hafengeburtstag | 375,000 | Elbe waterfront [68] |
| Hamburg | 05/12 | Bundesliga | 30,000 | Millerntor-Stadion |
| Hamburg | 05/17 | Marius Müller-Westernhagen | 11,000 | Barclays Arena [69] |
| Hamburg | 05/19 | Bundesliga | Tab. S1 | |
| Hamburg | 05/25 | Regionalliga (female) | 30,000 | Millerntor-Stadion |
| Hamburg | 06/01 | Juste Debout x Kampnage | 30,000 | Millerntor-Stadion |
| Hamburg | 06/16 | POL–NED | Tab. S1 | |
| Hamburg | 06/19 | ALB–CRO | Tab. S1 | |
| Hamburg | 06/22 | GEO–CZE | Tab. S1 | |
| Hamburg | 06/26 | CZE–TUR | Tab. S1 | |
| Hamburg | 07/05 | POR–FRA | Tab. S1 | |
| Hamburg | 07/14 | American Football | Tab. S1 | |
| Hamburg | 07/23 | Taylor Swift | 50,000 | Volksparkstadion [70] |
| Hamburg | 07/24 | Taylor Swift | 50,000 | Volksparkstadion [70] |
| Cologne | 05/04 | Bundesliga | Tab. S1 | |
| Cologne | 05/09 | DFB Pokal | Tab. S1 | |
| Cologne | 05/11 | Bundesliga | Tab. S1 | |
| Cologne | 05/24 | Howard Carpendale | 20,000 | LANXESS Arena |
| Cologne | 06/15 | HUN–SUI | Tab. S1 | |
| Cologne | 06/19 | SCO–SUI | Tab. S1 | |
| Cologne | 06/22 | BEL–ROM | Tab. S1 | |
| Cologne | 06/25 | ENG–SLO | Tab. S1 | |
| Cologne | 06/30 | ESP–GEO | Tab. S1 | |
| Cologne | 07/12 | Peter Maffay | 37,000 | Rhein-Energie-Stadion [71] |
| Cologne | 07/13 | Roland Kaiser | 42,000 | Rhein-Energie-Stadion [72] |
| Cologne | 07/20 | Christopher Street Day | 200,000 | downtown Cologne [73] |
| Cologne | 07/20 | Travis Scott | Tab. S1 | |
| Cologne | 07/21 | Christopher Street Day | 1,200,000 | downtown Cologne [73] |
| Cologne | 08/02 | Bundesliga | Tab. S1 | |
| Leipzig | 05/08 | Apache 207 | 12,000 | QUARTERBACK Immobilien ARENA [74] |
| Leipzig | 05/11 | Bundesliga | Tab. S1 | |
| Leipzig | 05/19 | Leipziger Weinfest | 27,500 | downtown Leipzig [75] |

| city | day (mm/dd) | MGE | capacity/audience | reference |
|---|---|---|---|---|
| Leipzig | 06/02 | Leipziger Stadtfest | 100,000 | downtown Leipzig [76] |
| Leipzig | 06/04 | Andre Rieu | 7,500 | QUARTERBACK Immobilien ARENA [77] |
| Leipzig | 06/06 | Andre Rieu | 7,500 | QUARTERBACK Immobilien ARENA [77] |
| Leipzig | 06/18 | POR–CZE | Tab. S1 | |
| Leipzig | 06/21 | NED–FRA | Tab. S1 | |
| Leipzig | 06/24 | CRO–ITA | Tab. S1 | |
| Leipzig | 07/02 | AUT–TUR | Tab. S1 | |
| Leipzig | 07/17 | P!NK | 43,000 | Red Bull Arena [78] |
| Leipzig | 07/19 | Roland Kaiser | 44,000 | Red Bull Arena [79] |
| Leipzig | 07/20 | Peter Maffay | 38,000 | Red Bull Arena [80] |
| Leipzig | 08/02 | SDP | 12,000 | QUARTERBACK Immobilien ARENA [81] |
| Munich | 04/30 | UEFA Champions League | Tab. S1 | |
| Munich | 05/12 | Bundesliga | Tab. S1 | |
| Munich | 05/24 | Metallica | 75,000 | Olympiastadion München [82] |
| Munich | 05/26 | Metallica | 75,000 | Olympiastadion München [82] |
| Munich | 06/09 | AC/DC | 66,000 | Olympiastadion München [83] |
| Munich | 06/12 | AC/DC | 66,000 | Olympiastadion München [83] |
| Munich | 06/14 | GER–SCO | Tab. S1 | |
| Munich | 06/17 | ROM–UKR | Tab. S1 | |
| Munich | 06/20 | SLO–SRB | Tab. S1 | |
| Munich | 06/22 | Andreas Gabalier | 60,000 | Olympiastadion München [84] |
| Munich | 06/25 | DEN–SRB | Tab. S1 | |
| Munich | 06/29 | ANTENNE BAYERN Open Air | 10,000 | UEFA EURO 2024 fan zone [85] |
| Munich | 07/02 | ROM–NED | Tab. S1 | |
| Munich | 07/05 | ANTENNE BAYERN Open Air | 10,000 | UEFA EURO 2024 fan zone [85] |
| Munich | 07/08 | Tollwood | 6,000 | Olympiapark [86] |
| Munich | 07/09 | ESP–FRA | Tab. S1 | |
| Munich | 07/13 | ANTENNE BAYERN Open Air | 10,000 | UEFA EURO 2024 fan zone [85] |
| Munich | 07/17 | B2Run | 30,000 | Olympiapark [87] |
| Munich | 07/18 | Tollwood | 6,000 | Olympiapark [86] |
| Munich | 07/27 | Taylor Swift | 70,000 | Olympiastadion München [88] |
| Munich | 07/28 | Taylor Swift | 70,000 | Olympiastadion München [88] |
| Stuttgart | 04/30 | Bushido | 6,000 | Porsche-Arena [89] |
| Stuttgart | 05/04 | Bundesliga | Tab. S1 | |
| Stuttgart | 05/12 | Apache 207 | 15,000 | Hanns-Martin-Schleyer-Halle |
| Stuttgart | 05/18 | Bundesliga | Tab. S1 | |
| Stuttgart | 05/31 | Bulent Ceylan | 6,500 | Porsche-Arena |
| Stuttgart | 06/16 | SLO–DEN | Tab. S1 | |
| Stuttgart | 06/19 | GER–HUN | Tab. S1 | |
| Stuttgart | 06/23 | SCO–HUN | Tab. S1 | |
| Stuttgart | 06/26 | UKR–BEL | Tab. S1 | |
| Stuttgart | 06/27 | fan zone concerts | 30,000 | UEFA EURO 2024 fan zone [90] |
| Stuttgart | 07/05 | ESP–GER | Tab. S1 | |
| Stuttgart | 07/11 | Five Finger Death Punch | 15,000 | Hanns-Martin-Schleyer-Halle |
| Stuttgart | 07/15 | Peter Maffay | 16,000 | Cannstatter Wasen [91] |
| Stuttgart | 07/16 | Peter Maffay | 16,000 | Cannstatter Wasen [91] |
| Stuttgart | 07/17 | AC/DC | 90,000 | Cannstatter Wasen [92] |
| Stuttgart | 07/19 | P!NK | 45,000 | MHP Arena [93] |
| Stuttgart | 08/03 | SDP | 30,000 | Cannstatter Wasen [94] |

TABLE S14. **MGEs with measurable footprint in GPS co-location contacts.** List of 169 MGEs held across 10 cities between April 29 and August 4, 2024, including various football matches, the UEFA EURO 2024 tournament, concerts, shows, festivals and fairs. For each MGE, the host city, day, name, host venue as well as the audience size (if available; reference is provided) or host venue capacity are indicated. Names are given as country code pairs of competing national teams in the case of UEFA EURO 2024 matches, artists' names in the case of concerts and shows, proper names in the case of festivals and fairs, or summary labels such as "Bundesliga" for Bundesliga matches.
